# Global and national influenza-associated hospitalization and mortality rates: a systematic review and meta-analysis

**DOI:** 10.1101/2025.08.08.25333141

**Authors:** Ya Gao, Ming Liu, Kenji Numata, Yamin Chen, Yunli Zhao, Yanjiao Shen, Zhifan Li, Wimonchat Tangamornsuksun, Jianguo Xu, Qingyong Zheng, Wanyu Zhao, Xianlin Gu, Liang Zhao, Luying Cheng, Liang Yao, Jiyuan Shi, Li Zheng, Mary Katharine Kennedy, Liangying Hou, Fuzhong Xue, Jinhui Tian, Gordon Guyatt, Qiukui Hao

## Abstract

**Background:** A global comprehensive assessment of influenza-associated hospitalization and mortality across diverse populations and geographical settings is currently lacking. To support an update of WHO influenza clinical guidelines, this systematic review and meta-analysis evaluated the baseline risks of hospitalization and all-cause mortality in patients with seasonal, pandemic, or zoonotic influenza.

**Methods:** We systematically searched Medline, Embase, CENTRAL, CINAHL, and Global Health for studies published through 26 October 2023, that reported hospitalization and/or mortality among patients with laboratory-confirmed influenza. Paired reviewers independently identified studies, extracted data, and assessed the risk of bias. We performed inverse variance method fixed effects model meta-analyses to summarize the evidence and evaluated the certainty of evidence using the GRADE (Grading of Recommendations Assessment, Development and Evaluation) approach. We registered the protocol with PROSPERO, CRD42023456645.

**Findings:** Of 30054 records, 355 studies with 18,640,366 patients (mean age 0.1 to 87.9 years) proved eligible. The hospitalization rate was 1.01% (95%CI 1% to 1.01%; moderate certainty) for patients with seasonal or pandemic influenza and 93.72% (95%CI 90.12% to 96.58 %; low certainty) for patients with zoonotic influenza. The all-cause mortality rate was 0.23% (95%CI 0.22% to 0.24%; high certainty) for patients with non-severe seasonal or pandemic influenza, 4.15% (95%CI 4.10% to 4.20%; moderate certainty) for patients with severe seasonal or pandemic influenza, and 38.72% (95%CI 36.79% to 40.66%; moderate certainty) for patients with zoonotic influenza.

**Interpretation:** This review estimated a hospitalization rate of 1.01% for seasonal or pandemic influenza and all-cause mortality rates of 0.23% for non-severe seasonal or pandemic influenza, 4.15% for severe seasonal or pandemic influenza, and 38.72% for severe zoonotic influenza.

**Funding:** World Health Organization to McMaster University in 2023.

**Research in context:** *Evidence before this study:* A comprehensive understanding of influenza-associated hospitalization and mortality rates is essential for guiding effective prevention and control strategies and formulating evidence-based clinical practice guidelines and policies. Previous systematic reviews and meta-analyses have investigated influenza-associated hospitalization and mortality rates, but they have often been limited by their focus on a specific strain of influenza, particular patient subgroups, or individual countries, or by a lack of assessment of the certainty of evidence. A global comprehensive assessment of influenza-associated hospitalization and mortality rates across diverse strains, populations, and geographical settings is currently lacking.

*Added value of this study:* We found a hospitalization rate of 1.01% for seasonal or pandemic influenza and all-cause mortality rates of 0.23% for non-severe seasonal or pandemic influenza, 4.15% for severe seasonal or pandemic influenza, and 38.72% for zoonotic influenza. Among severe seasonal or pandemic influenza cases, patients aged ≥65 years (8.31%) had a higher all-cause mortality rate than those aged 0-14 years (0.76%) and 15-64 years (6.50%).

*Implications of all the available evidence:* Our study provides a comprehensive assessment of the global evidence regarding hospitalization and mortality rates among influenza patients. Our findings highlight the critical need for distinct clinical and public health strategies tailored to influenza type and severity.

## Introduction

Influenza, an infectious respiratory illness caused by the influenza virus, remains a significant global public health concern.^1,2^ Presenting with symptoms such as fever and cough, influenza can progress to severe illness, leading to hospitalization and death, particularly among vulnerable populations such as children and adults aged 65 years or older.^3–6^ According to estimates published on the World Health Organization (WHO) website, influenza annually causes 3 to 5 million severe cases and contributes to approximately 290,000 to 650,000 respiratory-related deaths worldwide, highlighting its substantial impact on healthcare systems.^7,8^

A comprehensive understanding of influenza-associated hospitalization and mortality rates is essential for guiding effective prevention and control strategies and formulating evidence-based clinical practice guidelines and policies.^9^ Previous systematic reviews^9–14^ have investigated influenza-associated hospitalization and mortality rates. However, these studies often narrowed their focus on a specific strain of influenza (e.g. seasonal influenza)^10,11,14^, particular patient subgroups (e.g. children, indigenous populations)^3,10,11^, or individual countries (e.g. Italy, China).^9,13^ Furthermore, previous reviews have not assessed the certainty of evidence. A global comprehensive assessment of influenza-associated hospitalization and mortality rates across diverse strains, populations, and geographical settings is currently lacking.

To determine the baseline risk estimates of death and hospitalization for influenza, in support of an update of the WHO influenza guidelines^15^, we conducted a comprehensive systematic review and meta-analysis of observational studies. This review provided estimates for the hospitalization and mortality rates among patients with influenza virus infection globally and within specific countries, across patient groups, and across different influenza etiologies, considering the certainty of evidence.

### Methods

This systematic review adhered to the Preferred Reported Items for Systematic Reviews and Meta-Analyses 2020 (PRISMA 2020) statement and Meta-analysis Of Observational Studies in Epidemiology (MOOSE) guidelines.^16,17^ We registered the systematic review protocol with PROSPERO (CRD42023456645).

### Search strategy

With the aid of an expert librarian, we searched Medline, Embase, Cochrane Central Register of Controlled Trials (CENTRAL), Cumulative Index to Nursing and Allied Health Literature (CINAHL), and Global Health from database inception to October 26, 2023. The search terms included “influenza”, “hospital”, “death”, “mortality”, and “fatality”. To identify additional eligible studies, we screened the reference lists of eligible studies and relevant systematic reviews. Appendix 1 presents the details of the search strategy.

### Eligibility criteria and study selection

We included studies of patients with influenza virus infections confirmed by PCR assay or other nucleic acid amplification assays, rapid antigen test, or immunofluorescence assay reporting hospitalization and/or mortality among influenza patients. We also included studies enrolling patients with influenza-like illness without clear mention of the laboratory confirmation in the diagnosis if the proportion of such patients was 20% or less, or if authors reported separately on our population of interest. To reduce inclusion of overlapping data among studies, we included only cohort studies or studies with surveillance data.

We did not apply restrictions on publication language, age and sex of patients, influenza severity, strains of influenza viruses, or comorbidity of patients. We excluded narrative, scoping, systematic reviews, randomized controlled trials, case reports, and conference abstracts and studies with hospitalization and mortality estimated with modelling techniques.

Using Covidence (https://covidence.org/), paired reviewers independently screened titles and abstracts and, subsequently, reviewed the full texts of potentially eligible records. When eligible studies used the same source of data and sampling period, we included the study with a larger sample size. Reviewers resolved disagreements by discussion or, if necessary, by consultation with a third reviewer.

### Data extraction

Using a predesigned form, pairs of reviewers independently performed data extraction and resolved discrepancies by discussion or, if necessary, with the assistance of a third party. Reviewers extracted the following information: study characteristics (first author, setting, study type, publication year, study period, country, and sample size); patient characteristics (age, sex, disease severity, comorbidities, vaccination status, influenza virus strains, and influenza etiology); and data on hospitalization and mortality (e.g., all-cause mortality, influenza-related mortality, or in-hospital mortality, if available).

### Risk of bias assessment

Using the Quality In Prognosis Studies (QUIPS) tool, pairs of reviewers independently assessed the risk of bias of eligible studies.^18^ We evaluated four domains: study participation, study attrition, outcome measurement, and statistical analysis and reporting. We rated each domain as either low, moderate, or high risk of bias. We considered studies at overall high risk of bias if we judged one or more domains at high risk of bias; studies at low risk of bias if we judged three domains at low risk of bias and the other domain at moderate risk of bias, or all domains at low risk of bias. We judged the remaining studies at moderate risk of bias. Reviewers resolved discrepancies by discussion and, when necessary, with adjudication by a third reviewer.

### Statistical analysis

To estimate pooled hospitalization and mortality rates and their associated 95% confidence intervals (CIs), we conducted meta-analyses of proportions. To minimize small study effects, we used the inverse variance method fixed effects model.^19^ To stabilize variances, we applied the Freeman-Tukey double arcsine transformation.^20^ We assessed the between-study heterogeneity with a visual inspection of forest plots. For mortality, we performed analyses for severe and non-severe influenza separately, defining severe influenza as an illness with laboratory-confirmed influenza that requires hospitalization or according to specific study definitions.^21^ Non-severe influenza cases were those that did not meet the criteria for severe illness.

We classified studies as predominantly non-severe if they comprised 80% or more non-severe patients, as predominantly severe if they included 80% or more severe patients, and as mixed-severity if they did not meet either of these criteria. We followed published guidance suggesting avoiding statistical tests in the context of meta-analysis of proportions and so did not assess publication bias.^20^ To ensure the robustness of results, we performed sensitivity analyses including only studies with a low risk of bias. We performed analyses using R (version 4.2.1, R Foundation for Statistical Computing).

### Subgroup analysis

To provide comprehensive summaries of current evidence and explore potential heterogeneity, we estimated hospitalization and mortality rates for different subgroups. If data were available (at least two studies in each of the subgroups of interest providing relevant information), we performed the following prespecified subgroup analyses:

1. Countries: low- and middle-income countries (LMICs) versus high-income countries (HICs) (hypothesis: higher hospitalization and mortality rates in LMICs).
2. Influenza etiology: seasonal versus pandemic influenza viruses (hypothesis: higher hospitalization and mortality rates in patients with pandemic influenza virus infection than seasonal influenza virus infection).
3. Age: infants (< 2 years) and children (2-14 years) versus adults and adolescents (15-64 years) versus elderly (≥ 65 years) (hypothesis: higher hospitalization and mortality rates in infants, children, or elderly).
4. Vaccination status: vaccinated versus unvaccinated patients (hypothesis: higher hospitalization and mortality rates in unvaccinated patients).
5. Chronic co-morbidities (e.g. cardiovascular disease, respiratory disease, neurological disease, extreme obesity, immunocompromised patients): patients with at least one chronic comorbidity versus those without (hypothesis: higher hospitalization and mortality rates in patients with at least one chronic comorbidity).

We prioritized the use of within-study subgroup analyses rather than between-study subgroup analyses. To assess the credibility of the apparent subgroup effect, we used a modified version of the Instrument for assessing the Credibility of Effect Modification Analyses (ICEMAN) tool tailored for prognostic studies.^22^

### Certainty of evidence

To assess the certainty of the evidence regarding the proportions of each outcome, we used the GRADE (Grading of Recommendations Assessment Development, and Evaluation) guidance for overall prognosis in broad populations.^23^ By assessing the domains of risk of bias^24^, imprecision around the pooled estimate^25^, inconsistency across studies^26^, and indirectness of evidence identified^27^, we rated the overall certainty of the evidence for each outcome as ‘‘high’’, ‘‘moderate’’, ‘‘low’’, or ‘‘very low’’. We assessed imprecision using the minimally important difference (MID) as a threshold and specified the MID as 4% for mortality and hospitalization.

## Results

The electronic search identified 30,054 records. After screening 16381 unduplicated titles and abstracts and 1991 full texts, 355 studies proved eligible (Figure 1). Appendix 2 lists the eligible studies.

**Figure 1.**
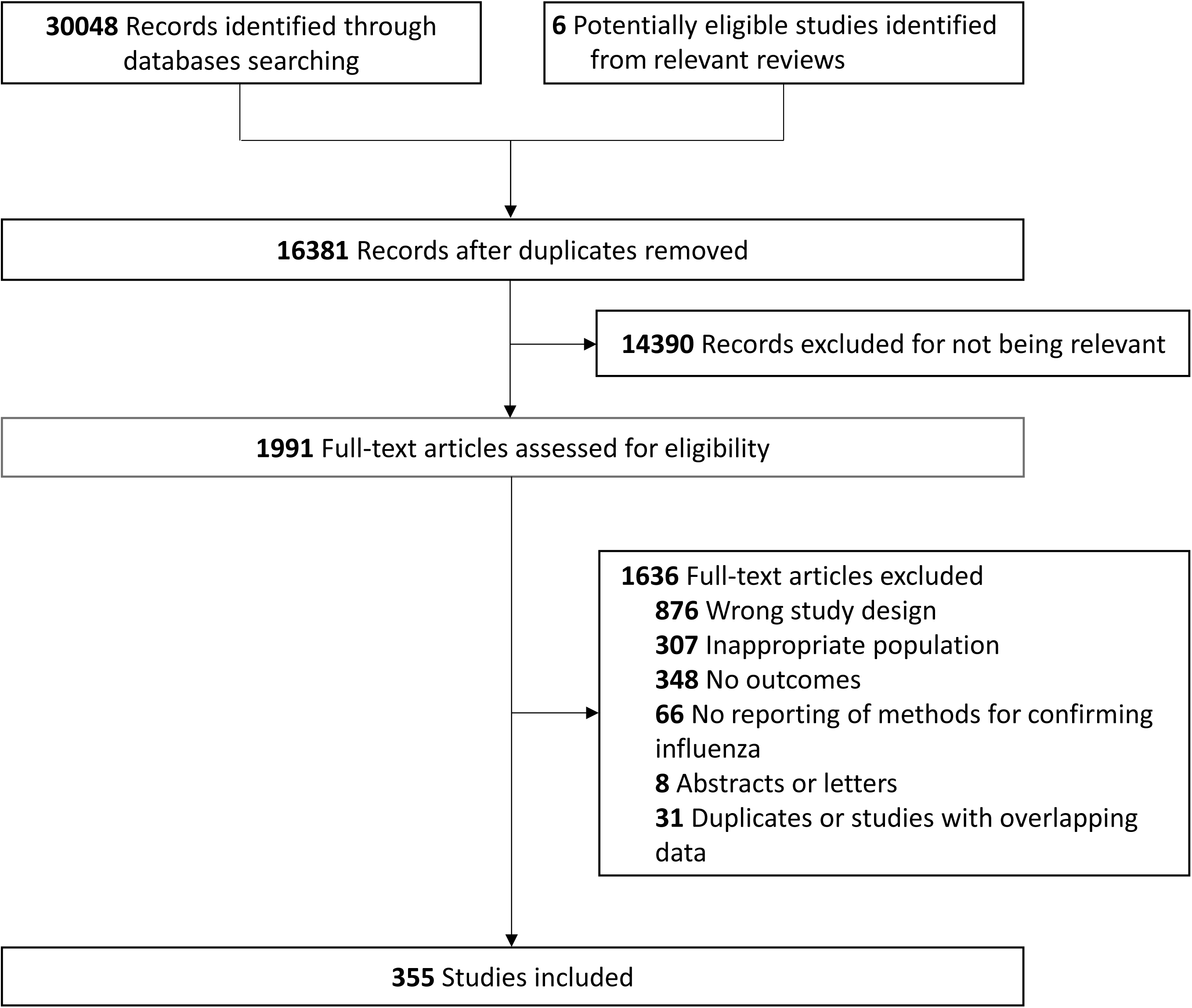
Study selection

### Characteristics of included studies

Appendix 3 summarizes the characteristics of the 355 eligible studies. The number of patients in the included studies ranged from 17 to 16,636,913, including a total of 18,640,366 patients. The median mean age was 43.4 years (range from 0.1 to 87.9 years) and the median proportion of male patients was 52.5% (range from 0% to 94.6%). Seventy-five studies reported hospitalization and 339 reported all-cause mortality. Due to unclear reporting, we could not ascertain in-hospital mortality or influenza-related mortality and so assumed that all deaths were related to influenza. Of the 339 studies that reported all-cause mortality, 21 included primarily non-severe patients, 275 studies included primarily severe influenza patients, 37 included both non-severe and severe patients, and 6 did not report sufficient information to determine the severity of patients.

### Risk of bias

Of 75 studies that reported hospitalization, 73 had low and 2 had high risk of bias. Of the 339 studies that reported all-cause mortality, 333 had low, 3 had moderate, and 3 had high risk of bias. The main limitations included insufficient descriptions of study participation and substantial attrition rates (Appendix 4).

### Hospitalization

#### Seasonal or pandemic influenza

Moderate certainty evidence showed the hospitalization rate of patients with seasonal or pandemic influenza was 1.01% (95%CI 1.00% to 1.01%; 74 studies, 17,715,918 patients) (Table 1 and Appendix 5). Within-study subgroup analysis revealed significant difference in hospitalization rate between patients aged up to 14 years and patients aged 65 years or older (pooled ratio of rate 0.84, 95% CI: 0.73 to 0.96, P for interaction = 0.01, Table 2 and Appendix 6). However, due to the test of many effect modifiers and chance remaining a likely explanation for findings, we rated the credibility of this subgroup effect as low (Appendix 7). We did not find a subgroup effect between vaccinated patients and unvaccinated patients in the hospitalization rate (Table 2 and Appendix 8). Between-study subgroup analyses indicated that patients in HICs had a lower hospitalization rate (0.95% versus 20.50%, P for interaction < 0.01, Appendix 9) than patients in LMICs, patients with seasonal influenza had a lower hospitalization rate (1.02% versus 6.70%, P for interaction < 0.01, Appendix 10) than patients with pandemic influenza, and patients without chronic comorbidities had a lower hospitalization rate (P for interaction < 0.01, Appendix 11) than patients with chronic comorbidities (Table 2). However, due to the between-study comparison and test of many effect modifiers, we rated the credibility of these subgroup effects as low (Appendix 7). Sensitivity analysis only including studies with a low risk of bias showed a similar result (1.01%, 95%CI 1.00% to 1.01%) to the primary analysis (Appendix 12).

**Table 1.** GRADE assessment for hospitalization and all-cause mortality rates in influenza patients.

| Outcomes | No of studies | Effect |  |  | Certainty of the evidence |
| --- | --- | --- | --- | --- | --- |
|  |  | No of events | No of individuals | Rate (95% CI) |  |
| Hospitalization: Seasonal or pandemic influenza | 74 | 239858 | 17715918 | event rate<br>10 per 1000 (10 to 10.1) | ⊕⊕⊕○<br>Moderate* |
| Hospitalization: Zoonotic influenza | 1 | 209 | 223 | event rate<br>937 per 1000 (901 to 966) | ⊕⊕○○<br>Low†‡ |
| Mortality: Non-severe seasonal or pandemic influenza | 23 | 2698 | 788548 | event rate<br>2.3 per 1000 (2.2 to 2.4) | ⊕⊕⊕⊕<br>High |
| Mortality: Severe seasonal or pandemic influenza | 269 | 38053 | 709672 | event rate<br>42 per 1000 (41 to 42) | ⊕⊕⊕○<br>Moderate* |
| Mortality: Severe zoonotic influenza | 9 | 972 | 2500 | event rate<br>387 per 1000 (368 to 407) | ⊕⊕⊕○<br>Moderate* |

\*Rated down one level for inconsistency due to the high heterogeneity.
†Rated down one level for risk of bias due to the substantial attrition rates.
‡Rated down one level for imprecision due to the wide 95% confidence interval.

**Table 2.**
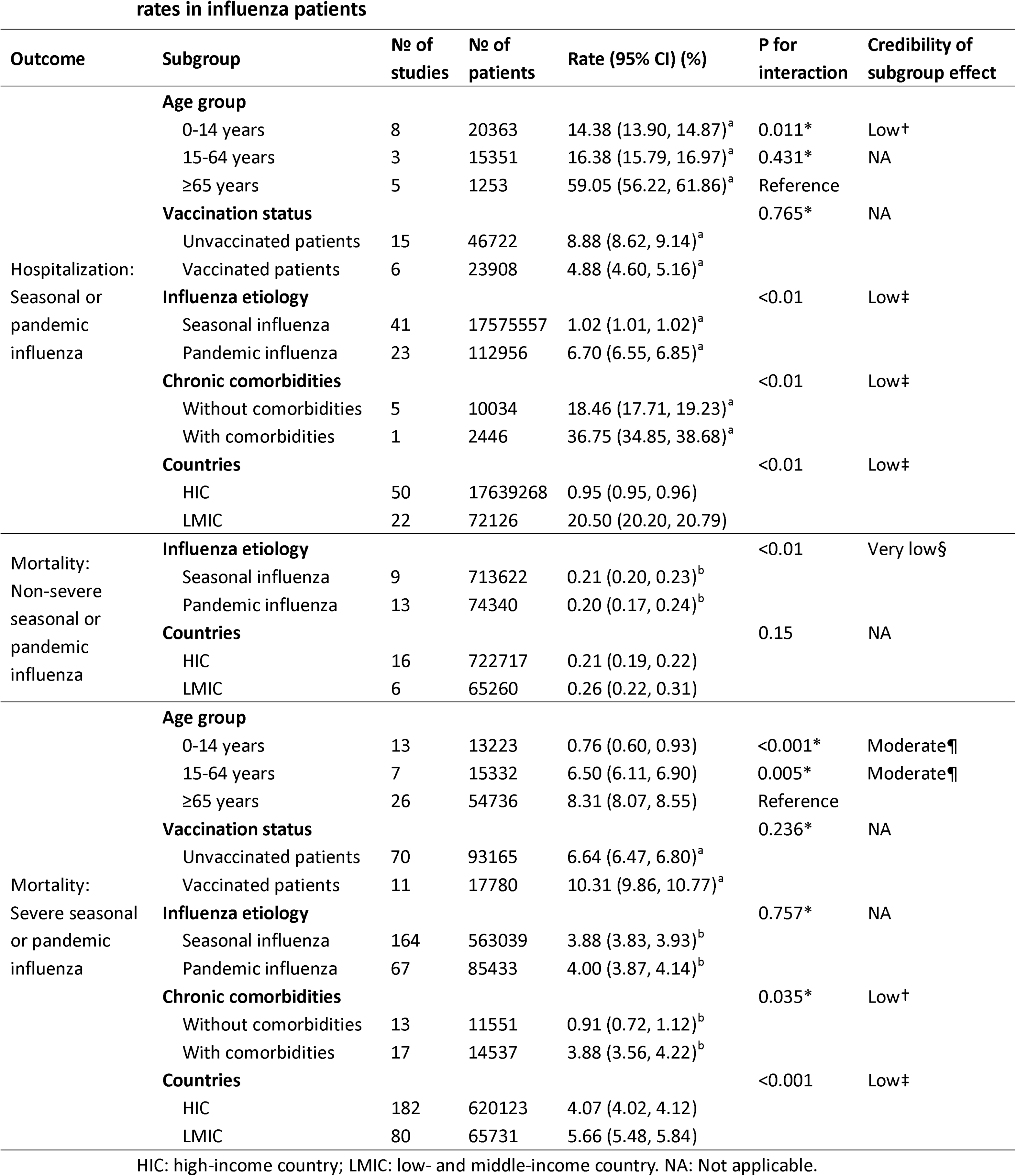
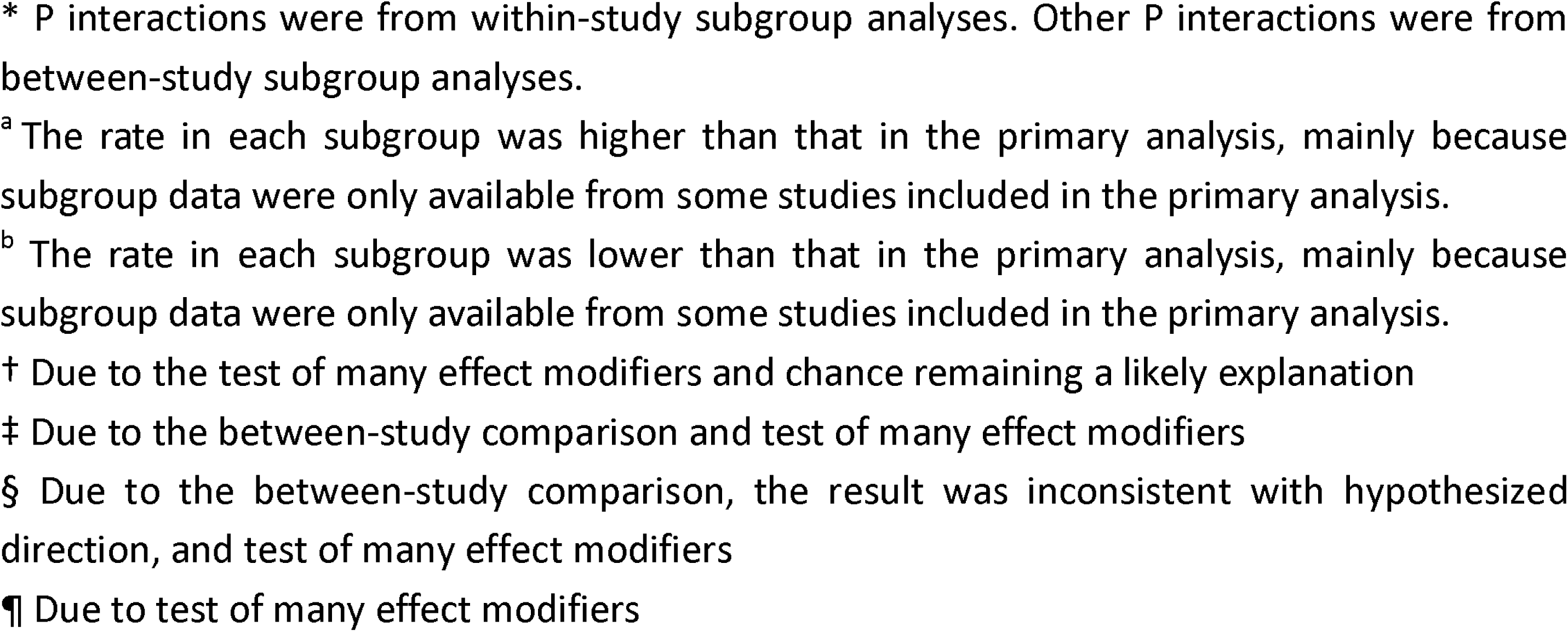
Results of subgroup analyses for hospitalization and all-cause mortality rates in influenza patients.

#### Zoonotic influenza

Low certainty evidence revealed that the hospitalization rate of patients with zoonotic influenza was 93.72% (95%CI 90.12% to 96.58 %; 1 study, 223 patients) (Table 1 and Appendix 13).

### All-cause mortality

The pooled all-cause mortality rate of patients with seasonal or pandemic influenza was 1.75% (95%CI 1.73% to 1.77%; 339 studies, 1,784,166 patients) (Appendix 14).

#### Non-severe seasonal or pandemic influenza

High certainty evidence showed that the all-cause mortality rate of patients with non-severe seasonal or pandemic influenza was 0.23% (95%CI 0.22% to 0.24%; 23 studies, 788,548 patients) (Table 1 and Appendix 15). Between-study subgroup analysis suggested no subgroup effect in all-cause mortality between HICs and LMICs (0.21% versus 0.26%, P for interaction = 0.15, Table 2 and Appendix 16) among patients with non-severe seasonal or pandemic influenza. The analysis revealed comparable all-cause mortality rates between patients with non-severe seasonal influenza and non-severe pandemic influenza (0.21% versus 0.20%), with a statistically significant interaction effect (P < 0.01, Table 2 and Appendix 17). Due to the between-study comparison, the result was inconsistent with hypothesized direction, and test of many effect modifiers, we rated the credibility of this subgroup effect as very low (Appendix 18).

#### Severe seasonal or pandemic influenza

Moderate certainty evidence indicated that the all-cause mortality rate of patients with severe seasonal or pandemic influenza was 4.15% (95%CI 4.10% to 4.20%; 269 studies, 709,672 patients) (Table 1 and Appendix 19). Within-study subgroup analyses showed that patients aged up to 14 years had a lower all-cause mortality rate (0.76% versus 8.31%) than those aged 65 years or older (pooled ratio of rate 0.04, 95% CI: 0.02 to 0.10, P for interaction < 0.001) and patients aged between 15 and 64 years also had a lower all-cause mortality rate (6.50% versus 8.31%) than those aged 65 years or older (pooled ratio of rate 0.49, 95% CI: 0.33 to 0.72, P for interaction = 0.005, Figures 2 and 3). We judged the credibility of these subgroup effects as moderate (Table 2). Within-study subgroup analyses indicated no subgroup effects in all-cause mortality rate between patients with severe seasonal influenza and severe pandemic influenza (P for interaction = 0.76, Table 2 and Appendix 20) and between vaccinated and unvaccinated patients (P for interaction = 0.24, Appendix 21), but a significant difference between patients with chronic comorbidities and those without chronic comorbidities (pooled ratio of rate 2.09, 95% CI: 1.05 to 4.13, P for interaction = 0.04, Appendix 22). However, due to test of many effect modifiers and chance remaining a likely explanation for findings, we rated the credibility of this subgroup effect as low (Appendix 23). Between-study subgroup analysis indicated that patients in HICs had a lower all-cause mortality rate (4.07% versus 5.66%, P for interaction < 0.01, Table 2 and Appendix 24) compared with those in LMICs. However, due to the between-study comparison and test of many effect modifiers, we rated the credibility of this subgroup effect as low (Appendix 23). Sensitivity analysis only including studies with a low risk of bias showed a similar result (4.18%, 95%CI 4.13% to 4.23%) to the primary analysis (Appendix 25).

**Figure 2.**
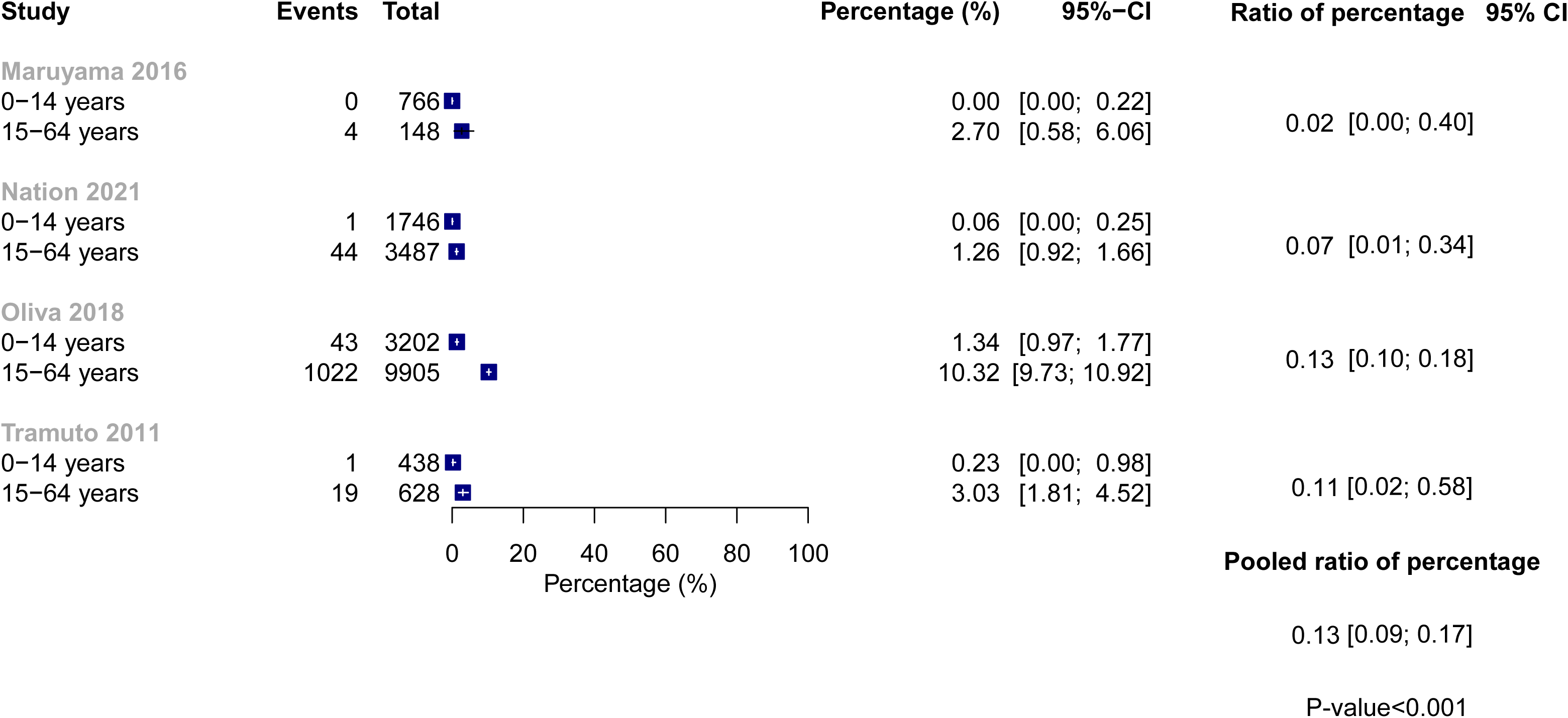
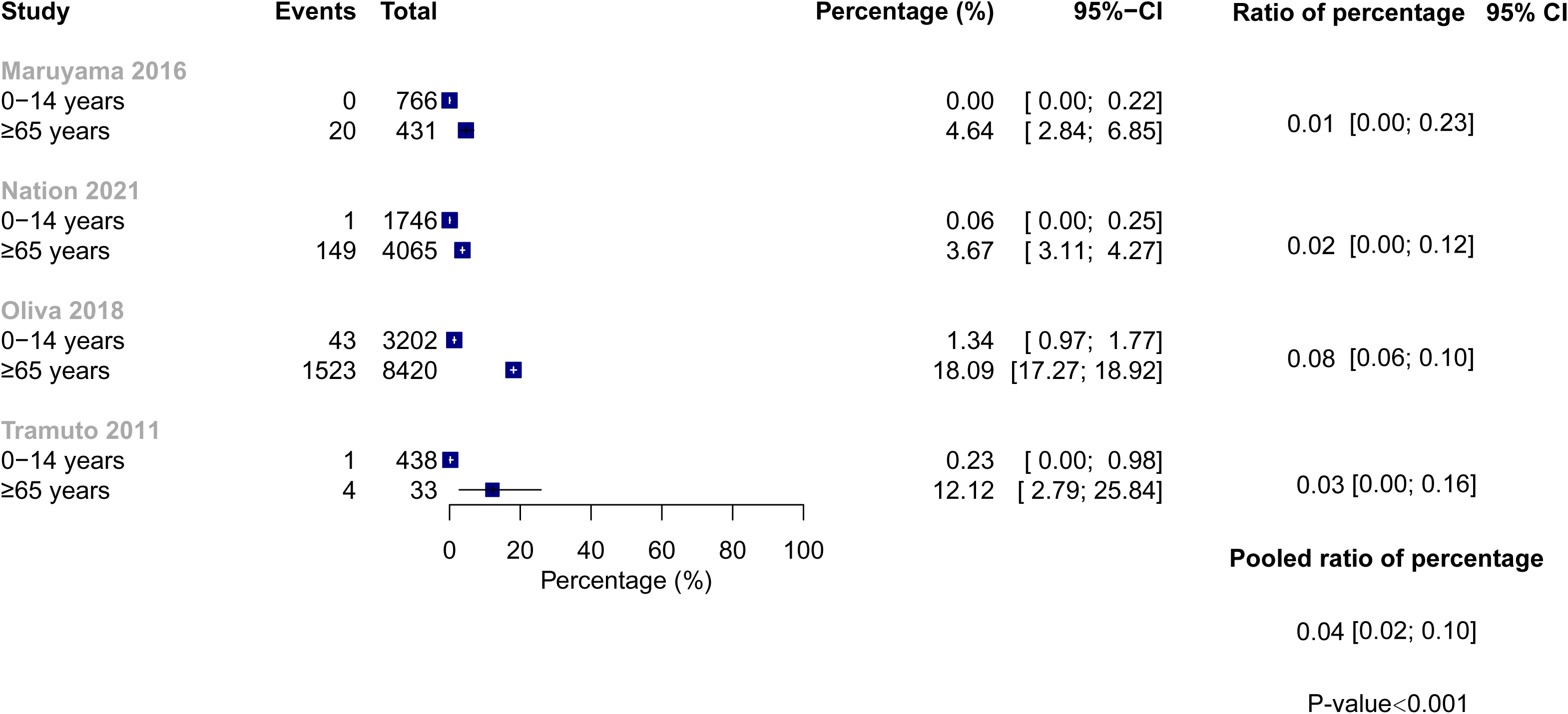
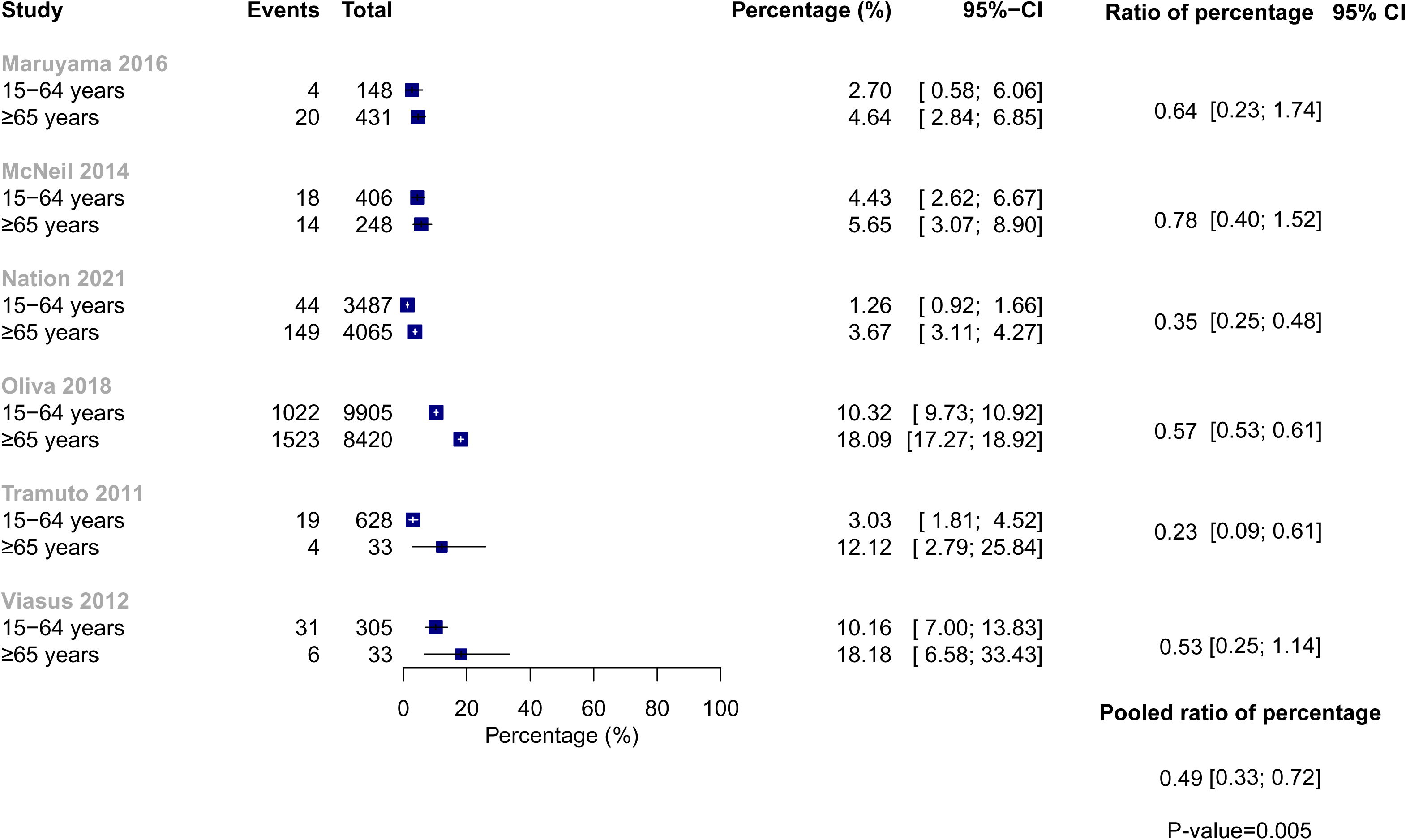
Within-study subgroup analysis of all-cause mortality rate in severe seasonal or pandemic influenza by age (A) 0-14 years vs 15-64 years, (B) 0-14 years vs ≥65 years, and (C) 15-64 years vs ≥65 years

**Figure 3.**
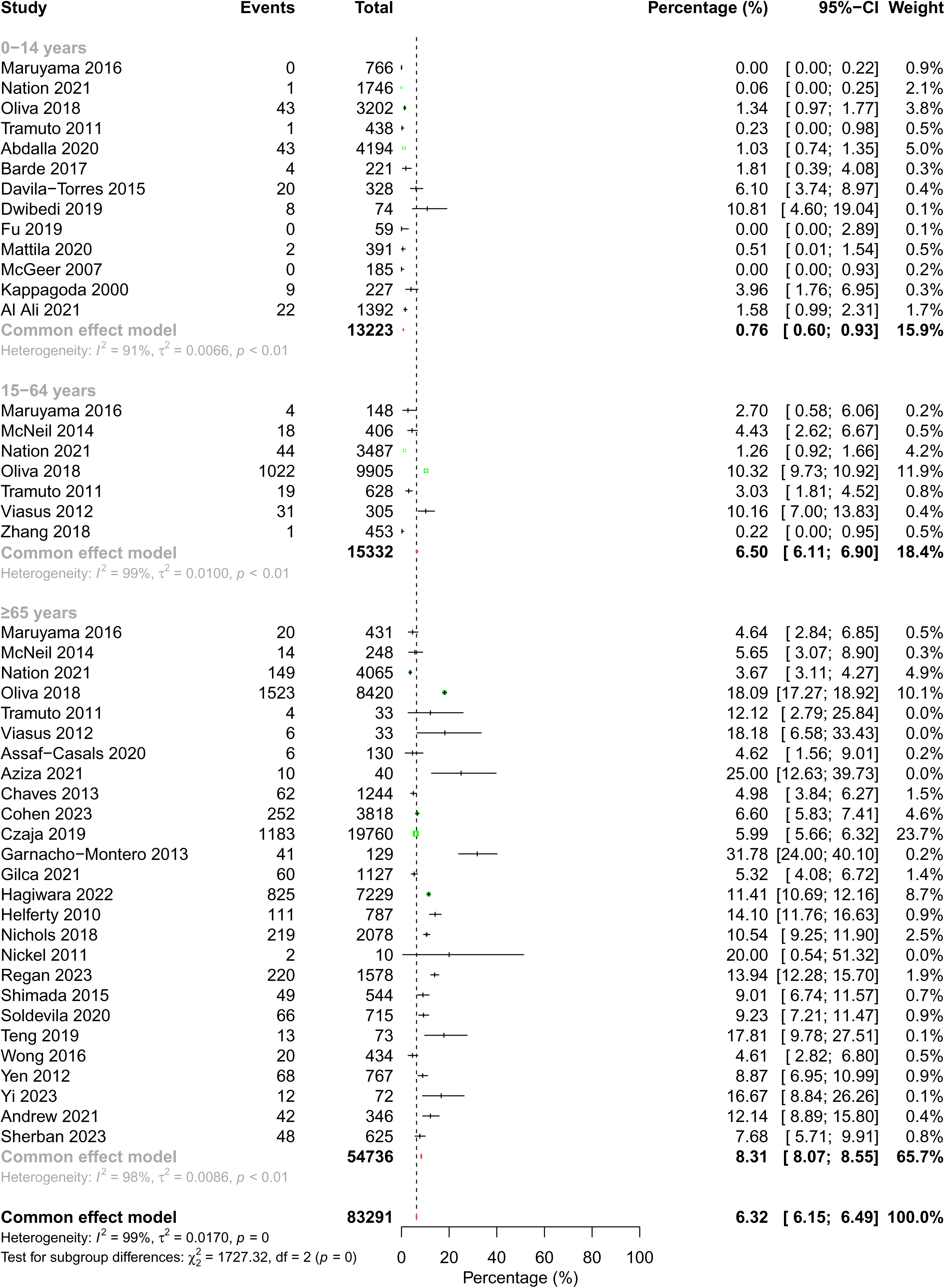
All-cause mortality rate in severe seasonal or pandemic influenza for different age group

#### Severe zoonotic influenza

Moderate certainty evidence showed that the all-cause mortality rate of patients with severe zoonotic influenza was 38.72% (95%CI 36.79% to 40.66%; 9 studies, 2500 patients) (Table 1 and Appendix 26).

## Discussion

This systematic review provided moderate certainty evidence that the hospitalization rate for patients with seasonal or pandemic influenza was 1.01% and low certainty evidence that the hospitalization rate for patients with zoonotic influenza was 93.72%. Moderate to high certainty evidence demonstrated that all-cause mortality rates were 0.23% for patients with non-severe seasonal or pandemic influenza, 4.15% for those with severe seasonal or pandemic influenza, and 38.72% for patients with zoonotic influenza (Table 1). Among severe seasonal or pandemic influenza cases, patients aged ≥65 years (8.31%) had a higher all-cause mortality rate than those aged 0-14 years (0.76%) and 15-64 years (6.50%) that met ICEMAN criteria for a moderate credibility subgroup analysis.

### Strengths and Limitations

The strengths of the review include comprehensive literature searches; independent duplicate study identification, selection, data extraction, and risk of bias assessment; and application of the GRADE approach to rate the certainty of evidence. Based on the WHO guideline development panel’s discussion and suggestions, we summarized hospitalization and all-cause mortality rates separately for seasonal or pandemic influenza and zoonotic influenza, which provide the baseline risk estimates for the updated WHO influenza guideline. To explore the variation of hospitalization and mortality rates across different patient groups, we performed comprehensive subgroup analyses evaluated using the rigorously developed ICEMAN instrument.^22^

Our review has limitations. Although we conducted comprehensive literature searches, the available data for some patient groups, particularly those with zoonotic influenza, were limited. Due to insufficient within-study subgroup information across included studies, we were only able to perform within-trial subgroup analyses for limited subgroups, while relying on between-study subgroup analyses for others. In part as a result, the credibility of identified subgroup effects was generally low even for subgroups with interaction p-values less the 0.05. Furthermore, in some subgroup analyses, the hospitalization or mortality rates for all subgroups were either consistently higher or lower than the results of the overall population. This discrepancy was primarily due to the fact that not all eligible studies reported subgroup data. For studies that did report subgroup data, the overall proportion of each outcome aligned within the middle of the reported subgroup estimates.

To minimize data overlap in included studies, we implemented strict inclusion criteria, focusing specifically on cohort studies and those using surveillance data. For studies derived from the same population source, we prioritized the dataset offering the most comprehensive information based on predefined criteria including sample size and completeness of outcome and subgroup information reporting. To identify potential overlapping populations, we also cross-checked author affiliations, study locations, and recruitment periods. However, despite these rigorous selection methods and careful data extraction procedures, the possibility of residual data duplication cannot be entirely excluded.

The studies that reported all-cause mortality often lacked clear differentiation between in-hospital mortality and post-discharge deaths at specific time points (e.g., 28 or 30-day, 60-day, 90-day mortality). This uncertainty in mortality reporting precluded separate analyses of mortality outcomes at different time points. We therefore pooled these data and estimated only the overall all-cause mortality.

There proved substantial heterogeneity across studies in most meta-analyses, although we have conducted many subgroup analyses to explore potential sources of the heterogeneity. The differences in study populations, influenza virus strains, geographical regions, and time periods may contribute to the heterogeneity. Clinicians and researchers should carefully consider these sources of heterogeneity when interpreting our findings and assess their applicability to specific settings.

### Comparison to other studies

A meta-analysis of 63 datasets reported that influenza was associated with 14.1% (95% CI 12.1% to 16.5%) of acute respiratory hospitalizations among adults worldwide.^12^ A meta-analysis reported a pooled influenza-associated all-cause mortality rate of 14.33 (95% CI 11.56% to 17.10%) per 100,000 persons in China.^13^ Another meta-analysis reported that, in Italy, hospitalization occurred in 0.43% of patients visited by general practitioners for clinical influenza.^9^

Our findings showed a hospitalization rate of 1.01% and an all-cause mortality rate of 1.75% for patients with seasonal or pandemic influenza. To our knowledge, our study presents the most comprehensive review of hospitalization and mortality rates among influenza patients. Compared to previous reviews, our meta-analysis included latest evidence and a large number of participants, assessed the certainty of evidence using the GRADE approach, and provided separate estimates for non-severe seasonal or pandemic influenza, severe seasonal or pandemic influenza, and zoonotic influenza.

### Remaining uncertainties

Our study identifies evidence gaps. Considerable uncertainty remains in hospitalization and all-cause mortality rates among specific populations, particularly children, vaccinated individuals, and patients with various chronic comorbidities. Future studies could prioritize testing of important subgroup analyses, such as influenza etiology (i.e., seasonal, pandemic, or zoonotic influenza), age groups, vaccination status, and comorbidities. The heterogeneity in reported rates underscores the need for more rigorous research with standardized methods and clear definitions of mortality outcomes.

### Conclusions

This systematic review identified a hospitalization rate of 1.01% for seasonal or pandemic influenza and all-cause mortality rates of 0.23% for non-severe seasonal or pandemic influenza, 4.15% for severe seasonal or pandemic influenza, and 38.72% for zoonotic influenza.

## Data Availability

All data produced in the present work are contained in the manuscript.

## Contributors

GG, QH, and YG conceived and designed the study. YG and QH designed and performed the search strategy. YG, ML, KN, YC, YZ, YS, WT, JX, QZ, WZ, XG, LiaZ, LC, JS, LiZ, and MK screened and selected the articles. YG, ML, KN, YC, YZ, YS, ZL, WT, JX, QZ, WZ, XG, LiaZ, LC, and LH extracted the data and assessed the risk of bias. YG and QH analyzed the data. LY provided methodological advice on data analyses. GG and JT supervised the data analyses. YG and QH rated the certainty of evidence. GG provided methodological advice on GRADE assessment. YG, GG, and QH interpreted the data. YG and QH drafted the manuscript. YG, JT, FX, GG, and QH revised the manuscript. All authors approved the final version of the manuscript. YG and QH accessed and verified the underlying data. All authors had full access to all the data in the study and had final responsibility for the decision to submit for publication.

## Data sharing statement

Data in this systematic review with meta-analysis are extracted from published studies available on the internet. All processed data are presented in this article and the appendix.

## Acknowledgements

We thank Timothy M Uyeki (Influenza Division, US Centers for Disease Control and Prevention, Atlanta, Georgia) for help with providing clinical advice and interpreting the results. We thank members of the WHO for critical feedback on the review question, subgroup and outcome selection, and GRADE judgments: Janet Diaz (World Health Organization, Geneva, Switzerland); Steven Mcgloughlin (World Health Organization, Geneva, Switzerland); Jamie Rylance (World Health Organization, Geneva, Switzerland). We thank Rachel Couban (librarian at McMaster University) for helping with developing the search strategy. This work was supported by the WHO.

