## Appendix for "Global and national influenza-associated hospitalization and mortality rates: a systematic review and meta-analysis"

### Appendix 1. Search strategy for databases

**Ovid MEDLINE(R) ALL**

1	exp Influenza, Human/

2	exp Influenza A virus/

3	exp Influenza B virus/

4	exp Influenzavirus C/

5	(Influenza or flu or H1N1 or PH1N1 or H3N2 or AH1N1 or AH3N2 or H5N1 or H7N9).mp. [mp=title, book title, abstract, original title, name of substance word, subject heading word, floating sub-heading word, keyword heading word, organism supplementary concept word, protocol supplementary concept word, rare disease supplementary concept word, unique identifier, synonyms, population supplementary concept word, anatomy supplementary concept word]

6	1 or 2 or 3 or 4 or 5

7	exp Hospitalization/

8	exp Mortality/

9	exp Death/

10	(hospital* or ((inpatient or outpatient) and (admission or care)) or "length of Stay" or ICU or "intensive care unit" or "intensive-care unit" or mortalit* or death* or fatalit*).mp. [mp=title, book title, abstract, original title, name of substance word, subject heading word, floating sub-heading word, keyword heading word, organism supplementary concept word, protocol supplementary concept word, rare disease supplementary concept word, unique identifier, synonyms, population supplementary concept word, anatomy supplementary concept word]

11	7 or 8 or 9 or 10

12	exp Case-Control Studies/

13	(case* adj control*).tw.

14	exp cohort studies/

15	(cohort adj (study or studies)).tw.

16	cohort analy*.tw.

17	(follow up adj (study or studies)).tw.

18	exp Observational Study/

19	(observational adj (study or studies)).tw.

20	exp Longitudinal Studies/

21	longitudinal.tw.

22	exp Retrospective Studies/

23	retrospective.tw.

24	exp Prospective Studies/

25	prospective.tw.

26	exp Cross-Sectional Studies/

27	cross sectional.tw.

28	exp Epidemiologic Studies/

29	(epidemiologic* adj (study or studies)).tw.

30	(case* and series).tw.

31	((population or population-based) adj (study or studies or analys#s)).tw.

32	exp Population Surveillance/

33	Surveillance.tw.

34	12 or 13 or 14 or 15 or 16 or 17 or 18 or 19 or 20 or 21 or 22 or 23 or 24 or 25 or 26 or 27 or 28 or 29 or 30 or 31 or 32 or 33

35	6 and 11 and 34

36	exp animals/ not humans.sh.

37	35 not 36

**Ovid Embase**

1	exp Influenza/ or Influenza virus/

2	exp Influenza A virus/ or exp Influenza A virus/

3	exp Influenza B/ or exp Influenza B virus/

4	exp Influenza C/ or exp Influenza C virus/

5	(Influenza or flu or H1N1 or PH1N1 or H3N2 or AH1N1 or AH3N2 or H5N1 or H7N9).mp.

6	or/1-5

7	exp hospitalization/

8	exp Mortality/

9	exp Death/

10	(hospital* or ((inpatient or outpatient) and (admission or care)) or "length of Stay" or ICU or "intensive care unit" or "intensive-care unit" or mortalit* or death* or fatalit*).mp.

11	or/7-10

12	exp case control study/

13	(case* adj control*).tw.

14	cohort analysis/

15	(cohort adj (study or studies)).tw.

16	cohort analy*.tw.

17	(follow up adj (study or studies)).tw.

18	exp Observational Study/

19	(observational adj (study or studies)).tw.

20	exp longitudinal study/

21	longitudinal.tw.

22	exp retrospective study/

23	retrospective.tw.

24	exp prospective study/

25	prospective.tw.

26	exp cross-sectional study/

27	cross sectional.tw.

28	(epidemiologic* adj (study or studies)).tw.

29	(case* and series).tw.

30	((population or population-based) adj (study or studies or analys#s)).tw.

31	exp population surveillance/

32	or/12-31

33	6 and 11 and 32

34	exp animal experimentation/ or exp models animal/ or exp animal experiment/ or nonhuman/ or exp vertebrate/

35	exp humans/ or exp human experimentation/ or exp human experiment/	25681958

36	34 not 35

37	33 not 36

**Cochrane Central Register of Controlled Trials**

1	exp Influenza, Human/

2	exp Influenza A virus/

3	exp Influenza B virus/

4	exp Influenzavirus C/

5	(Influenza or flu or H1N1 or PH1N1 or H3N2 or AH1N1 or AH3N2 or H5N1 or H7N9).mp.

6	or/1-5

7	exp Hospitalization/

8	exp Mortality/

9	exp Death/

10	(hospital* or ((inpatient or outpatient) and (admission or care)) or "length of Stay" or ICU or "intensive care unit" or "intensive-care unit" or mortalit* or death* or fatalit*).mp.

11	or/7-10

12	6 and 11

**Global Health**

1	exp Influenza/ or Influenza virus/

2	exp Influenza A virus/ or exp Influenza A virus/

3	exp Influenza B/ or exp Influenza B virus/

4	exp Influenza C/ or exp Influenza C virus/

5	(Influenza or flu or H1N1 or PH1N1 or H3N2 or AH1N1 or AH3N2 or H5N1 or H7N9).mp.

6	or/1-5

7	(hospitals or hospital admission or hospital stay).sh.

8	exp Mortality/

9	exp Death/

10	(hospital* or ((inpatient or outpatient) and (admission or care)) or "length of Stay" or ICU or "intensive care unit" or "intensive-care unit" or mortalit* or death* or fatalit*).mp.

11	or/7-10

12	exp Case-Control Studies/

13	(case* adj control*).tw.

14	exp cohort studies/

15	(cohort adj (study or studies)).tw.

16	cohort analy*.tw.

17	(follow up adj (study or studies)).tw.

18	exp observational studies/

19	(observational adj (study or studies)).tw.

20	exp Longitudinal Studies/

21	longitudinal.tw.

22	exp Retrospective Studies/

23	retrospective.tw.

24	prospective.tw.

25	exp Cross-Sectional Studies/

26	cross sectional.tw.

27	(epidemiologic* adj (study or studies)).tw.

28	(case* and series).tw.

29	((population or population-based) adj (study or studies or analys#s)).tw.

30	surveillance.sh.

31	or/12-30

32	6 and 11 and 31

**CINAHL**

| **#** | **Query** |
| --- | --- |
| S33 | S26 NOT S32 |
| S32 | S30 NOT S31 |
| S31 | MH (human) |
| S30 | S27 OR S28 OR S29 |
| S29 | TI (animal model*) |
| S28 | MH (animal studies) |
| S27 | MH animals+ |
| S26 | S3 AND S6 AND S25 |
| S25 | S7 OR S8 OR S9 OR S10 OR S11 OR S12 OR S13 OR S14 OR S15 OR S16 OR S17 OR S18 OR S19 OR S20 OR S21 OR S22 OR S23 OR S24 |
| S24 | (MH "Population Surveillance+") |
| S23 | TI ((population or population-based) W0 (study or studies or analys#s)) OR AB ((population or population-based) W0 (study or studies or analys#s)) |
| S22 | TI case* W0 series OR AB case* W0 series |
| S21 | TI (epidemiologic* W0 (study or studies)) OR AB (epidemiologic* W0 (study or studies)) |
| S20 | (MH "Epidemiological Research+") |
| S19 | TI cross sectional OR AB cross sectional |
| S18 | (MH "Cross Sectional Studies") |
| S17 | TI prospective OR AB prospective |
| S16 | (MH "Prospective Studies+") |
| S15 | TI retrospective OR AB retrospective |
| S14 | (MH "Retrospective Panel Studies") |
| S13 | TI longitudinal OR AB longitudinal |
| S12 | TI (observational W0 (study or studies)) OR AB (observational W0 (study or studies)) |
| S11 | TI (follow up W5 (study or studies) ) OR AB (follow up W5 (study or studies)) |
| S10 | TI cohort analy* OR AB cohort analy* |
| S9 | TI (cohort W0 (study or studies)) OR AB (cohort W0 (study or studies)) |
| S8 | TI case* W0 control* OR AB case* W0 control* |
| S7 | (MH "Case Control Studies+") |
| S6 | S4 OR S5 |
| S5 | TI (hospital* or ((inpatient or outpatient) and (admission or care)) or "length of Stay" or ICU or "intensive care unit" or "intensive-care unit" or mortalit* or death* or fatalit*) OR AB (hospital* or ((inpatient or outpatient) and (admission or care)) or "length of Stay" or ICU or "intensive care unit" or "intensive-care unit" or mortalit* or death* or fatalit*) |
| S4 | (MH "Hospitalization+") OR (MH "Mortality+") OR (MH "Death+") |
| S3 | S1 OR S2 |
| S2 | TI (Influenza or flu or H1N1 or PH1N1 or H3N2 or AH1N1 or AH3N2 or H5N1 or H7N9) OR AB (Influenza or flu or H1N1 or PH1N1 or H3N2 or AH1N1 or AH3N2 or H5N1 or H7N9) |
| S1 | (MH "Influenza+") OR (MH "Influenza A Virus+") OR (MH "Influenzavirus C") OR (MH "Influenza B Virus") |

### Appendix 2. References of included studies

### Appendix 3. Basic characteristics of eligible studies

| **Study** | **Country** | **Study design** | **Center 1=multi-center 2=single center** | **Recruitment time period** | **Sample size** | **Male (%)** | **Age, mean or median (years)** | **Age range (years)** | **Severe patients (%)** | **Influenza vaccination (%)** | **Type of influenza (%)** |
| --- | --- | --- | --- | --- | --- | --- | --- | --- | --- | --- | --- |
| Abdalla 2020 | Saudi Arabia | Retrospective | 1 | 2010-2016 | 17094 | NR | NR | NR | 100 | NR | A(H1N1)pdm09 (100) |
| Guesneau 2021 | France | Retrospective | 2 | 2015.1-2015.4 | 114 | 28.1 | 87.9 | ≥75 | 100 | 38.6 | A (93.9), B (6.1) |
| Ackerson 2019 | USA | Retrospective | 1 | 2011.1.1-2015.6.30 | 1878 | 49.7 | 77.4 | 60-104 | 100 | 73.1 | NR |
| Adams 2022 | USA | Retrospective | 1 | 2021.10.1-2022.4.30 | 575 | 56.9 | 6.0 | 0-17 | 100 | 37.2 | A (97.4), B (1.9), A and B (0.7) |
| Adisasmito 2010 | Indonesia | Retrospective | 1 | 2005.5-2009.8 | 93 | 45.0 | 18.0 | 1-67 | 100 | NR | A(H5N1) (100) |
| Adlhoch 2023 | Austria, Belgium, Czech Republic, Ireland, Malta, Netherlands, Portugal, Romania, Slovakia, Spain, Sweden | Retrospective | 1 | 2010.11-2019.12 | 19937 | 53.5 | 60.0 | NR | 100 | 32.2 | A (80.7), B (19.3) |
| Adlhoch 2019 | Austria, Czech Republic, Spain, Finland, France, Ireland, Malta, Netherlands, Portugal, Romania, Sweden, Slovakia | Retrospective | 1 | 2009-2017 | 13368 | 56.9 | 59.0 | 0-104 | 100 | NR | A(H1N1)pdm09 (39.1), A(H3N2) (14.8), A unsubtyped (34.5), B (11.7) |
| Adlhoch 2018 | Czech Republic, Denmark, Finland, France, Ireland, Netherlands, Romania, Spain, Sweden, UK | Prospective | 1 | 2016-2018 | 32472 | NR | NR | NR | 100 | NR | NR |
| Aguirre 2011 | Canada | Retrospective | 2 | 2009.4.16-2009.7.31, 2006.6-2009.3 | 237 | 51.1 | 7.7 | 0.08-17 | 0.0 | NR | A (100) |
| Ahout 2018 | Netherlands | Retrospective | 1 | 2009.8-2010.8 | 940 | 56.0 | 3.0 | 0-18 | 100 | NR | A(H1N1)pdm09 (100) |
| Akinci 2011 | Turkey | Prospective | 2 | 2009.10-2009.12 | 113 | 35.4 | 35.7 | 17-77 | 100 | 0 | A(H1N1)pdm09 (100) |
| Al Ali 2021 | United Arab Emirates | Retrospective | 2 | 2012-2017 | 1392 | 57.0 | NR | 0-15 | 0.3 | 10.2 | A (82.2), B (17.8) |
| Al Subaie 2012 | Saudi Arabia | Prospective | 2 | 2009.7-2009.12 | 375 | 60.0 | 3.0 | 0.08-12 | 13.3 | NR | A(H1N1)pdm09 (100) |
| Al-Abdallat 2016 | Jordan | Retrospective | 1 | 2008.1-2014.2 | 257 | 60.0 | 3.0 | 0-85 | 100 | NR | A(H3N2) (35.0), A(H1N1)pdm09 (34.2), Seasonal A(H1N1) (5.4), B (25.3) |
| Al-Awaidy 2015 | Oman | Prospective | 1 | 2008.1-2013.6 | 423 | 48.0 | 6.0 | 0-87 | 100 | NR | A (64.0), B (36.0) |
| Al-Baadani 2019 | Saudi Arabia | Retrospective | 2 | 2012.1-2015.12 | 366 | 49.7 | 54.4 | NR | 100 | NR | A (41.0), B (59.0) |
| Al-Busaidi 2016 | Oman | Retrospective | 2 | 2009.8-2009.12 | 1388 | 39.7 | 23.0 | 0.07-67 | 4.9 | NR | A(H1N1) (100) |
| Allam 2013 | India | Retrospective | 1 | 2009.5-2010.12 | 1480 | 52.4 | NR | NR | NR | NR | A(H1N1) (100) |
| Al-Lawati 2010 | Oman | Retrospective | 2 | 2009.7.21-2009.12.23 | 131 | 36.6 | 25.9 | 0.08-78 | 100 | NR | A(H1N1) (100) |
| Alvarez-Lerma 2017 | Spain | Retrospective | 1 | 2009.1.1-2015.12.31 | 1327 | 57.9 | 49.7 | NR | 100 | 6.02 | A(H1N1)pdm09 (100) |
| Amaravathi 2015 | India | Retrospective | 2 | 2014.12.10-2015.5.11 | 88 | 51.1 | 31.2 | NR | 100 | NR | A(H1N1) (100) |
| Ampofo 2006 | USA | Retrospective | 2 | 2001.7-2014.6 | 325 | NR | NR | <18 | 100 | NR | A (87.6), B (12.3), A and B (0.1) |
| Andres 2019 | Spain | Retrospective | 2 | 2012.10-2016.5 | 2684 | 51.0 | 35.0 | 4-62 | 55.7 | NR | A (66.1), B (33.9) |
| Andrew 2021 | Canada | Prospective | 1 | 2011-2012 | 346 | 45.1 | 80.6 | ≥65 | 100 | 59.8 | NR |
| Andrew 2023 | Argentina, Brazil, Canada, China, Colombia, France, India, Ivory Coast, Kenya, Lebanon, Mexico, Peru, Romania, Serbia, South Africa, Spain, Russia | Prospective | 1 | 2018.11-2019.10 | 3512 | 50.4 | NR | >5 | 100 | 9.2 | A(H1N1)pdm09 (49.2), A(H3N2) (38.0), A unsubtyped (10.7), B Yamagata (2.1), B Victoria (1.2), B unsubtyped (1.4) |
| Angeles-Sistac 2020 | Mexico | Retrospective | 2 | 2009.4-2017.3 | 188 | 47.3 | 47.0 | 15-79 | 60.6 | 9.6 | A(H1N1)pdm09 (54.2), A(H3N2) (26.0), B (12.2) |
| CDC 2009 | USA | Retrospective | 1 | 2009.4-2009.5 | 1557 | 43.0 | 12.0 | 0.07-91 | 13.2 | NR | A(H1N1)pdm09 (100) |
| CDC 2010 | Greece | Retrospective | 1 | 2009.5-2010.2 | 18075 | NR | NR | NR | 1.6 | NR | A(H1N1)pdm09 (100) |
| CDESS 2022 | Australia | Retrospective | 1 | 2011-2018 | 669370 | 45.6 | NR | NR | 2.0 | NR | A (55.7), B (32.1), C (<0.1) |
| Ao 2019 | Guatemala | Retrospective | 1 | 2008.5-2012.7 | 446 | 57.0 | 18.3 | 0.02-83.6 | 100 | NR | A (81.0), B (18.0), A and B (1.0) |
| Assaf-Casals 2020 | Lebanon | Retrospective | 2 | 2008.1-2016.6 | 1829 | 49.9 | NR | NR | 26.6 | 22.8 | A (83.7), B (13.3), A and B (3.0) |
| Auvinen 2022 | Finland | Prospective | 2 | 2018.11-2019.4 | 106 | 52.0 | 67.0 | 18-92 | 100 | 53.8 | A (98.1), B (1.9) |
| Aziza 2021 | Canada | Retrospective | 2 | 2014-2019 | 130 | 55.0 | 56.0 | ≥18 | 100 | 9 | A (83.0), B (16.0), A and B (1.0) |
| Babamahmoodi 2017 | Iran | Retrospective | 1 | 2015.3-2016.3 | 428 | NR | 46.9 | 14-95 | 100 | NR | A(H1N1) (100) |
| Baggett 2012 | Thailand | Prospective | 1 | 2009.1-2010.12 | 902 | NR | 24.4 | 0-95 | 100 | NR | A (86.4), B (13.3), A and B (0.3) |
| Baigalmaa 2012 | Mongolia | Retrospective | 1 | 2009.12-2010.1 | 1322 | NR | NR | NR | 100 | NR | A(H1N1)pdm09 (100) |
| Baker 2017 | USA | Retrospective | 1 | 2013.8-2014.4 | 431 | NR | NR | NR | 100 | NR | NR |
| Barakat 2012 | Morroco | Prospective | 1 | 2009.6-2010.2 | 1398 | 49.3 | NR | NR | 100 | NR | A(H1N1)pdm09 (100) |
| Barde 2017 | India | Prospective | 1 | 2015.1-2015.5 | 1607 | 47.5 | 35.2 | 0.08-90 | 100 | NR | A(H1N1)pdm09 (100) |
| Baselga-Moreno 2019 | Russia, Kazakhstan, Czech Republic, Canada, Romania, Turkey, Spain, Tunisia, China, India, Mexico, Ivory Coast, Peru, South Africa | Prospective | 1 | 2016.10-2017.5 | 2895 | 46.3 | 64.0 | 18-76 | 100 | NR | A (70.6), B (29.4) |
| Bassetti 2019 | Italy | Retrospective | 2 | 2017.10-2018.4 | 29 | 51.7 | 48.3 | 0-87 | 100 | 31.0 | A (34.5), B (65.5) |
| Begley 2022 | USA | Prospective | 1 | 2016.9-2019.5 | 1940 | 44.0 | NR | NR | 100 | 50.6 | NR |
| Belchior 2011 | Algeria | Prospective | 1 | 2009.10-2013.4 | 1234 | NR | NR | NR | 3.7 | 1.7 | A(H1N1) (27.9), A(H1N1pdm09) (6.0), A(H3N2) (25.3), B (38.1), A and B (0.3) |
| Bergmann 2023 | Austria | Retrospective | 2 | 2015.1-2022.4 | 39 | 53.8 | 58.6 | NR | 100 | NR | A (86.0), B (15.4) |
| Biasco 2022 | Switzerland | Prospective | 1 | 2018.12-2019.3 | 145 | 44.1 | 76.0 | ≥18 | 100 | NR | A (99.3), A and B (0.7) |
| Bilgin 2023 | Turkey | Retrospective | 2 | 2017.10-2020.3 | 201 | 47.3 | 62.0 | >17 | 100 | NR | A (72.6), B (26.9), unknown (0.5) |
| Birkelo 2021 | USA | Retrospective | 1 | 2019.10-2020.5 | 3680 | 92.8 | 66.5 | NR | 100 | NR | NR |
| Chao 2018 | China | Retrospective | 1 | 2015.10-2016.3 | 296 | 62.8 | 61.4 | NR | 100 | NR | A (75.7), B (8.1), unknown (16.2) |
| Chen 2020 | China | Retrospective | 1 | 2013.1-2019.5 | 1079 | 54.1 | 61.0 | NR | 100 | NR | A (64.2), B (35.8) |
| Chien 2022 | China | Retrospective | 1 | 2010-2016 | 6193 | 53.8 | 5.2 | ≤18 | 31.7 | NR | A (65.7), B (32.2), A and B (2.1) |
| Choi 2011 | South Korea | Retrospective | 1 | 2009.5-2010.2 | 269 | 53.2 | 48.0 | NR | 98.9 | NR | A(H1N1) (100) |
| Choi 2017 | South Korea | Prospective | 1 | 2013.9-2014.5 | 3341 | 39.4 | 47.5 | >20 | 23.8 | 7.6 | A (71.8), B (27.6), A and B (0.5) |
| Chorazka 2021 | Switzerland | Retrospective | 1 | 2017-2019 | 469 | 42.6 | 74.0 | ≥18 | 100 | NR | NR |
| Chowell 2012 (a) | USA | Retrospective | 1 | 2019.4-2020.3 | 609 | 44.5 | 21.0 | 0-96 | 87.4 | NR | A(H1N1)pdm09 (100) |
| Chowell 2012 (b) | Mexico | Prospective | 1 | 2009.8-2009.12 | 2944 | NR | NR | ≥18 | 100 | 7.3 | A(H1N1)pdm09 (100) |
| Chu 2023 | China | Retrospective | 1 | 2015.1-2020.1 | 373 | 60.0 | 63.0 | NR | 100 | 0 | A (100) |
| Chuang 2012 | China | Retrospective | 1 | 2009.7-2011.3 | 6737 | NR | NR | NR | 45.2 | NR | A(H1N1)pdm09 (79.7), B (13.7), A(H3N2) (5.4), seasonal A(H1N1) (0.3) |
| Chuaychoo 2021 | Thailand | Retrospective | 2 | 2014.1-2017.12 | 421 | NR | 66.0 | ≥18 | 100 | 11.4 | A(H1N1) (22.1), A(H3N2) (51.5), B (26.4) |
| Ciftci 2011 | Turkey | Retrospective | 1 | 2009.7-2010.2 | 821 | 57.4 | 3.0 | ≤18 | 100 | 4 | A(H1N1)pdm09 (100) |
| Cobb 2021 | USA | Retrospective | 1 | 2019.1.1-2020.4.15 | 74 | 56.8 | 56.8 | 20-92 | 100 | NR | A (78.4), B (21.6) |
| Cohen 2014 | South Africa | Prospective | 1 | 2009.2-2012.12 | 1239 | NR | NR | NR | 100 | NR | A(H3N2) (37.4), A(H1N1)pdm09 (27.3), A subtype unknown (2.6), B (33.7) |
| Cohen 2023 | Argentina, Brazil, Canada, China, Colombia, France, India, Ivory Coast, Kenya, Lebanon, Mexico, Peru, Romania, Serbia, South Africa, Spain, Russia | Prospective | 1 | 2012-2019 | 15560 | 49.6 | NR | NR | 100 | 51.6 | A(H3N2) (35.3), A(H1N1)pdm09 (29.3), A subtype not determined (7.9), B Victoria (8.1), B Yamagata (14.2), B lineage not determined (5.2) |
| Collins 2019 | USA | Retrospective | 1 | 2011-2015 | 5262 | 56.4 | NR | NR | 100 | 32.7 | A (73.3), B (25.3), A and B (0.6), unknown (0.8) |
| Cordero 2012 | Spain | Prospective | 1 | 2009.6-2010.1 | 51 | 54.9 | 48.0 | 16-73 | 100 | 1.9 | A(H1N1) (100) |
| Correia 2010 | Portugal | Prospective | 1 | 2009.6-2010.2 | 5690 | NR | NR | NR | NR | NR | A(H1N1) (100) |
| Cost 2011 | USA | Retrospective | 2 | 2009.4.27-2009.12.5 | 30 | 50.0 | 10.0 | ≤21 | 33.3 | NR | A(H1N1) (100) |
| Coussement 2022 | France, Belgium | Retrospective | 1 | 2011.11.1-2018.4.30 | 309 | 53.7 | 65.3 | ≥18 | 100 | NR | NR |
| Cox 2012 | USA | Prospective | 1 | 2009.4-2010.4 | 7717 | 47.0 | NR | NR | 100 | 0.79 | A(H1N1)pdm09 (100) |
| Cui 2010 | China | Retrospective | 2 | 2009.11.4-2009.12.31 | 68 | 73.5 | 41.0 | 18-66 | 100 | NR | A(H1N1) (100) |
| Cummings 2022 | USA | Retrospective | 1 | 2011.1-2019.12 | 3551 | 91.5 | 64.5 | NR | NR | 100 | NR |
| Czaja 2019 | USA | Retrospective | 1 | 2011.10-2012.4, 2014.10-2015.4 | 19760 | 44.8 | NR | ≥65 | 100 | 57.8 | A (100) |
| Da Dalt 2011 | Italy | Retrospective | 1 | 2009.10-2010.1 | 200 | 57.0 | 4.2 | ≤15 | 100 | 2 (H1N1 vaccine), 4 (seasonal influenza vaccine) | A(H1N1) (100) |
| DaeiSorkhabi 2022 | Iran | Retrospective | 1 | 2012.2-2021.2 | 3101 | 47.5 | NR | 0-98 | 100 | 5.32 | A(H3N2) (51.2), A(H1N1) (28.1), B (20.7) |
| Dao 2010 | USA | Retrospective | 1 | 2005.10-2006.4, 2007.10-2008.4 | 5055 | 43.4 | NR | ≥18 | 100 | 33.7 | A (73.6), B (22.8), A and B (0.1), Unknown (12.7) |
| Davila-Torres 2015 | Mexico | Prospective | 1 | 2013.10.1-2014.3.31 | 2241 | NR | NR | NR | 100 | NR | A(H1N1) (100) |
| Dawood 2014 | USA | Prospective | 1 | 2003-2010 | 7293 | NR | 3.5 | <18 | 100 | NR | A(H1N1)pdm09 (34.1), seasonal (65.9) |
| Dawood 2020 | Australia, Canada, Israel, USA | Retrospective | 1 | 2010-2016 | 614 | 0.0 | NR | 18-50 | 100 | 12 | A(H1N1) (28.8), A unsubtyped (33.4), A(H3N2) (20.4), B (18.0) |
| Dawood 2010 | Australia | Prospective | 1 | 2009.6.1-2009.8.30 | 8210 | NR | NR | NR | NR | NR | A(H1N1)pdm09 (100) |
| Dawood 2009 | USA | Retrospective | 1 | 2009.4.15-2009.5.5 | 642 | 51.0 | 20.0 | 0.25-81 | 9.0 | NR | Swine-origin Influenza A(H1N1) (100) |
| de Morais 2022 | Brazil | Retrospective | 1 | 2020-2022 | 2273 | 43.0 | 40.5 | 18-59 | 100 | 11.9 | A (90.5), B (9.5) |
| Deguchi 2001 | Japan | Prospective | 1 | 1998.11-1999.3 | 950 | 43.6 | NR | ≥60 | 19.2 | 27 | A(H3N2) (100) |
| Delahoy 2023 | USA | Retrospective | 1 | 2017.10-2022.4 | 6774 | 56.3 | 3.0 | NR | 27.3 | NR | A(H1N1)pdm09 (100) |
| Delgado-Sanz 2020 | Spain | Retrospective | 1 | 2010.9-2017.5 | 8985 | 55.2 | NR | NR | 100 | 18.2 | A(H1N1)pdm09 (51.0), A(H3N2) (34.0), B (15.0) |
| Diaz 2012 | Spain | Prospective | 1 | 2009-2010 | 372 | 55.1 | 43.4 | NR | 100 | NR | A(H1N1)pdm09 (100) |
| Dickow 2023 | Germany | Retrospective | 1 | 2016.11-2022.8 | 4916 | 50.1 | 60.0 | NR | 100 | NR | NR |
| Dolan 2012 | UK | Retrospective | 1 | 2009.5-2010.1 | 395 | 0.0 | NR | 15-44 | 100 | 7.1 | A(H1N1)pdm09 (100) |
| Dou 2020 | USA | Retrospective | 2 | 2010.12-2014.7 | 433 | 53.8 | 67.8 | NR | 100 | NR | NR |
| Doyle 2013 | USA | Retrospective | 1 | 2009.6-2010.9 | 194 | 0.0 | 25.9 | NR | 100 | NR | A(H1N1)pdm09 (100) |
| Draganescu 2019 | Romania | Retrospective | 1 | 2017.12-2018.4 | 259 | 51.4 | 6.0 | 2.5−36 | 100 | 2.9 | NR |
| Duarte 2009 | Brazil | Retrospective | 1 | 2009.7-2009.8 | 37 | 48.6 | 35.0 | ≥13 | 100 | NR | A(H1N1)pdm09 (100) |
| Dwibedi 2019 | India | Retrospective | 1 | 2009-2017 | 606 | 50.7 | NR | NR | 100 | NR | A(H1N1)pdm09 (100) |
| Dwyer 2017 | Argentina, Australia, Belgium, Canada, Chile, China, Colombia, Denmark, Estonia, Germany, Greece, Japan, Norway, Peru, Poland, Spain, Thailand, UK, USA | Prospective | 1 | 2009.10-2015.9 | 5350 | 47.7 | 43.8 | ≥18 | 26.1 | 22 | A (80.7), B (19.3) |
| Ebrahimi 2022 | Iran | Retrospective | 1 | 2016.1-2018.12 | 9146 | 44.5 | 40.0 | 25-74 | 100 | NR | A (72.8), B (27.2) |
| Echevarria-Zuno 2009 | Mexico | Prospective | 1 | 2009.4-2009.7 | 6945 | 50.0 | 16.0 | NR | 7.8 | NR | A(H1N1) (100) |
| Emborg 2022 | Denmark | Retrospective | 1 | 2021.12-2022.3 | 7691 | 46.7 | NR | ≥2 | 18.0 | 25.7 | A(H3N2) (96.7), A(H1N1)pdm09 (0.7), B (2.6) |
| Esposito 2011 | Italy | Prospective | 1 | 2007-2009 | 729 | 70.8 | 4.6 | <15 | 16.9 | NR | A (67.2), B (32.8) |
| Ezzine 2023 | Morocco | Retrospective | 1 | 2015-2019 | 1323 | 46.7 | NR | ≥2 | 41.7 | 1.5 | A (79.4), B (20.6) |
| Fahim 2021 | Egypt | Prospective | 1 | 2016-2019 | 7164 | 51.4 | NR | NR | 100 | NR | A (66.9), B (33.1) |
| Fahim 2022 | Egypt | Prospective | 1 | 2020.1-2022.4 | 52 | 55.8 | 33.2 | NR | 38.5 | NR | NR |
| Farias 2010 | Argentina | Prospective | 1 | 2009.6-2009.7 | 147 | 57.8 | 21.3 | <18 | 100 | NR | A(H1N1) (100) |
| Feikin 2012 | Kenya | Prospective | 1 | 2007.6-2009.5 | 204 | NR | NR | NR | 100 | NR | A (86.8), B (13.7), A and B (0.5) |
| Feldstein 2021 | USA | Prospective | 1 | 2015.11.6-2016.6.16 | 139 | 32.4 | 2.0 | 0.5-17 | 100 | 38.8 | A(H1N1)pdm09 (42.0), A(H3N2) (14.0), A with unknown subtype (2.0), B (42.0) |
| Fielding 2009 | Australia | Retrospective | 1 | 2009.4-2009.9 | 513 | NR | NR | NR | 100 | NR | A(H1N1) (44.4), Other type A (54.8), Type B (0.2) |
| Frohlich 2022 | Switzerland | Retrospective | 1 | 2018.10-2020.3 | 1381 | 48.2 | 74.0 | >18 | 100 | NR | A (96.4), B (3.6) |
| Fu 2019 | China | Retrospective | 1 | 2011.1-2018.2 | 458 | 59.2 | NR | NR | 100 | 0 | A(H1N1)pdm09 (40.8), A(H3N2) (16.2), B (43.0) |
| Fuhrman 2011 | France | Prospective | 1 | 2009.7-2010.2 | 1065 | 53.0 | 49.0 | ≥15 | 100 | 1.5 | A(H1N1) (97.0), A undetermined subtype (3.0) |
| Fullana Barcelo 2021 | Spain | Retrospective | 2 | 2012.10-2013.5, 2015.10-2016.5 | 666 | 50.8 | NR | >18 | 100 | 38.7 | A(H1N1) (36.3), A(H3N2) (36.2), A no subtype (5.0), B (22.5) |
| Fuller 2022 | USA | Retrospective | 1 | 2017.3-2020.3 | 29520 | 42.3 | 64.4 | ≥18 | 100 | NR | NR |
| Gachari 2022 | Kenya | Prospective | 1 | 2014.1-2018.12 | 716 | 59.2 | NR | NR | 100 | NR | NR |
| Gacouin 2020 | France | Retrospective | 2 | 2009.10-2020.3 | 103 | 53.0 | NR | >18 | 100 | NR | A(H1N1) (46.0), A(H3N2) (44.0), B (10.0) |
| Galindo-Fraga 2013 | Mexico | Prospective | 1 | 2010.4-2011.4 | 150 | NR | NR | >0.25 | 42.0 | 41.3 | A (5.3), H1N1 (4.0), A(H3N2) (48.7), B (42.0) |
| Garnacho-Montero 2013 | Spain | Prospective | 1 | 2009.4-2011.7 | 1120 | 60.5 | 47.2 | ≥15 | 100 | NR | A(H1N1) (100) |
| Garnacho-Montero 2018 | Spain | Prospective | 1 | 2009.1-2015.12 | 1899 | 60.2 | 51.0 | 39-61 | 100 | NR | A(H1N1) (100) |
| Geerdes-Fenge 2022 | Germany | Retrospective | 2 | 2017.12-2018.5 | 272 | 57.0 | 48.4 | 0-94 | 63.6 | 32.4 | A (25.7), B (73.9), A and B (0.4) |
| Gefenaite 2018 | Romania | Retrospective | 1 | 2011-2015 | 496 | NR | NR | NR | 100 | NR | NR |
| Ghosh 2021 | India | Retrospective | 2 | 2009.8-2017.7 | 855 | NR | NR | 0.88-15 | 36.3 | NR | A(H1N1) (100) |
| Giannella 2012 | Spain | Prospective | 2 | 2010.12-2011.2 | 31 | 64.5 | 64.0 | 48-70 | 100 | 22.6 | A(H1N1) (87.1), A(H3N2) (3.2), B (9.7) |
| Gilca 2021 | Canada | Prospective | 1 | 2011-2019 | 1831 | 45.5 | NR | NR | 100 | 33.2 | A(H1N1) (24.0), A(H3N2) (59.0), B (15.0) |
| Gioula 2010 | Greece | Retrospective | 1 | 2009.4-2009.12 | 1439 | 52.0 | 27.0 | 1-81 | 14.6 | 6.6 | A(H1N1) (100) |
| Godoy 2018 | Spain | Retrospective | 1 | 2010-2016 | 1727 | 57.0 | NR | ≥18 | 100 | 26.3 | A (85.9), B (14.1) |
| Golagana 2023 | India | Prospective, retrospective | 1 | 2018.9-2019.1 | 154 | 59.1 | 58.2 | ≥16 | 100 | 0 | A(H1N1) (100) |
| Gouya 2011 | Iran | Retrospective | 1 | 2009.5.22-2009.12.21 | 3672 | NR | NR | NR | NR | NR | A(H1N1) (100) |
| Groeneveld 2020 | Netherlands | Retrospective | 1 | 2013.10.1-2016.4.1 | 176 | 57.9 | 65.0 | ≥18 | 100 | NR | A (80.1), B (19.9) |
| Gubbels 2013 | Denmark | Prospective | 1 | 2009.11-2011.4 | 159 | 56.7 | 50.0 | NR | 100 | NR | A(H1N1)pdm09 (100) |
| Gutierrez-Cuadra 2012 | Spain | Prospective | 1 | 2009.6-2009.12 | 111 | 61.3 | 49.0 | 15-89 | 100 | NR | A(H1N1)pdm09 (100) |
| Gutierrez-Pizarraya 2012 | Spain | Prospective | 2 | 2010.12.1-2011.3.31 | 130 | 30.0 | 38.0 | 26-60 | 64.6 | 30 | A(H1N1) (61.5), B (38.5) |
| Hagerman 2015 | Switzerland | Retrospective | 1 | 2009.6-2010.1 | 326 | NR | 3.6 | 0-18.3 | 93.3 | 6.4 | A(H1N1)pdm09 (100) |
| Hagiwara 2022 | Japan | Retrospective | 1 | 2010.9-2019.9 | 31122 | 47.3 | NR | ≥60 | 24.6 | NR | A (69.8), B (17.8), A and B (9.0) |
| Han 2011 | China | Retrospective | 1 | 2009.10.20-2009.11.30 | 83 | NR | NR | NR | 100 | 0 | A(H1N1)pdm09 (100) |
| Han 2021 | South Korea | Retrospective | 2 | 2014.10-2015.3, 2018.10-2019.3 | 2522 | 53.8 | NR | <20 | 22.4 | NR | A(H3N2) (37.1), A(H1N1)pdm09 (13.4), B/Yamagata (18.9), B/Victoria (17.1) |
| Harris 2019 | USA | Retrospective | 2 | 2017.8-2018.3 | 33 | 39.4 | 71.2 | NR | 100 | NR | A unspecified type (36.4), A(H3N2) (48.5), A(H1N1) (6.1), B (9.1) |
| Hedberg 2022 | Sweden | Retrospective | 2 | 2011.10-2020.9 | 3339 | NR | NR | NR | 100 | NR | NR |
| Helferty 2010 | Canada | Retrospective | 1 | 2009.4-2010.4 | 8678 | 50.0 | NR | NR | 100 | NR | A(H1N1)pdm09 (100) |
| Herbstreit 2021 | Germany | Retrospective | 2 | 2016.10-2020.3 | 64 | 64.1 | 54.1 | NR | 100 | NR | A(H1N1) (62.0), other A (28.6), B (9.4) |
| Hernandez-Garces 2021 | Spain | Retrospective | 2 | 2010.6-2018.12 | 54 | 40.7 | 66.8 | NR | 100 | NR | A(H1N1)pdm09 (100) |
| Hernandez-Cardenas 2016 | Mexico | Prospective | 2 | 2013.10-2014.5 | 70 | 61.4 | 47.0 | NR | 100 | 5.7 | A(H1N1)pdm09 (100) |
| Hernu 2021 | France | Retrospective | 1 | 2008.12-2013.4 | 201 | 53.4 | 63.0 | NR | 100 | 12 | A(H1N1)pdm09 (51.0), A(H3N2) (32.0), B (15.0) |
| Heyd 2017 | Germany | Retrospective | 1 | 2014.12-2015.5 | 857 | 55.8 | 59.0 | NR | 70.0 | NR | A(H1N1)pdm09 (13.0), A(H3N2) (27.0), other A (38.0), B (21.7) |
| Hiba 2011 | Israel | Retrospective | 1 | 2009.7-2010.1 | 449 | NR | NR | 16-93 | 100 | 17.3 | A(H1N1)pdm09 (100) |
| Hobbs 2019 | Canada | Retrospective | 1 | 2010.9-2014.8 | 29734 | NR | NR | NR | 29.7 | NR | NR |
| Hong 2021 | China | Retrospective | 2 | 2006.1-2016.5 | 99 | 61.6 | 62.2 | NR | 100 | NR | A (83.8), Non-influenza A (16.2) |
| Hoy 2023 | Nicaragua | Prospective, retrospective | 1 | 2011-2020 | 1394 | 50.7 | 5.9 | 0-14 | 1.9 | 3.1 | A(H3N2) (39.4), A(H1N1)pdm09 (24.7), B/Victoria (17.0), B/Yamagata (16.7) |
| Hsieh 2018 | China | Retrospective | 2 | 2009.7-2016.5 | 639 | 57.3 | 12.2 | NR | 70.9 | NR | A(H1N1)pdm09 (100) |
| Hsing 2022 | China | Retrospective | 2 | 2009.1-2019.12 | 558 | 55.0 | 3.2 | 0-18 | 100 | NR | A (62.0), B (38.0) |
| Hu 2023 | China | Retrospective | 2 | 2009.1-2022.12 | 243 | 69.6 | 3.2 | NR | 100 | NR | A (81.5), B (20.2) |
| Huai 2017 | China | Prospective | 1 | 2010.4-2012.4 | 1774 | 57.7 | 2.5 | NR | 100 | 17 | A(H1N1)pdm09 (19.0), A(H3N2) (40.0), B (41.0) |
| Huang 2017 | China | Retrospective | 2 | 2009-2010 | 526 | 59.9 | 13.6 | NR | 7.8 | NR | A(H1N1)pdm09 (85.0), A(H3N2) (15.0) |
| Irving 2012 | USA | Prospective | 2 | 2004-2008 | 1185 | 46.0 | NR | NR | 5.0 | 36 | A (76.0), B (24.0) |
| Jamoussi 2022 | Tunisia | Prospective | 1 | 2009.11.1-2019.10.31 | 120 | 53.3 | 48.0 | NR | 100 | 5.8 | A(H1N1)pdm09 (84.2), A(H3N2) (15.8) |
| Jorda 2023 | Austria | Retrospective | 2 | 2014.2-2021.12 | 180 | 52.2 | 60.2 | ≥18 | 100 | NR | A (74.4), B (25.6) |
| Kandeel 2016 | Egypt | Prospective | 1 | 2007.11-2014.11 | 2936 | NR | 30.0 | 0.08-90 | 100 | NR | A (68.6), B (31.4) |
| Kappagoda 2000 | Australia | Retrospective | 2 | 1977-1994 | 227 | NR | NR | 0.02-11.3 | 100 | NR | A (63.4), B (36.6) |
| Karolyi 2021 | Austria | Prospective | 1 | 2008-2009 | 490 | 51.2 | 73.0 | NR | 100 | 6.1 | A (100) |
| Karolyi 2019 | Austria | Retrospective | 2 | 2017.10-2018.4 | 396 | 46.2 | 75.5 | NR | 100 | 19.6 | A (24.2), B (75.8) |
| Katzen 2019 | USA | Retrospective | 2 | 2009.4-2014.3 | 699 | 46.2 | 55.6 | ≥18 | 100 | NR | A(H1N1) (48.6), A(H3N2) (33.3), Untypeable A (3.4), B (15.2) |
| Khandaker 2014 | Australia | Prospective | 1 | 2009.6-2009.9 | 506 | 58.9 | 3.7 | 0-14.9 | 100 | 11.3 | A(H1N1)pdm09 (100) |
| Kim 2023 | Canada | Prospective | 1 | 2010.9-2017.8 | 1228 | 49.9 | NR | 50-64 | 100 | 33.5 | A(H1N1) (30.1), A(H3N2) (39.9), untyped A (8.1), B (21.8) |
| Kini 2018 | India | Retrospective | 2 | 2017.10-2018.9 | 56 | 60.7 | 5.9 | 0.9-17.5 | 100 | 0 | B (100) |
| Kohlmaier 2020 | Austria | Prospective | 2 | 2017.11-2018.3 | 166 | 56.6 | 2.0 | 1-5 | 100 | 0.8 | A (67.5), B (29.5), A and B (1.2), untyped (1.8) |
| Kojicic 2012 | Bosnia and herzegovina, Serbia | Retrospective | 1 | 2009.11-2010.3 | 50 | 62.0 | 43.0 | 35-54 | 100 | NR | A(H1N1)pdm09 (100) |
| Kok 2013 | Australia | Prospective | 1 | 2009.6-2009.8 | 173 | 45.1 | 46.0 | NR | 100 | NR | A(H1N1)pdm09 (83.8), A(H3N2) (9.8), Seasonal A (0.6), Untyped (5.8) |
| Kosasih 2013 | Indonesia | Retrospective | 1 | 2003.1-2007.12 | 4236 | NR | 21.1 | 0.1-90 | 5.2 | NR | A(H1N1) (19.1), A(H3N2) (35.3), A(H5N1) (0.2), Untyped A (10.3), B (35.1) |
| Kovacevic 2020 | Bosnia and herzegovina, Serbia | Retrospective | 1 | 2019.1-2019.3 | 89 | 55.1 | 56.1 | 27‑83 | 100 | 1.1 | A(H1N1)pdm09 (100) |
| Kraft 2012 | USA | Retrospective | 2 | 2009.9-2009.10 | 99 | 35.4 | 37.1 | ≥18 | 36.4 | NR | A(H1N1)pdm09 (100) |
| Kuchar 2013 | Poland | Retrospective | 2 | 2009.5-2010.5 | 561 | NR | NR | 0-85 | 86.3 | 2 | A(H1N1)pdm09 (100) |
| Kumar 2010 | USA, Canada, Netherlands | Retrospective | 1 | 2009.4-2009.12 | 237 | 54.4 | 32.0 | 1-95 | 70.5 | 69.9 | A(H1N1)pdm09 (100) |
| Kumar 2016 | India | Retrospective | 2 | 2015.1-2015.4 | 181 | 47.0 | 37.1 | NR | NR | NR | Swine flu (H1N1) (100) |
| Kusznierz 2017 | Argentina | Retrospective | 1 | 2013.1-2013.12 | 106 | 53.8 | NR | NR | 100 | NR | A(H1N1)pdm09 (51.8), A(H3N2) (39.6), A no sub-typeable (8.6) |
| Kwon 2017 | South Korea | Retrospective | 2 | 2013.1-2015.12 | 312 | 49.0 | 62.6 | ≥18 | 100 | NR | A (78.2), B (21.8) |
| Lalueza 2020 | Spain | Prospective | 2 | 2016.12-2018.4 | 494 | 53.4 | 74.4 | ≥18 | 100 | 45.2 | A(H1N1)pdm09 (11.7), A(H3N2) (80.6), Non-typable A (7.7) |
| Laris-Gonzalez 2021 | Mexico | Retrospective | 2 | 2013-2018 | 295 | 52.5 | 3.7 | <18 | 100 | NR | NR |
| Lee 2020 | South Korea | Retrospective | 1 | 2017.8-2018.5 | 542 | 48.2 | NR | ≥18 | 100 | NR | A (60.9), B (38.2), A and B (0.9) |
| Lee 2011 | China | Prospective | 1 | 2007.1.1-2010.5.31 | 1136 | 44.5 | 62.1 | >16 | 100 | NR | A (81.4), B (18.6) |
| Lehners 2013 | Germany | Retrospective | 2 | 2009.5-2011.4 | 178 | 51.7 | NR | NR | 100 | NR | A(H1N1)pdm09 (100) |
| Lenzi 2012 | Brazil | Retrospective | 1 | 2010.3-2010.12 | 4740 | 46.7 | NR | 0-90 | 40.3 | 2.8 | A(H1N1)pdm09 (100) |
| Leung 2014 | China | Retrospective | 2 | 2009.4-2011.2 | 1203 | 51.9 | 17.2 | 0.08-99 | 100 | NR | NR |
| Li 2021 | USA | Retrospective | 2 | 2016.6-2019.2 | 3991 | 43.1 | 62.6 | ≥21 | 38.4 | NR | NR |
| Li 2011 | China | Prospective | 1 | 2009.5-2011.4 | 81 | NR | NR | NR | 100 | NR | A(H1N1)pdm09 (100) |
| Liem 2009 | Vietnam | Retrospective | 1 | 2004.1.1-2006.12.31 | 67 | 55.0 | 25.0 | NR | 100 | NR | A(H5N1) (100) |
| Lim 2015 | Singapore | Retrospective | 1 | 2009.6.18-2010.8.10 | 172 | 53.5 | 46.0 | NR | 100 | NR | A(H1N1)pdm09 (100) |
| Lim 2010 | Singapore | Prospective | 2 | 2009.5.26-2009.9.14 | 211 | 0.0 | 29.0 | 16-42 | 29.4 | NR | A(H1N1) (100) |
| Lindblade 2010 | Guatemala | Retrospective | 1 | 2008-2009 | 138 | 39.9 | 3.0 | NR | 100 | NR | A (100) |
| Liu 2020 | China | Retrospective | 2 | 2016.1-2018.12 | 96 | 57.3 | 65.0 | NR | 100 | NR | A (76.0), B (24.0) |
| Lo 2013 | China | Prospective | 1 | 2011.7.1-2012.6.30 | 1704 | 53.2 | NR | NR | 100 | NR | A(H3N2) (32.2), A(H1N1) (5.1), A untyped (2.1), B (60.7) |
| Lopez Montesinos 2022 | Spain | Retrospective | 2 | 2017.1.1-2019.12.1 | 187 | 54.5 | 76.0 | NR | 100 | 17.2 | A(H1N1) (67.0), B (26.7) |
| Lopez-Delgado 2013 | Spain | Prospective | 1 | 2009.8-2011.3 | 60 | 63.3 | 49.2 | NR | 100 | NR | A(H1N1) (100) |
| Lopez-Medina 2012 | USA | Retrospective | 2 | 2009.4.27-2010.3.23 | 73 | 48.0 | 0.1 | 0-0.5 | 100 | NR | A(H1N1) (100) |
| Louie 2009 | USA | Retrospective | 1 | 2009.4.23-2009.8.11 | 1088 | 49.0 | 27.0 | 1-92 | 100 | NR | A(H1N1)pdm09 (100) |
| Louie 2013 | USA | Retrospective | 1 | 2009.4.3-2012.9.30 | 784 | 61.0 | 6.0 | 0-17 | 100 | NR | A(H1N1) (73.2), B (6.8) |
| Louriz 2010 | Morocco | Retrospective | 2 | 2009.6-2009.12 | 186 | 57.0 | 17.6 | 0.08-57 | 100 | NR | A(H1N1)pdm09 (100) |
| Lovato-Salas 2010 | Mexico | Prospective | 2 | 2009.4-2010.3 | 192 | NR | NR | NR | 100 | NR | A(H1N1)pdm09 (94.7), other (5.3) |
| Lucker 2011 | Switzerland | Prospective | 2 | 2009.10-2010.1 | 85 | 53.0 | 39.0 | 0.08-94 | 100 | 5 | A(H1N1)pdm09 (100) |
| Lynfield 2014 | 17 countries | Prospective | 1 | 2009.10-2012.12 | 982 | 47.9 | NR | ≥18 | 39.9 | NR | A(H1N1)pdm09 (100) |
| Lytras 2020 | Greece | Prospective | 1 | 2010-2011 to 2018-2019 | 2325 | NR | NR | NR | 100 | NR | NR |
| Mabayoje 2021 | UK | Retrospective | 2 | 2017-2018 | 127 | 42.5 | NR | ≥18 | NR | NR | A (55.9), B (44.1) |
| Malhotra 2016 | India | Retrospective | 1 | 2015.1-2015.3 | 6203 | 51.7 | NR | NR | NR | NR | A(H1N1)pdm09 (100) |
| Mansour 2012 | Saudi Arabia | Prospective | 2 | 2009.10-2010.1 | 89 | 51.7 | 6.7 | 0.13-15 | 73.0 | NR | A(H1N1)pdm09 (100) |
| Mao 2014 | China | Retrospective | 1 | 2013.2-2014.2 | 334 | 67.0 | 58.0 | NR | 93.7 | NR | H7N9 avian influenza A (100) |
| Marbus 2020 | Netherlands | Retrospective | 1 | 2014-2016 | 380 | 57.4 | 64.5 | ≥18 | 100 | NR | A (77.4), B (20.0), A and B (2.6) |
| Martin 2013 | USA | Retrospective | 1 | 2011.1-2011.3 | 161 | 64.6 | 43.5 | ≥18 | 62.7 | 34.5 | A(H1N1) (69.6), H3N2 (18.6), B (11.8) |
| Martinez-Briseno 2016 | Mexico | Retrospective | 1 | 2013.1-2014.3 | 99 | 63.6 | 46.9 | ≥18 | 100 | NR | A(H1N1) (100) |
| Martin-Loeches 2011 | Spain, Brazil, UK, Portugal, Israel, France, Ecuador, Colombia, Argentina | Prospective | 2 | 2009.6-2010.2 | 220 | 51.4 | 43.3 | NR | 100 | NR | A(H1N1)pdm09 (100) |
| Martin-Loeches 2017 | Spain | Prospective | 1 | 2009-2015 | 2901 | 59.1 | 51.6 | >16 | 100 | NR | NR |
| Martin-Loeches 2019 | Ireland, Netherlands, Brazil, Denmark, Spain, USA, Belgium, Czech Republic, Norway, France, Canada, Italy, Finland, UK, Uruguay, Brazil | Prospective | 1 | 2015.11-2016.7 | 153 | 59.5 | 63.0 | ≥18 | 100 | NR | NR |
| Maruyama 2016 | Japan | Prospective | 1 | 2010.10-2013.5 | 1345 | 52.8 | 34.9 | ≥1 | 100 | 38.6 | A (80.8), B (19.2) |
| Mattila 2020 | Finland | Retrospective | 2 | 2004.7-2018.6 | 391 | 56.5 | 4.9 | <16 | 100 | NR | A (71.4), B (28.6) |
| McGeer 2007 | Canada | Prospective | 1 | 2005.1-2006.5 | 512 | NR | NR | 0-99 | 100 | NR | A (81.0) |
| McGrath 2023 | USA | Retrospective | 1 | 2019.4-2020.3 | 4349 | 57.5 | 1.4 | <5 | 100 | NR | NR |
| McNeil 2014 | Canada | Prospective | 1 | 2013.11-2014.2 | 654 | 48.9 | 58.5 | 16-98 | 100 | 34.7 | A(H1N1) (54.6), A(H3N2) (2.8), A subtypes unknown (39.1), B (3.4) |
| McRae 2022 | Australia | Prospective, retrospective | 1 | 2011-2019 | 680 | 57.9 | 0.2 | 0.12-0.35 | 100 | NR | A(H1N1) (12.7), A(H3N2) (15.7), A Unsubtyped (72.1), B (20.3), Multiple subtypes (1.8) |
| Mehta 2013 | India | Prospective | 2 | 2009.8-2011.12 | 115 | 50.0 | 31.6 | 1-78 | 76.5 | NR | Swine-origin A(H1N1) (100) |
| Mehta 2016 | India | Retrospective | 2 | 2009.8-2013.3 | 166 | 55.4 | NR | NR | 100 | NR | A(H1N1)pdm09 (42.2), Seasonal influenza (57.8) |
| Mendez-Dominguez 2019 | Mexico | Retrospective | 2 | 2018 | 264 | 45.0 | 27.1 | NR | 37.9 | 18.6 | NR |
| Miller 2010 | USA | Retrospective | 1 | 2009.5-2009.6 | 47 | 43.0 | 34.0 | 15-62 | 100 | NR | A(H1N1)pdm09 (100) |
| Minney-Smith 2019 | Australia | Retrospective | 2 | 2012.1-2015.12 | 242 | 52.1 | NR | ≥18 | 100 | NR | A(H1N1) (26.9), A(H3N2) (73.1) |
| Miron 2021 | Romania | Prospective | 2 | 2019.11-2020.3 | 516 | 54.1 | 6.2 | NR | 100 | 6.4 | A(H1N1) (20.0), A(H3N2) (30.4), A Unsubtyped (2.3), B Victoria (45.9), B lineage (1.2), co-infection with A(H3)+B Victoria (0.4) |
| Modemann 2022 | Germany | Retrospective | 2 | 2012.1-2021.1 | 50 | 68.0 | 58.5 | NR | 100 | NR | A (80.0), B (20.0) |
| Montes 2005 | Spain | Retrospective | 2 | 2001.7-2004.6 | 49 | 55.1 | NR | NR | 100 | 0 | A(H1N1) (6.1), A(H3N2) (83.7), Unknown (2.0), B (8.2) |
| Moreno 2021 | Spain | Retrospective | 1 | 2009.6-2018.4 | 2124 | 61.2 | 54.0 | 43-65 | 100 | 14.5 | A(H1N1)pdm09 (72.2), A(H3N2) (15.8), B (7.0), A Unsubtyped (5.0) |
| Murti 2018 | Canada | Retrospective | 1 | 2012.9-2016.8 | 1586 | NR | NR | NR | 100 | NR | NR |
| Muscatello 2014 | Australia | Retrospective | 1 | 2005-2008 | 2568 | 53.7 | NR | NR | 56.5 | NR | A (70.0), B (14.8) |
| Nandhini 2015 | India | Retrospective | 1 | 2009.5-2013.12 | 403 | 48.3 | NR | NR | 48.6 | NR | A(H1N1) (71.2), A(H3N2) (22.8), B (6.0) |
| Nasir 2021 | Pakistan | Retrospective | 2 | 2017-2019 | 55 | 40.0 | 55.0 | 36-65 | 100 | NR | A(H1N1) (100) |
| Nation 2021 | Australia | Retrospective | 1 | 2010-2017 | 9298 | 49.7 | NR | NR | 100 | 45.9 | NR |
| Nguyen 2013 | Vietnam | Prospective | 1 | 2006.1.1-2010.12.31 | 6516 | 52.2 | 12.0 | 0.08-94 | 8.1 | NR | A(H1N1) (19.9), A(H3N2) (34.5), B (33.2), A(H1N1)pdm09 (12.1) |
| Nichols 2018 | Canada | Prospective | 1 | 2011-2014 | 3394 | 46.8 | 67.6 | NR | 100 | 46.7 | A(H1N1) (32.5), A(H3N2) (24.1), A-not subtyped (10.2), B/Victoria (3.0), B/Yamagatha (22.9), B not subtyped (22.0), unknown (0.0) |
| Nickel 2011 | USA | Retrospective | 2 | 2009.4-2009.9 | 184 | 37.0 | NR | NR | 100 | NR | A(H1N1)pdm09 (100) |
| Nieto-Guevara 2011 | Panama | Prospective | 1 | 2009.4-2010.1 | 806 | 50.0 | 13.0 | 0-88 | 100 | 15.9 (<2 years), 5 (>60 years), 0 (other age) | A(H1N1) (100) |
| Nordenskjold 2022 | Sweden | Prospective | 1 | 2017-2020 | 181 | 52.5 | 55.0 | ≥18 | 27.6 | 29.2 | A (52.0), B (48.0) |
| Oliva 2018 | Spain | Retrospective | 1 | 2010-2016 | 21693 | NR | NR | NR | 100 | NR | A(H1N1)pdm09 (69.0), A(H3N2) (25.6), B (5.6) |
| Ono 2016 | Japan | Retrospective | 1 | 2013.10-2014.12 | 88054 | 48.9 | NR | <65 | 0.3 | NR | A (62.3), B (36.5), A and B (1.2), Unknown (<0.01) |
| Pang 2021 | Malaysia | Prospective | 1 | 2013.7-2019.8 | 114 | 40.4 | 59.1 | ≥18 | 100 | 1.8 | A (85.1), B (5.6) |
| Parisi 2023 | USA | Retrospective | 1 | 2016-2017, 2018-2019 | 48430 | 44.6 | 66.1 | ≥18 | 100 | 58.2 | NR |
| Pascale 2021 | Panama | Retrospective | 1 | 2011-2017 | 1839 | 49.2 | NR | NR | 50.6 | NR | A (79.6), B (20.4) |
| Pedroni 2010 | Chile | Retrospective | 2 | 2008-2009 | 190 | 47.4 | 27.0 | NR | 100 | NR | NR |
| Phung 2011 | Australia | Retrospective | 1 | 2009.5.25-2009.10.3 | 1236 | 45.9 | 29.0 | NR | 100 | NR | A(H1N1) (100) |
| Plumb 2021 | UK | Retrospective | 1 | 2009.5.27-2010.1.5 | 2380 | 48.2 | NR | NR | 100 | NR | A(H1N1)pdm09 (100) |
| Poggensee 2010 | Germany | Retrospective | 1 | 2009.4.27-2009.11.11 | 53968 | NR | NR | NR | 3.9 | NR | A(H1N1) (100) |
| Puig-Barbera 2014 (a) | Spain | Prospective | 1 | 2011.12.18-2012.3.31 | 544 | 45.0 | NR | ≥18 | 100 | 58 | A(H2N3) (100) |
| Puig-Barbera 2016 | Spain, Russia, Turkey, China | Prospective | 1 | 2013-2014 | 1086 | 49.6 | 34.9 | NR | 100 | 12.7 | A(H1N1) (33.3), A(H3N2) (49.2), A untyped (3.7), B (13.9) |
| Puig-Barbera 2014 (b) | Spain, France, Russia, Turkey | Prospective | 1 | 2012-2013 | 1545 | 49.5 | NR | NR | 100 | 9.97 | A (62.1), B (37.9) |
| Punpanich 2014 | Thailand | Retrospective | 2 | 2010.1.1-2010.12.31 | 289 | 60.2 | 3.7 | 0.08-18 | 100 | NR | A (70.6), B (27.3), A and B (2.1) |
| Raff 2022 | USA | Retrospective | 2 | 2013.11-2020.3 | 28 | 75.0 | 41.2 | NR | 100 | NR | NR |
| Samransamruajkit 2008 | Thailand | Prospective | 2 | 2006.3-2007.2 | 32 | 43.8 | 2.0 | 0.08-15 | 100 | NR | NR |
| Saroch 2018 | India | Retrospective | 2 | 2015.11-2016.4 | 30 | 36.7 | 40.0 | ≥14 | 100 | NR | A(H1N1) (100) |
| Satterwhite 2010 | USA | Retrospective | 1 | 2009.8.1-2009.10.31 | 109 | 40.4 | 41.1 | NR | 100 | NR | A(H1N1)pdm09 (100) |
| Schauwvlieghe 2018 | Belgium, Netherlands | Retrospective | 1 | 2009.1.1-2016.6.30 | 432 | 55.6 | 59.0 | ≥18 | 100 | NR | A (82.0), B (18.0) |
| Schober 2023 | Canada | Prospective | 1 | 2010.9.1-2021.8.31 | 8982 | 52.6 | 4.2 | 0-16 | 100 | NR | A (67.6), A and B (0.5), B (31.9) |
| SerpaNeto 2021 | Australia | Prospective | 1 | 2009.6.1-2009.8.31 | 625 | 48.5 | 42.0 | NR | 100 | NR | A(H1N1) (100) |
| Shalabi 2022 | Israel | Retrospective | 2 | 2009.9-2020.5 | 1071 | 54.9 | 2.5 | <18 | 100 | NR | A (74.0), B (26.6), A and B (0.6) |
| Shannon 2022 | USA | Retrospective | 1 | 2014.11.1-2015.4.30 | 1070 | NR | 45.0 | 30-55 | NR | NR | A (83.8), B (17.2) |
| Sharma 2020 | Australia | Retrospective | 1 | 2016.1-2020.4 | 1846 | 47.1 | 66.5 | ≥18 | 100 | NR | A (88.3), B (11.7) |
| Sherban 2023 | Israel | Retrospective | 2 | 2017.10-2018.4 | 625 | 53.3 | 70.8 | ＞65 | 100 | 15.2 | A (31.2), B (50.9) |
| Shimada 2015 | Japan | Prospective | 1 | 2009.7-2010.9 | 13581 | 63.4 | 7.0 | 0-102 | 100 | 2 | A(H1N1)pdm09 (100) |
| Shusterman 2023 | Israel | Retrospective | 2 | 2020.3-2021.11 | 2041 | 47.9 | 73.0 | NR | 100 | NR | NR |
| Susilarini 2018 | Indonesia | Retrospective | 1 | 2013.5-2016.4 | 199 | 53.8 | NR | NR | 100 | 2.01 | NR |
| Sutton 2020 | USA | Retrospective | 2 | 2011.1-2019.1 | 12806 | 87.2 | 57.2 | NR | NR | NR | NR |
| Talbot 2021 | USA | Retrospective | 1 | 2016-2019 | 1937 | 44.2 | 63.0 | ≥18 | 100 | 61.8 | A(H3N2) (59.9), A(H1N1) (18.7), B (21.5) |
| Tamma 2010 | USA | Retrospective | 2 | 2007-2009 | 266 | 51.5 | 7.2 | ≤18 | 100 | NR | A(H1N1) (100) |
| Taniguchi 2022 | Japan | Retrospective | 1 | 2017-2020 | 27870 | 53.2 | 67.0 | NR | 100 | NR | NR |
| Taylor 2016 | Canada | Prospective | 1 | 2006-2012 | 3866 | 45.4 | 69.3 | 16-104 | 100 | NR | NR |
| Tempia 2017 | South Africa | Prospective | 1 | 2012.5-2015.4 | 687 | 57.2 | NR | NR | 100 | NR | B (36.7), A(H1N1)pdm09 (14.7), A(H3N2) (48.6) |
| Teng 2019 | China | Retrospective | 2 | 2010-2018 | 209 | 65.1 | 59.0 | NR | 100 | 0 | A(H1N1) (50.2), A unclassified (33.5), B (13.9), A and B (2.4) |
| Teros-Jaakkola 2019 | Finland | Prospective | 2 | 2008.1-2010.4 | 56 | 55.6 | 10.4 | 0.02-1.9 | 5.4 | NR | A(H1N1) (28.6), A(H3N2) (25.0), B (46.4) |
| Thangaraj 2023 | India | Retrospective | 2 | 2013.6-2018.6 | 68 | 58.0 | 1.0 | 0.08-4.9 | 100 | NR | A(H1N1)pdm09 (63.2), A(H3N2) (2.2), B (34.1) |
| Thelen 2021 | Netherlands | Retrospective | 1 | 2015-2018 | 653 | 53.8 | 72.5 | NR | NR | NR | A (49.8), B (50.2) |
| To 2010 | China | Retrospective | 2 | 2009.6-2009.10 | 186 | 48.4 | NR | NR | 100 | NR | A(H1N1)pdm09 (37.1), A(H1N1) (10.8), A(H3N2) (52.1) |
| Torres 2010 | Chile | Retrospective | 2 | 2009.5-2009.7 | 4591 | 50.6 | 13.0 | NR | 4.1 | 1.3 | A(H1N1) (100) |
| Tramuto 2011 | Italy | Retrospective | 1 | 2009.4-2010.12 | 1193 | 52.1 | 18.0 | NR | 100 | NR | A(H1N1)pdm09 (100) |
| Tran 2016 | Canada | Retrospective | 1 | 2004.9-2013.6 | 4155 | 58.5 | NR | NR | 100 | NR | A (63.7), B (36.3) |
| Truelove 2011 | USA | Prospective | 1 | 2009.4-2010.1 | 1329 | 54.2 | NR | NR | 100 | NR | A(H1N1) (100) |
| Tsukui 2012 | Japan | Prospective | 1 | 2009.7-2009.11 | 7781 | NR | 12.0 | 1-87 | 2.4 | NR | NR |
| Unal 2023 | Germany | Retrospective | 2 | 2014-2019 | 159 | 56.0 | 57.1 | 21-83 | 47.8 | NR | A (59.8), B (40.2) |
| van't Klooster 2010 | Netherlands | Retrospective | 1 | 2009.6-2009.12 | 2181 | 47.1 | 19.0 | 0-89 | 100 | NR | A(H1N1)pdm09 (100) |
| Vandroux 2019 | France | Retrospective | 2 | 2013.1-2017.12 | 127 | 59.8 | 59.0 | NR | 100 | 3.1 | A(H1N1)pdm09 (45.0), A non-H1N1 (24.0), B (31.0) |
| Viasus 2012 | Spain | Prospective | 1 | 2009.6-2009.12, 2010.12-2011.3 | 348 | 56.3 | 44.0 | 30-60 | 100 | 4.8 | A(H1N1)pdm09 (100) |
| vonBaum 2011 | Germany | Prospective | 1 | 2002.6-2007.4 | 160 | NR | NR | NR | 63.8 | 26 | A (83.8), B (16.2) |
| Borgatta 2012 | Spain | Prospective | 1 | 2009.6-2010.1 | 505 | 56.4 | 43.0 | 33-54 | 100 | NR | A(H1N1)pdm09 (100) |
| Borja-Aburto 2012 | Mexico | Prospective | 1 | 2011.12-2012.3 | 1115 | NR | NR | NR | 100 | NR | A(H1N1)pdm09 (100) |
| Bouneb 2018 | Tunisia | Retrospective | 2 | 2010.10-2016.4 | 40 | 60.0 | 53.0 | NR | 100 | 0 | A(H1N1)pdm09 (100) |
| Bunthi 2013 | Thailand | Retrospective | 1 | 2009.5-2010.1 | 27254 | NR | NR | NR | NR | NR | A(H1N1)pdm09 (100) |
| Burkert 2022 | Austria | Retrospective | 2 | 2012.1-2019.3 | 250 | 54.4 | 65.9 | ≥18 | 100 | NR | NR |
| Burton 2008 | Canada | Prospective | 1 | 2006-2007 | 371 | 38.8 | NR | 0-16 | 100 | 14 | A (83.8), B (15.4), A and B (0.8) |
| Campbell 2020 | USA | Retrospective | 1 | 2018-2019 | 226 | 44.0 | NR | 0.5-17 | 100 | 47 | A(H1N1)pdm09 (39.0), A(H3N2) (51.0), B (7.0) |
| Campbell 2021 | USA | Prospective | 1 | 2019-2020 | 335 | 36.0 | NR | <18 | 100 | NR | A (50.0), B (50.0) |
| Campbell 2011 | UK | Retrospective | 1 | 2009.4.1-2010.6 | 2416 | 48.1 | 20.0 | NR | 100 | NR | A(H1N1)pdm09 (100) |
| Casas-Aparicio 2018 | Mexico | Retrospective | 2 | 2014.11-2015.5 | 60 | 65.0 | 47.5 | ≥18 | 100 | 1.7 | A(H1N1) (100) |
| Castillo-Palencia 2012 | Mexico | Retrospective | 2 | 2009.3-2009.10 | 2767 | 48.0 | NR | NR | 5.8 | NR | A(H1N1)pdm09 (100) |
| Chakhunashvili 2018 | USA | Retrospective | 1 | 2015.9-2019.3 | 542 | 54.0 | NR | NR | 100 | 2 | A(H1N1)pdm09 (61.1), A(H3N2) (24.7), B (14.2) |
| Chan 2015 | China | Retrospective | 2 | 1998-2012 | 9635 | NR | NR | NR | 100 | NR | A (79.6), B (20.6) |
| Chan 2011 | USA | Retrospective | 1 | 2009.10.16-2009.12.1 | 291 | 52.9 | 20.0 | 0-97 | 100 | NR | A(H1N1)pdm09 (100) |
| Chan 2017 | China | Retrospective | 2 | 2014.10.1-2015.9.30 | 110 | 56.4 | 80.7 | ≥65 | 100 | NR | A (94.5), B (5.5) |
| Chatterjee 2020 | India | Retrospective | 1 | 2010-2017 | 114667 | NR | NR | NR | NR | NR | A(H1N1) (100) |
| Chaves 2013 | USA | Retrospective | 1 | 2010.10-2011.4 | 3621 | 48.0 | NR | 2-65 | 100 | 33.6 | A (73.8), B (26.2) |
| Chaves 2023 | USA | Retrospective | 1 | 2016-2019 | 44456 | 42.0 | NR | ≥65 | 37.6 | NR | NR |
| Chaves 2014 | USA | Retrospective | 1 | 2003-2012 | 3157 | 58.0 | NR | 0-1 | 100 | NR | A (82.0), B (11.0) |
| Chen 2013 | USA | Prospective | 1 | 2007-2009 | 146 | 55.0 | 10.0 | NR | 38.4 | 38 | A (62.0) |
| Chen 2021 | China | Retrospective | 1 | 2012.1-2018.12 | 1313 | 54.1 | 59.0 | ＞14 | 100 | NR | A (35.0) |
| Cheng 2015 | Austria | Retrospective | 1 | 2014.3-2014.10 | 1692 | 45.9 | NR | >16 | 100 | 50 | A(H1N1)pdm09 (16.0), A(H3N2) (18.3), A unsubtyped (58.7), B (6.8) |
| Rao 2023 | USA | Retrospective | 2 | 2021.10-2022.4 | 51 | 55.3 | 6.1 | < 18 | 100 | 68 | NR |
| Rao 2020 | USA | Prospective | 2 | 2017.11-2018.4 | 411 | 50.1 | 4.0 | 0.5-8 | 6.3 | 36 | A (56.0), B (44.0) |
| Reacher 2019 | UK | Retrospective | 2 | 2016.8-2017.3 | 332 | 47.3 | 68.6 | 0-102 | 100 | 33.1 | A(H3N2) (100) |
| Reed 2014 | USA | Retrospective | 1 | 2005.4-2009.10, 2009.10-2010.4 | 10232 | 42.5 | NR | ≥18 | 100 | 34.9 | A(H1N1)pdm09 (48.5) |
| Regan 2023 | USA | Retrospective | 1 | 2013-2019 | 2747 | 52.8 | NR | NR | 100 | 37.9 | A(H3N2) (41.5), A(H1N1) (35.0), B (17.0) |
| Reyes 2007 | Canada | Retrospective | 1 | 2005.8-2006.8 | 374 | NR | NR | NR | 100 | 84.2 | A (61.0), B (39.0) |
| Reyes 2010 | Guatemala | Retrospective | 1 | 2009.5-2009.12 | 239 | 56.9 | 8.8 | 0.05-82 | 31.8 | NR | A(H1N1) (100) |
| Ristic 2018 | Serbia | Retrospective | 1 | 2010.10-2015.5 | 720 | 51.4 | NR | NR | 55.8 | NR | A(H1N1)pdm09 (40.0), A(H3N2) (35.1), B (21.9), A without subtypes (3.0) |
| Rodriguez 2011 | Spain | Prospective | 1 | 2010-2011 | 300 | 62.6 | 49.0 | ≥16 | 100 | 7.3 | A(H1N1) (100) |
| Roedl 2021 | Germany | Retrospective | 2 | 2009.1-2021.1 | 61 | 69.0 | 52.0 | ≥18 | 100 | NR | NR |
| Rosler 2021 | Germany | Retrospective | 1 | 2017.12-2018.4 | 1539 | 50.0 | 72.0 | 0-102 | 100 | NR | NR |
| Rothman 2023 | Sweden | Prospective | 1 | 2013.12-2019.5 | 4110 | 49.9 | 75.0 | ≥18 | 100 | NR | A(H1N1)pdm09 (23.1), A(H3N2) (45.2), B (31.1) |
| Rovina 2014 | Greece | Retrospective | 2 | 2009.4-2010.12 | 90 | 43.3 | 37.0 | NR | 100 | 0.9 | A(H1N1)pdm09 (100) |
| Rozencwajg 2018 | France | Retrospective | 2 | 2009.10-2016.5 | 77 | 69.0 | NR | NR | 100 | NR | A(H1N1)pdm09 (94.0), B (6.0) |
| Smiechowicz 2021 | Poland | Retrospective | 1 | 2018.11-2019.3 | 76 | 51.0 | 62.0 | 19-86 | 100 | NR | A (98.6), A(H1N1)pdm09 (53.3), B (1.3) |
| Smit 2012 | Netherlands | Prospective | 2 | 2009.8-2009.12 | 132 | 53.0 | 6.8 | 0-17 | 16.7 | NR | A(H1N1) (100) |
| Snacken 2012 | Austria, Finland, France, Ireland, Malta, Portugal, Romania, Slovakia, Spain | Retrospective | 1 | 2010-2011 | 2271 | 55.4 | 45.5 | NR | 100 | 9.7 | A(H1N1)pdm09 (100) |
| Sohn 2013 | South Korea | Prospective | 2 | 2009.9-2009.12 | 59 | 52.5 | 46.0 | >15 | 100 | NR | A(H1N1) (100) |
| Soldevila 2020 | Spain | Retrospective | 1 | 2013.12-2015.3 | 715 | 53.0 | NR | ≥65 | 100 | 49.2 | A (94.6), B (5.2) |
| Song 2013 | South Korea | Retrospective | 1 | 2011-2012 | 7213 | NR | NR | NR | 20.0 | NR | A (54.0), B (44.6), undermined (1.4) |
| Song 2020 | USA | Retrospective | 2 | 2019.10-2020.6 | 1402 | 53.0 | 3.9 | 0.03-40.4 | 20.8 | NR | A (48.1), B (51.9) |
| Soyemi 2014 | USA | Retrospective | 1 | 2009.4-2009.12 | 2824 | 47.2 | 23.7 | NR | 100 | NR | A(H1N1)pdm09 (100) |
| Stein 2010 | Israel | Prospective | 1 | 2009.7-2009.12 | 478 | 55.9 | 6.1 | 0.03-18 | 100 | NR | A(H1N1) (100) |
| Streng 2011 | Germany | Prospective | 1 | 2005.10-2008.7 | 20 | 60.0 | 7.5 | 0.1-15 | 100 | 0 | A (70.0), B (25.0), unknown (5.0) |
| Streng 2018 | Germany | Prospective | 1 | 2013-2015 | 217 | 49.3 | 3.7 | <6 | 0.0 | 0 | A(H3N2) (56.2), A(H1N1)pdm09 (25.8), B (18.0) |
| Subramony 2010 | Singapore | Retrospective | 1 | 2009.7-2009.9 | 1348 | 48.7 | 25.0 | NR | 100 | NR | A(H1N1)pdm09 (100) |
| Wallemacq 2022 | Belgium | Retrospective | 2 | 2015.1-2020.12 | 225 | 49.8 | 69.8 | ≥18 | 100 | NR | A(H1N1)pdm09 (100) |
| Wane 2012 | Rwanda | Retrospective | 1 | 2009.10-2010.5 | 532 | 48.3 | 19.4 | 0.4-62 | 12.0 | NR | A(H1N1)pdm09 (92.8), A(H3N2) (2.8), B (4.0) |
| Watanabe 2021 | Japan | Retrospective | 2 | 2014.9-2018.8 | 3741 | 52.6 | 5.6 | 0.1-15.7 | 4.2 | NR | NR |
| Webb 2011 | Australia, New Zealand | Prospective | 1 | 2009.6-2009.10, 2010.6-2010.10 | 1428 | NR | NR | NR | 100 | 13.7 | A(H1N1) (86.8), A(H3N2) (1.9) |
| Wei 2021 | USA | Retrospective | 1 | 2013-2014 | 132965 | 45.2 | 66.7 | NR | 100 | NR | NR |
| Wie 2013 | South Korea | Prospective | 1 | 2011.10-2012.5 | 850 | 60.1 | 46.3 | ≥18 | 9.3 | 23.3 | A (77.2), B (22.8) |
| Wong 2014 | Canada | Retrospective | 1 | 2012.8-2013.8 | 31737 | NR | NR | NR | NR | NR | A (85.1), B (14.9) |
| Wong 2016 | China | Retrospective | 2 | 2009.6-2009.12 | 6458 | NR | NR | NR | 100 | NR | A(H1N1)pdm09 (100) |
| Wongwiwatwaitaya 2014 | Thailand | Retrospective | 2 | 2009.5-2011.12 | 49 | NR | 7.5 | 0-15 | 61.2 | NR | A(H1N1)pdm09 (100) |
| Wu 2010 | China | Retrospective | 2 | 2009.4-2009.12 | 22794 | NR | NR | 5-59 | 18.7 | NR | A(H1N1)pdm09 (100) |
| Xiao 2015 | China | Retrospective | 2 | 2014.1-2014.4 | 128 | 67.2 | 58.9 | 20-86 | 100 | NR | A(H7N9) (100) |
| Xie 2020 | USA | Retrospective | 2 | 2017.1-2019.12 | 12676 | 94.6 | 70.3 | NR | 100 | NR | NR |
| Xu 2013 | USA | Retrospective | 1 | 2010.12-2011.5 | 701 | 58.6 | 22.0 | NR | 100 | 23.1 | A(H1N1)pdm09 (100) |
| Yang 2010 | China | Retrospective | 1 | 2009.1-2009.12 | 617 | 60.0 | NR | NR | 100 | NR | A(H1N1)pdm09 (100) |
| Yang 2014 | China | Prospective | 1 | 2010.12-2011.4 | 88 | 54.5 | 52.0 | NR | 100 | 8 | A (100) |
| Yang 2017 | China | Retrospective | 1 | 2013.8-2017.3 | 256 | 66.4 | 56.0 | 1-88 | 100 | NR | H7N9 (100) |
| Yen 2012 | USA | Retrospective | 1 | 2009.4-2010.5 | 8959 | NR | NR | NR | 100 | NR | A(H1N1) (100) |
| Yi 2023 | China | Retrospective | 2 | 2017-2019 | 174 | 60.9 | 61.5 | NR | 100 | NR | A (90.2), B (8.6), A and B (1.1) |
| Ylipalosaari 2017 | Finland | Retrospective | 2 | 2009-2010, 2012-2016 | 76 | 65.8 | NR | NR | 100 | 19.7 | A(H1N1)pdm09 (100) |
| Yokomichi 2019 | Japan | Retrospective | 1 | 2012.1-2016.12 | 16636913 | 53.4 | NR | 0-74 | 1.0 | NR | NR |
| Yoon 2021 | South Korea | Retrospective | 1 | 2014.8-2015.8 | 1747 | 40.9 | 51.7 | NR | 22.4 | 41.7 | A(H1N1)pdm09 (13.7), A(H3N2) (86.3) |
| Youngs 2019 | UK | Retrospective | 2 | 2017-2018 | 808 | 48.6 | 65.4 | NR | 100 | NR | A (60.1), B (39.9) |
| Zhang 2012 | China | Retrospective | 1 | 2009.9-2009.12 | 394 | 0.0 | 25.0 | NR | 100 | NR | A(H1N1)pdm09 (100) |
| Zhang 2018 | China | Prospective | 1 | 2014-2016 | 453 | 59.2 | 33.1 | 14-44 | 100 | NR | A (41.5), B (41.5) |
| Zheng 2020 | China | Retrospective | 1 | 2013-2017 | 350 | 66.9 | 57.0 | NR | 100 | NR | A(H7N9) (100) |
| Zogheib 2018 | USA | Retrospective | 2 | 2009.9-2016.3 | 27 | 63.0 | 56.0 | NR | 100 | NR | A(H1N1)pdm09 (100) |
| HPA 2009 | UK | Prospective | 1 | 2009.4-2009.6 | 252 | 53.0 | 20.0 | 0-73 | 1.6 | NR | A(H1N1) (100) |
| Cowling 2013 | China | Retrospective | 1 | 2003.11.23-2013.2.3 | 43 | 51.2 | 26.0 | NR | 100 | NR | A(H5N1) (100) |
| Jiang 2017 | China | Retrospective | 1 | 2010.9-2019.9 | 17 | 41.2 | 40.0 | NR | 100 | NR | A(H5N6) (100) |
| Wang 2017 | China | Retrospective | 1 | 2013.2-2017.2 | 1220 | 70.2 | 58.0 | 1-93 | 100 | NR | A(H7N9) (100) |

NR, not reported.

### Appendix 4. Risk of bias for eligible studies

| **Study** | **Study participation** | **Study attrition** | **Outcome measurement** | **Statistical analysis and reporting** | **Overall quality** |
| --- | --- | --- | --- | --- | --- |
| **Hospitalization** | | | | | |
| Aguirre 2011 | Low | Low | Low | Low | Low |
| Al Ali 2021 | Low | Low | Low | Low | Low |
| Al Subaie 2012 | Low | Low | Low | Low | Low |
| Al-Busaidi 2016 | Moderate | Low | Low | Low | Low |
| Angeles-Sistac 2020 | Low | Low | Low | Low | Low |
| Andres 2019 | Low | Low | Low | Low | Low |
| CDC 2009 | Low | Low | Low | Low | Low |
| CDESS 2022 | Moderate | Low | Low | Low | Low |
| Assaf-Casals 2020 | Moderate | Low | Low | Low | Low |
| Belchior 2011 | Moderate | Low | Low | Low | Low |
| Chien 2022 | Low | Low | Low | Low | Low |
| Choi 2017 | Low | Low | Low | Low | Low |
| Chuang 2012 | Moderate | Low | Low | Low | Low |
| Cost 2011 | Low | Low | Low | Low | Low |
| Dawood 2010 | Moderate | Low | Low | Low | Low |
| Dawood 2009 | Low | High | Low | Low | High |
| Deguchi 2001 | Low | Low | Low | Low | Low |
| Dwyer 2017 | Low | Low | Low | Low | Low |
| Echevarria-Zuno 2009 | Low | Low | Low | Low | Low |
| Emborg 2022 | Low | Low | Low | Low | Low |
| Esposito 2011 | Low | Low | Low | Low | Low |
| Ezzine 2023 | Low | Low | Low | Low | Low |
| Galindo-Fraga 2013 | Moderate | Low | Low | Low | Low |
| Geerdes-Fenge 2022 | Low | Low | Low | Low | Low |
| Ghosh 2021 | Moderate | Low | Low | Low | Low |
| Gioula 2010 | Low | Low | Low | Low | Low |
| Gutierrez-Pizarraya 2012 | Low | Low | Low | Low | Low |
| Hagiwara 2022 | Low | Low | Low | Low | Low |
| Han 2021 | Low | Low | Low | Low | Low |
| Hoy 2023 | Low | Low | Low | Low | Low |
| Irving 2012 | Low | Low | Low | Low | Low |
| Kosasih 2013 | Moderate | Low | Low | Low | Low |
| Kraft 2012 | Low | Low | Low | Low | Low |
| Kuchar 2013 | Moderate | Low | Low | Low | Low |
| Kumar 2010 | Low | Low | Low | Low | Low |
| Kusznierz 2017 | Moderate | Low | Low | Low | Low |
| Lenzi 2012 | Low | Low | Low | Low | Low |
| Li 2021 | Low | Low | Low | Low | Low |
| Lim 2010 | Low | Low | Low | Low | Low |
| Lynfield 2014 | Low | Low | Low | Low | Low |
| Mansour 2012 | Low | Low | Low | Low | Low |
| Mao 2014 | Low | High | Low | Low | High |
| Mehta 2013 | Moderate | Low | Low | Low | Low |
| Muscatello 2014 | Low | Low | Low | Low | Low |
| Nandhini 2015 | Low | Low | Low | Low | Low |
| Nguyen 2013 | Low | Low | Low | Low | Low |
| Nordenskjold 2022 | Low | Low | Low | Low | Low |
| Ono 2016 | Low | Low | Low | Low | Low |
| Pascale 2021 | Low | Moderate | Low | Low | Low |
| Poggensee 2010 | Low | Low | Low | Low | Low |
| Shannon 2022 | Low | Low | Low | Low | Low |
| Sutton 2020 | Low | Low | Low | Low | Low |
| Teros-Jaakkola 2019 | Low | Low | Low | Low | Low |
| Torres 2010 | Low | Low | Low | Low | Low |
| Tsukui 2012 | Low | Low | Low | Low | Low |
| Unal 2023 | Low | Low | Low | Low | Low |
| vonBaum 2011 | Low | Low | Low | Low | Low |
| Castillo-Palencia 2012 | Low | Low | Low | Low | Low |
| Chaves 2023 | Low | Low | Low | Low | Low |
| Chen 2013 | Low | Low | Low | Low | Low |
| Rao 2020 | Low | Low | Low | Low | Low |
| Reyes 2010 | Low | Low | Low | Low | Low |
| Ristic 2018 | Low | Low | Low | Low | Low |
| Smit 2012 | Low | Low | Low | Low | Low |
| Song 2013 | Low | Low | Low | Low | Low |
| Song 2020 | Low | Low | Low | Low | Low |
| Streng 2018 | Low | Low | Low | Low | Low |
| Wane 2012 | Low | Low | Low | Low | Low |
| Watanabe 2021 | Low | Low | Low | Low | Low |
| Wie 2013 | Low | Low | Low | Low | Low |
| Wong 2014 | Low | Low | Low | Low | Low |
| Wu 2010 | Low | Low | Low | Low | Low |
| Yoon 2021 | Low | Low | Low | Low | Low |
| HPA 2009 | Low | Low | Low | Low | Low |
| Yokomichi 2019 | Low | Low | Low | Low | Low |
| **All-cause mortality** | | | | | |
| Abdalla 2020 | Moderate | Low | Low | Low | Low |
| Guesneau 2021 | Low | Low | Low | Low | Low |
| Ackerson 2019 | Low | Low | Low | Low | Low |
| Adams 2022 | Low | Low | Low | Low | Low |
| Adisasmito 2010 | Low | Low | Low | Low | Low |
| Adlhoch 2023 | Low | Low | Low | Low | Low |
| Adlhoch 2019 | Low | Low | Low | Low | Low |
| Adlhoch 2018 | Moderate | Low | Low | Low | Low |
| Ahout 2018 | Low | Low | Low | Low | Low |
| Akinci 2011 | Low | Low | Low | Low | Low |
| Al Ali 2021 | Low | Low | Low | Low | Low |
| Al Subaie 2012 | Low | Low | Low | Low | Low |
| Al-Abdallat 2016 | Low | Low | Low | Low | Low |
| Al-Awaidy 2015 | Low | Low | Low | Low | Low |
| Al-Baadani 2019 | Low | Low | Low | Low | Low |
| Al-Busaidi 2016 | Moderate | Low | Low | Low | Low |
| Allam 2013 | Low | Low | Low | Low | Low |
| Al-Lawati 2010 | Low | Low | Low | Low | Low |
| Alvarez-Lerma 2017 | Low | Low | Low | Low | Low |
| Amaravathi 2015 | Low | Low | Low | Low | Low |
| Ampofo 2006 | Moderate | Low | Low | Low | Low |
| Andrew 2021 | Low | Low | Low | Low | Low |
| Andrew 2023 | Low | Low | Low | Low | Low |
| Angeles-Sistac 2020 | Low | Low | Low | Low | Low |
| CDC 2010 | Moderate | Low | Low | Low | Low |
| CDESS 2022 | Moderate | Low | Low | Low | Low |
| Ao 2019 | Low | Low | Low | Low | Low |
| Assaf-Casals 2020 | Moderate | Low | Low | Low | Low |
| Auvinen 2022 | Low | Low | Low | Low | Low |
| Aziza 2021 | Low | Low | Low | Low | Low |
| Babamahmoodi 2017 | Moderate | Low | Low | Low | Low |
| Baggett 2012 | Moderate | Low | Low | Low | Low |
| Baigalmaa 2012 | Moderate | Low | Low | Low | Low |
| Baker 2017 | Moderate | Low | Low | Low | Low |
| Barakat 2012 | Moderate | Low | Low | Low | Low |
| Barde 2017 | Low | Low | Low | Low | Low |
| Baselga-Moreno 2019 | Low | Low | Low | Low | Low |
| Bassetti 2019 | Low | Low | Low | Low | Low |
| Begley 2022 | Low | Low | Low | Low | Low |
| Bergmann 2023 | Moderate | Moderate | Low | Low | Moderate |
| Biasco 2022 | Low | Low | Low | Low | Low |
| Bilgin 2023 | Low | Low | Low | Low | Low |
| Birkelo 2021 | Low | Low | Low | Low | Low |
| Chao 2018 | Low | Low | Low | Low | Low |
| Chen 2020 | Low | Low | Low | Low | Low |
| Chien 2022 | Low | Low | Low | Low | Low |
| Choi 2011 | Low | Low | Low | Low | Low |
| Choi 2017 | Low | Low | Low | Low | Low |
| Chorazka 2021 | Moderate | Low | Low | Low | Low |
| Chowell 2012 (a) | Low | Moderate | Low | Low | Low |
| Chowell 2012 (b) | Moderate | Low | Low | Low | Low |
| Chu 2023 | Low | Low | Low | Low | Low |
| Chuang 2012 | Moderate | Moderate | Low | Low | Moderate |
| Chuaychoo 2021 | Low | Low | Low | Low | Low |
| Ciftci 2011 | Low | Low | Low | Low | Low |
| Cobb 2021 | Low | Low | Low | Low | Low |
| Cohen 2014 | Moderate | Low | Low | Low | Low |
| Cohen 2023 | Moderate | Low | Low | Low | Low |
| Collins 2019 | Moderate | Low | Low | Low | Low |
| Cordero 2012 | Low | Low | Low | Low | Low |
| Correia 2010 | Moderate | Low | Low | Low | Low |
| Cost 2011 | Low | Low | Low | Low | Low |
| Coussement 2022 | Low | Low | Low | Low | Low |
| Cox 2012 | Moderate | Low | Low | Low | Low |
| Cui 2010 | Low | Low | Low | Low | Low |
| Cummings 2022 | Moderate | Low | Low | Low | Low |
| Czaja 2019 | Low | Low | Low | Low | Low |
| Da Dalt 2011 | Low | Low | Low | Low | Low |
| DaeiSorkhabi 2022 | Moderate | Low | Low | Low | Low |
| Dao 2010 | Low | Low | Low | Low | Low |
| Davila-Torres 2015 | Moderate | Low | Low | Low | Low |
| Dawood 2014 | Low | Low | Low | Low | Low |
| Dawood 2020 | Low | Low | Low | Low | Low |
| Dawood 2010 | Moderate | Low | Low | Low | Low |
| Dawood 2009 | Low | Low | Low | Low | Low |
| de Morais 2022 | Low | Low | Low | Low | Low |
| Deguchi 2001 | Low | Low | Low | Low | Low |
| Delahoy 2023 | Low | Low | Low | Low | Low |
| Delgado-Sanz 2020 | Moderate | Low | Low | Low | Low |
| Diaz 2012 | Low | Low | Low | Low | Low |
| Dickow 2023 | Low | Low | Low | Low | Low |
| Dolan 2012 | Low | Low | Low | Low | Low |
| Dou 2020 | Low | Low | Low | Low | Low |
| Doyle 2013 | Low | Low | Low | Low | Low |
| Draganescu 2019 | Low | Low | Low | Low | Low |
| Duarte 2009 | Low | Low | Low | Low | Low |
| Dwibedi 2019 | Moderate | Low | Low | Low | Low |
| Dwyer 2017 | Low | Low | Low | Low | Low |
| Ebrahimi 2022 | Low | Low | Low | Low | Low |
| Echevarria-Zuno 2009 | Low | Low | Low | Low | Low |
| Ezzine 2023 | Low | Low | Low | Low | Low |
| Fahim 2021 | Low | Low | Low | Low | Low |
| Fahim 2022 | Low | Low | Low | Low | Low |
| Farias 2010 | Low | Low | Low | Low | Low |
| Feikin 2012 | Moderate | Low | Low | Low | Low |
| Feldstein 2021 | Low | Low | Low | Low | Low |
| Fielding 2009 | Moderate | Low | Low | Low | Low |
| Frohlich 2022 | Low | Low | Low | Low | Low |
| Fu 2019 | Moderate | Low | Low | Low | Low |
| Fuhrman 2011 | Low | Low | Low | Low | Low |
| Fullana Barcelo 2021 | Low | Low | Low | Low | Low |
| Fuller 2022 | Moderate | Low | Low | Low | Low |
| Gachari 2022 | Moderate | Low | Low | Low | Low |
| Gacouin 2020 | Low | High | Low | Low | High |
| Galindo-Fraga 2013 | Moderate | Low | Low | Low | Low |
| Garnacho-Montero 2013 | Low | Low | Low | Low | Low |
| Garnacho-Montero 2018 | Low | Low | Low | Low | Low |
| Geerdes-Fenge 2022 | Low | Low | Low | Low | Low |
| Gefenaite 2018 | Moderate | Low | Low | Low | Low |
| Ghosh 2021 | Moderate | Low | Low | Low | Low |
| Giannella 2012 | Low | Low | Low | Low | Low |
| Gilca 2021 | Low | Low | Low | Low | Low |
| Gioula 2010 | Low | Low | Low | Low | Low |
| Godoy 2018 | Low | Moderate | Low | Low | Low |
| Golagana 2023 | Low | Low | Low | Low | Low |
| Gouya 2011 | Moderate | Low | Low | Low | Low |
| Groeneveld 2020 | Low | Moderate | Low | Low | Low |
| Gubbels 2013 | Low | Low | Low | Low | Low |
| Gutierrez-Cuadra 2012 | Low | Low | Low | Low | Low |
| Gutierrez-Pizarraya 2012 | Low | Low | Low | Low | Low |
| Hagerman 2015 | Low | Low | Low | Low | Low |
| Hagiwara 2022 | Low | Low | Low | Low | Low |
| Han 2011 | Moderate | Low | Low | Low | Low |
| Han 2021 | Low | Low | Low | Low | Low |
| Harris 2019 | Low | Low | Low | Low | Low |
| Hedberg 2022 | Low | Low | Low | Low | Low |
| Helferty 2010 | Low | Low | Low | Low | Low |
| Herbstreit 2021 | Low | Low | Low | Low | Low |
| Hernandez-Garces 2021 | Low | Low | Low | Low | Low |
| Hernandez-Cardenas 2016 | Low | Low | Low | Low | Low |
| Hernu 2021 | Low | Low | Low | Low | Low |
| Heyd 2017 | Low | High | Low | Low | High |
| Hiba 2011 | Low | Low | Low | Low | Low |
| Hobbs 2019 | Low | Low | Low | Low | Low |
| Hong 2021 | Low | Low | Low | Low | Low |
| Hsieh 2018 | Low | Low | Low | Low | Low |
| Hsing 2022 | Low | Low | Low | Low | Low |
| Hu 2023 | Low | Low | Low | Low | Low |
| Huai 2017 | Low | Low | Low | Low | Low |
| Huang 2017 | Low | Low | Low | Low | Low |
| Jamoussi 2022 | Low | Low | Low | Low | Low |
| Jorda 2023 | Low | Low | Low | Low | Low |
| Kandeel 2016 | Low | Low | Low | Low | Low |
| Kappagoda 2000 | Moderate | Low | Low | Low | Low |
| Karolyi 2021 | Low | Low | Low | Low | Low |
| Karolyi 2019 | Low | Low | Low | Low | Low |
| Katzen 2019 | Low | Low | Low | Low | Low |
| Khandaker 2014 | Low | Low | Low | Low | Low |
| Kim 2023 | Low | Low | Low | Low | Low |
| Kini 2018 | Low | Low | Low | Low | Low |
| Kohlmaier 2020 | Low | Low | Low | Low | Low |
| Kojicic 2012 | Low | Low | Low | Low | Low |
| Kok 2013 | Low | Low | Low | Low | Low |
| Kovacevic 2020 | Low | Low | Low | Low | Low |
| Kraft 2012 | Low | Low | Low | Low | Low |
| Kuchar 2013 | Moderate | Low | Low | Low | Low |
| Kumar 2010 | Low | Low | Low | Low | Low |
| Kumar 2016 | Low | Low | Low | Low | Low |
| Kusznierz 2017 | Moderate | Moderate | Low | Low | Moderate |
| Kwon 2017 | Low | Low | Low | Low | Low |
| Lalueza 2020 | Low | Low | Low | Low | Low |
| Laris-Gonzalez 2021 | Low | Low | Low | Low | Low |
| Lee 2020 | Low | Low | Low | Low | Low |
| Lee 2011 | Low | Moderate | Low | Low | Low |
| Lehners 2013 | Low | Low | Low | Low | Low |
| Lenzi 2012 | Low | Low | Low | Low | Low |
| Leung 2014 | Low | Low | Low | Low | Low |
| Li 2021 | Low | Low | Low | Low | Low |
| Li 2011 | Low | Low | Low | Low | Low |
| Liem 2009 | Low | Low | Low | Low | Low |
| Lim 2015 | Low | Low | Low | Low | Low |
| Lim 2010 | Low | Low | Low | Low | Low |
| Lindblade 2010 | Low | Low | Low | Low | Low |
| Liu 2020 | Low | Low | Low | Low | Low |
| Lo 2013 | Moderate | Low | Low | Low | Low |
| Lopez Montesinos 2022 | Low | Low | Low | Low | Low |
| Lopez-Delgado 2013 | Low | Low | Low | Low | Low |
| Lopez-Medina 2012 | Low | Low | Low | Low | Low |
| Louie 2009 | Low | Low | Low | Low | Low |
| Louie 2013 | Low | Low | Low | Low | Low |
| Louriz 2010 | Low | Low | Low | Low | Low |
| Lovato-Salas 2010 | Moderate | Low | Low | Low | Low |
| Lucker 2011 | Low | Low | Low | Low | Low |
| Lynfield 2014 | Low | Low | Low | Low | Low |
| Lytras 2020 | Moderate | Low | Low | Low | Low |
| Mabayoje 2021 | Moderate | Low | Low | Low | Low |
| Malhotra 2016 | Moderate | Low | Low | Low | Low |
| Mansour 2012 | Low | Low | Low | Low | Low |
| Mao 2014 | Low | Low | Low | Low | Low |
| Marbus 2020 | Low | Low | Low | Low | Low |
| Martin 2013 | Low | Low | Low | Low | Low |
| Martinez-Briseno 2016 | Low | Low | Low | Low | Low |
| Martin-Loeches 2011 | Low | Low | Low | Low | Low |
| Martin-Loeches 2017 | Low | Low | Low | Low | Low |
| Martin-Loeches 2019 | Low | Low | Low | Low | Low |
| Maruyama 2016 | Low | Low | Low | Low | Low |
| Mattila 2020 | Low | Low | Low | Low | Low |
| McGeer 2007 | Low | Low | Low | Low | Low |
| McGrath 2023 | Low | Low | Low | Low | Low |
| McNeil 2014 | Low | Low | Low | Low | Low |
| McRae 2022 | Low | Low | Low | Low | Low |
| Mehta 2013 | Low | Low | Low | Low | Low |
| Mehta 2016 | Moderate | Low | Low | Low | Low |
| Mendez-Dominguez 2019 | Low | Low | Low | Low | Low |
| Miller 2010 | Low | Low | Low | Low | Low |
| Minney-Smith 2019 | Moderate | Low | Low | Low | Low |
| Miron 2021 | Low | Low | Low | Low | Low |
| Modemann 2022 | Low | Low | Low | Low | Low |
| Montes 2005 | Moderate | Low | Low | Low | Low |
| Moreno 2021 | Low | Low | Low | Low | Low |
| Murti 2018 | Moderate | Low | Low | Low | Low |
| Muscatello 2014 | Low | Low | Low | Low | Low |
| Nandhini 2015 | Low | Low | Low | Low | Low |
| Nasir 2021 | Low | Low | Low | Low | Low |
| Nation 2021 | Moderate | High | Low | Low | High |
| Nichols 2018 | Low | Low | Low | Low | Low |
| Nickel 2011 | Low | Low | Low | Low | Low |
| Nieto-Guevara 2011 | Low | Low | Low | Low | Low |
| Nordenskjold 2022 | Low | Low | Low | Low | Low |
| Oliva 2018 | Moderate | Low | Low | Low | Low |
| Ono 2016 | Low | Low | Low | Low | Low |
| Pang 2021 | Low | Low | Low | Low | Low |
| Parisi 2023 | Low | Low | Low | Low | Low |
| Pascale 2021 | Low | Moderate | Low | Low | Low |
| Pedroni 2010 | Low | Low | Low | Low | Low |
| Phung 2011 | Low | Low | Low | Low | Low |
| Plumb 2021 | Low | Low | Low | Low | Low |
| Puig-Barbera 2014 (a) | Low | Low | Low | Low | Low |
| Puig-Barbera 2016 | Low | Low | Low | Low | Low |
| Puig-Barbera 2014 (b) | Low | Low | Low | Low | Low |
| Punpanich 2014 | Low | Low | Low | Low | Low |
| Raff 2022 | Low | Low | Low | Low | Low |
| Samransamruajkit 2008 | Low | Low | Low | Low | Low |
| Saroch 2018 | Low | Low | Low | Low | Low |
| Satterwhite 2010 | Low | Low | Low | Low | Low |
| Schauwvlieghe 2018 | Low | Low | Low | Low | Low |
| Schober 2023 | Low | Low | Low | Low | Low |
| SerpaNeto 2021 | Low | Low | Low | Low | Low |
| Shalabi 2022 | Low | Low | Low | Low | Low |
| Shannon 2022 | Low | Low | Low | Low | Low |
| Sharma 2020 | Low | Low | Low | Low | Low |
| Sherban 2023 | Low | Low | Low | Low | Low |
| Shimada 2015 | Low | Low | Low | Low | Low |
| Shusterman 2023 | Low | Low | Low | Low | Low |
| Susilarini 2018 | Low | Low | Low | Low | Low |
| Talbot 2021 | Low | Low | Low | Low | Low |
| Tamma 2010 | Low | Low | Low | Low | Low |
| Taniguchi 2022 | Low | Low | Low | Low | Low |
| Taylor 2016 | Low | Low | Low | Low | Low |
| Tempia 2017 | Low | Low | Low | Low | Low |
| Teng 2019 | Low | Low | Low | Low | Low |
| Teros-Jaakkola 2019 | Low | Low | Low | Low | Low |
| Thangaraj 2023 | Low | Low | Low | Low | Low |
| Thelen 2021 | Low | Low | Low | Low | Low |
| To 2010 | Low | Low | Low | Low | Low |
| Torres 2010 | Low | Low | Low | Low | Low |
| Tramuto 2011 | Low | Low | Low | Low | Low |
| Tran 2016 | Moderate | Low | Low | Low | Low |
| Truelove 2011 | Low | Low | Low | Low | Low |
| Tsukui 2012 | Low | Low | Low | Low | Low |
| Unal 2023 | Low | Low | Low | Low | Low |
| van't Klooster 2010 | Moderate | Low | Low | Low | Low |
| Vandroux 2019 | Low | Low | Low | Low | Low |
| Viasus 2012 | Low | Low | Low | Low | Low |
| vonBaum 2011 | Low | Low | Low | Low | Low |
| Borgatta 2012 | Low | Low | Low | Low | Low |
| Borja-Aburto 2012 | Low | Low | Low | Low | Low |
| Bouneb 2018 | Low | Low | Low | Low | Low |
| Bunthi 2013 | Low | Low | Low | Low | Low |
| Burkert 2022 | Low | Low | Low | Low | Low |
| Burton 2008 | Low | Low | Low | Low | Low |
| Campbell 2020 | Low | Low | Low | Low | Low |
| Campbell 2021 | Low | Low | Low | Low | Low |
| Campbell 2011 | Low | Low | Low | Low | Low |
| Casas-Aparicio 2018 | Low | Low | Low | Low | Low |
| Castillo-Palencia 2012 | Low | Low | Low | Low | Low |
| Chakhunashvili 2018 | Low | Low | Low | Low | Low |
| Chan 2015 | Low | Low | Low | Low | Low |
| Chan 2011 | Low | Low | Low | Low | Low |
| Chan 2017 | Low | Low | Low | Low | Low |
| Chatterjee 2020 | Low | Low | Low | Low | Low |
| Chaves 2013 | Low | Low | Low | Low | Low |
| Chaves 2023 | Low | Low | Low | Low | Low |
| Chaves 2014 | Low | Low | Low | Low | Low |
| Chen 2021 | Low | Low | Low | Low | Low |
| Cheng 2015 | Low | Low | Low | Low | Low |
| Rao 2023 | Low | Low | Low | Low | Low |
| Reacher 2019 | Low | Low | Low | Low | Low |
| Reed 2014 | Low | Low | Low | Low | Low |
| Regan 2023 | Low | Low | Low | Low | Low |
| Reyes 2007 | Low | Low | Low | Low | Low |
| Reyes 2010 | Low | Low | Low | Low | Low |
| Ristic 2018 | Low | Low | Low | Low | Low |
| Rodriguez 2011 | Low | Low | Low | Low | Low |
| Roedl 2021 | Low | Low | Low | Low | Low |
| Rosler 2021 | Low | Low | Low | Low | Low |
| Rothman 2023 | Low | Low | Low | Low | Low |
| Rovina 2014 | Low | Low | Low | Low | Low |
| Rozencwajg 2018 | Low | Low | Low | Low | Low |
| Smiechowicz 2021 | Low | Low | Low | Low | Low |
| Smit 2012 | Low | Low | Low | Low | Low |
| Snacken 2012 | Low | Low | Low | Low | Low |
| Sohn 2013 | Low | Low | Low | Low | Low |
| Soldevila 2020 | Low | Low | Low | Low | Low |
| Song 2013 | Low | Low | Low | Low | Low |
| Song 2020 | Low | Low | Low | Low | Low |
| Soyemi 2014 | Low | Low | Low | Low | Low |
| Stein 2010 | Low | Low | Low | Low | Low |
| Streng 2011 | Low | Low | Low | Low | Low |
| Subramony 2010 | Low | Low | Low | Low | Low |
| Wallemacq 2022 | Low | Low | Low | Low | Low |
| Wane 2012 | Low | Low | Low | Low | Low |
| Watanabe 2021 | Low | Low | Low | Low | Low |
| Webb 2011 | Low | Low | Low | Low | Low |
| Wei 2021 | Low | Low | Low | Low | Low |
| Wie 2013 | Low | Low | Low | Low | Low |
| Wong 2014 | Low | Low | Low | Low | Low |
| Wong 2016 | Low | Low | Low | Low | Low |
| Wongwiwatwaitaya 2014 | Low | Low | Low | Low | Low |
| Wu 2010 | Low | Low | Low | Low | Low |
| Xiao 2015 | Low | Low | Low | Low | Low |
| Xie 2020 | Low | Low | Low | Low | Low |
| Xu 2013 | Low | Low | Low | Low | Low |
| Yang 2010 | Low | Low | Low | Low | Low |
| Yang 2014 | Low | Low | Low | Low | Low |
| Yang 2017 | Low | Low | Low | Low | Low |
| Yen 2012 | Low | Low | Low | Low | Low |
| Yi 2023 | Low | Low | Low | Low | Low |
| Ylipalosaari 2017 | Low | Low | Low | Low | Low |
| Yoon 2021 | Low | Low | Low | Low | Low |
| Youngs 2019 | Low | Low | Low | Low | Low |
| Zhang 2012 | Low | Low | Low | Low | Low |
| Zhang 2018 | Low | Low | Low | Low | Low |
| Zheng 2020 | Low | Low | Low | Low | Low |
| Zogheib 2018 | Low | Low | Low | Low | Low |
| HPA 2009 | Low | Low | Low | Low | Low |
| Cowling 2013 | Low | Low | Low | Low | Low |
| Jiang 2017 | Low | Low | Low | Low | Low |
| Wang 2017 | Low | Low | Low | Low | Low |

### Appendix 5. Meta-analysis of hospitalization rate in seasonal or pandemic influenza

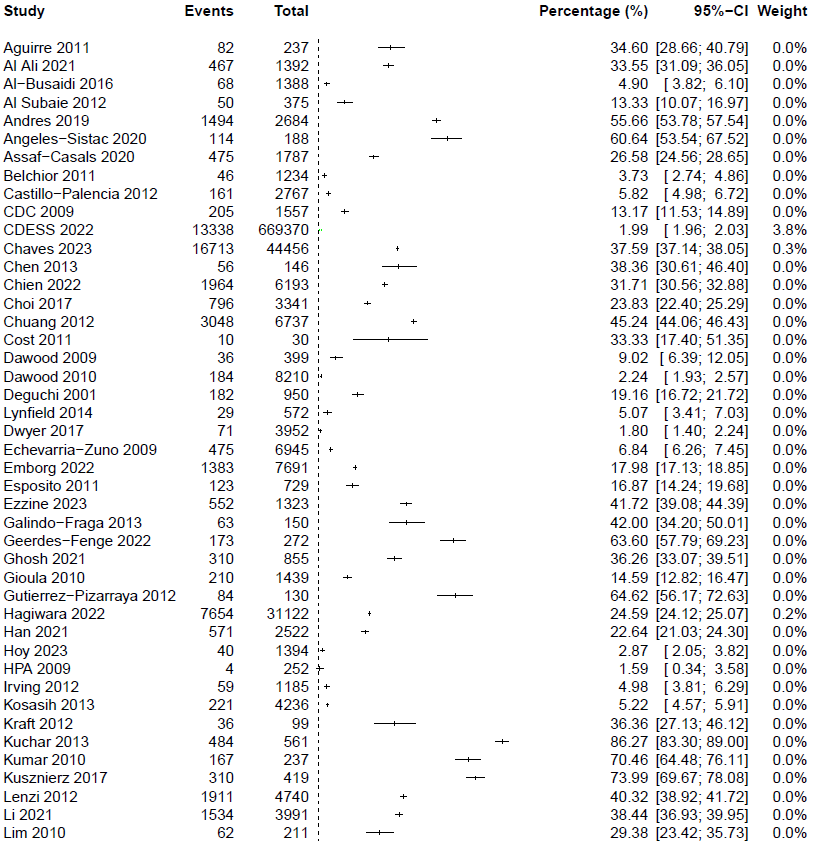

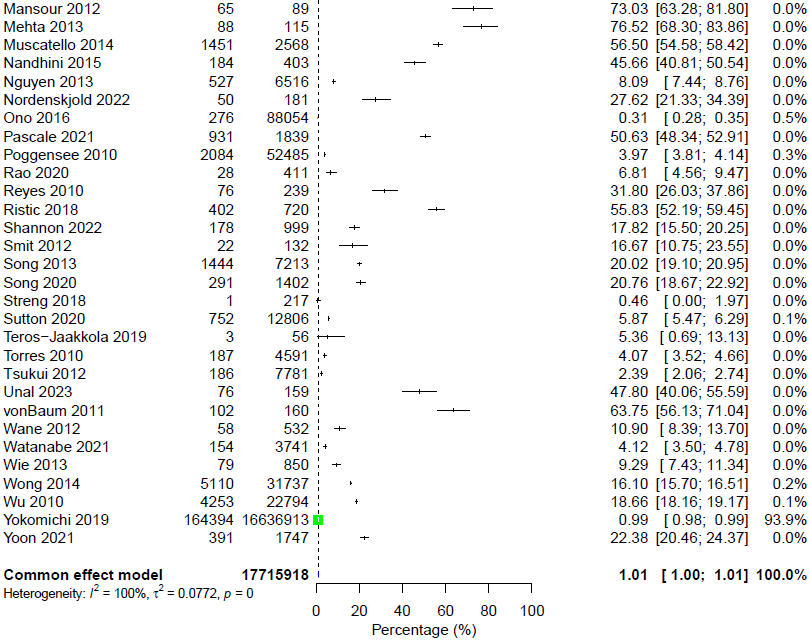

### Appendix 6. Subgroup analysis of hospitalization rate in seasonal or pandemic influenza by age

#### 6.1. Within-study subgroup analysis hospitalization rate in seasonal or pandemic influenza by age

0-14 years vs 15-64 years

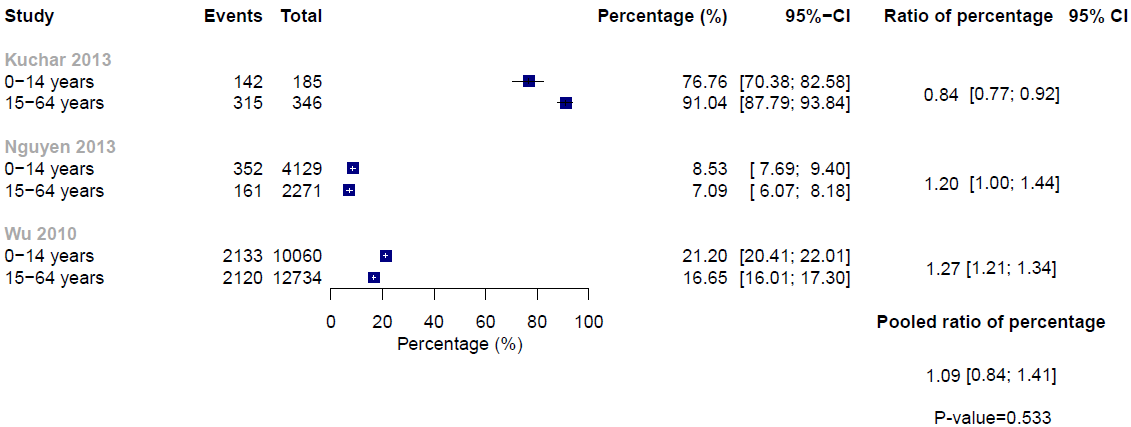

0-14 years vs ≥65 years
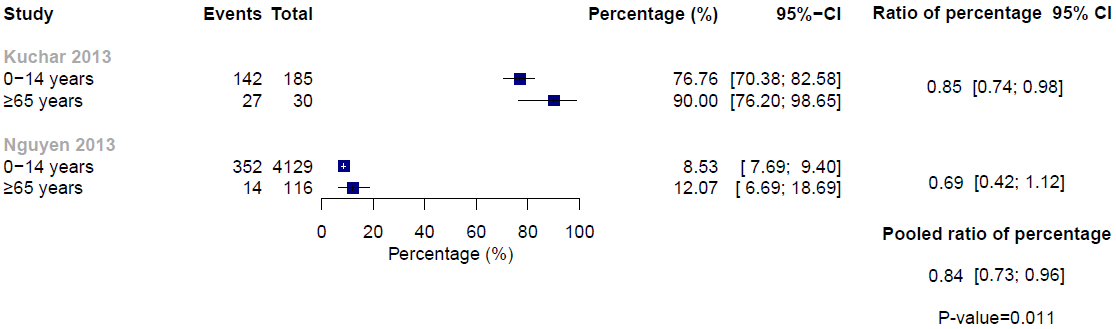

15-64 years vs ≥65 years

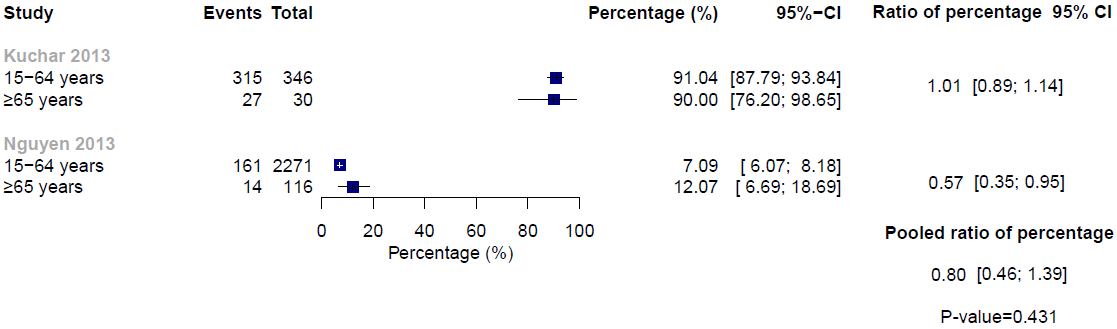

#### 6.2. Between-study subgroup analysis of hospitalization rate in seasonal or pandemic influenza by age

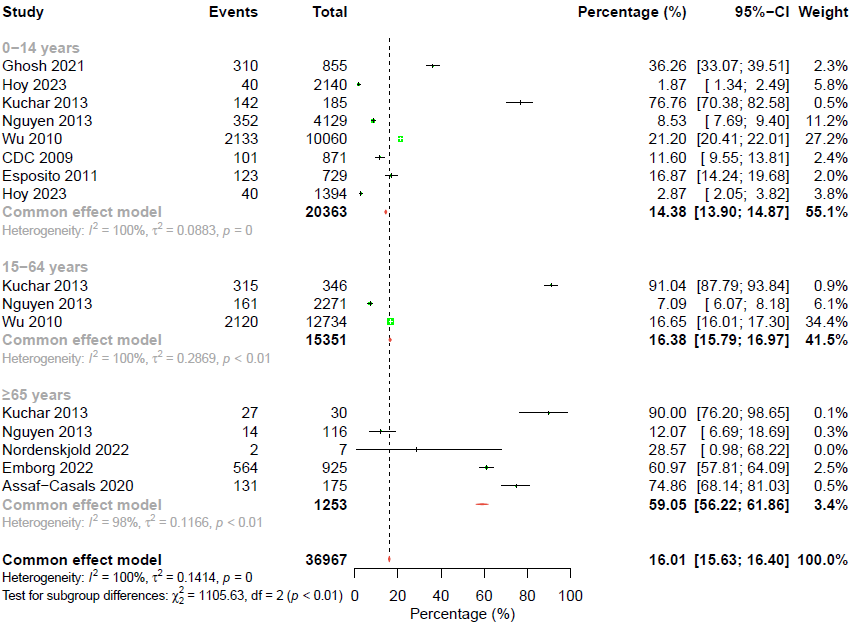

### Appendix 7. Credibility assessment of subgroup analyses for hospitalization rate in seasonal or pandemic influenza

| **ICEMAN item** | **By countries** | **By influenza etiology** | **By chronic comorbidities** | **By age (0-14 years vs ≥65 years)** |
| --- | --- | --- | --- | --- |
| Is the analysis of effect modification based on comparison within rather than between studies? | Completely between | Mostly between | Mostly between | Completely within |
| For within-study comparisons, is the effect modification similar from study to study? | Not applicable | Not applicable | Not applicable | Definitely similar |
| For between-study comparisons, is the number of studies large? | Large | Large | Very small | Not applicable |
| Was the direction of effect modification correctly hypothesised a priori? | Definitely yes | Definitely yes | Definitely yes | Definitely yes |
| Does a test for interaction suggest that chance is an unlikely explanation of the apparent effect modification? | Chance an unlikely explanation | Chance an unlikely explanation | Chance an unlikely explanation | Chance a likely explanation |
| Did the authors test only a small number of effect modifiers or consider the number in their statistical analysis? | Probably no | Probably no | Probably no | Probably no |
| Did the authors use a random effects model? | Definitely no | Definitely no | Definitely no | Definitely yes |
| If the effect modifier is a continuous variable, were arbitrary cut points avoided? | Not applicable | Not applicable | Not applicable | Not applicable |
| Are there any additional considerations that may increase or decrease credibility? | Not applicable | Not applicable | Not applicable | Not applicable |
| How would you rate the overall credibility of the proposed effect modification? | Low credibility | Low credibility | Low credibility | Low credibility |

### Appendix 8. Subgroup analysis of hospitalization rate in seasonal or pandemic influenza by vaccination status

#### 8.1. Within-study subgroup analysis hospitalization rate in seasonal or pandemic influenza by vaccination status

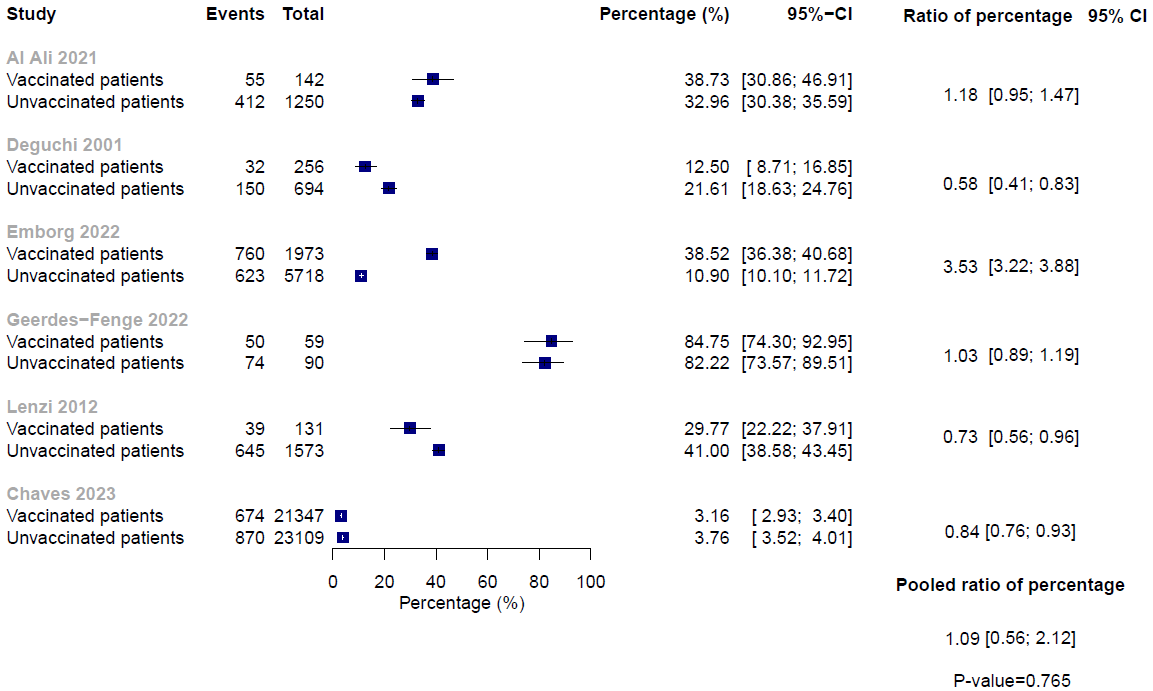

#### 8.2. Between-study subgroup analysis hospitalization rate in seasonal or pandemic influenza by vaccination status

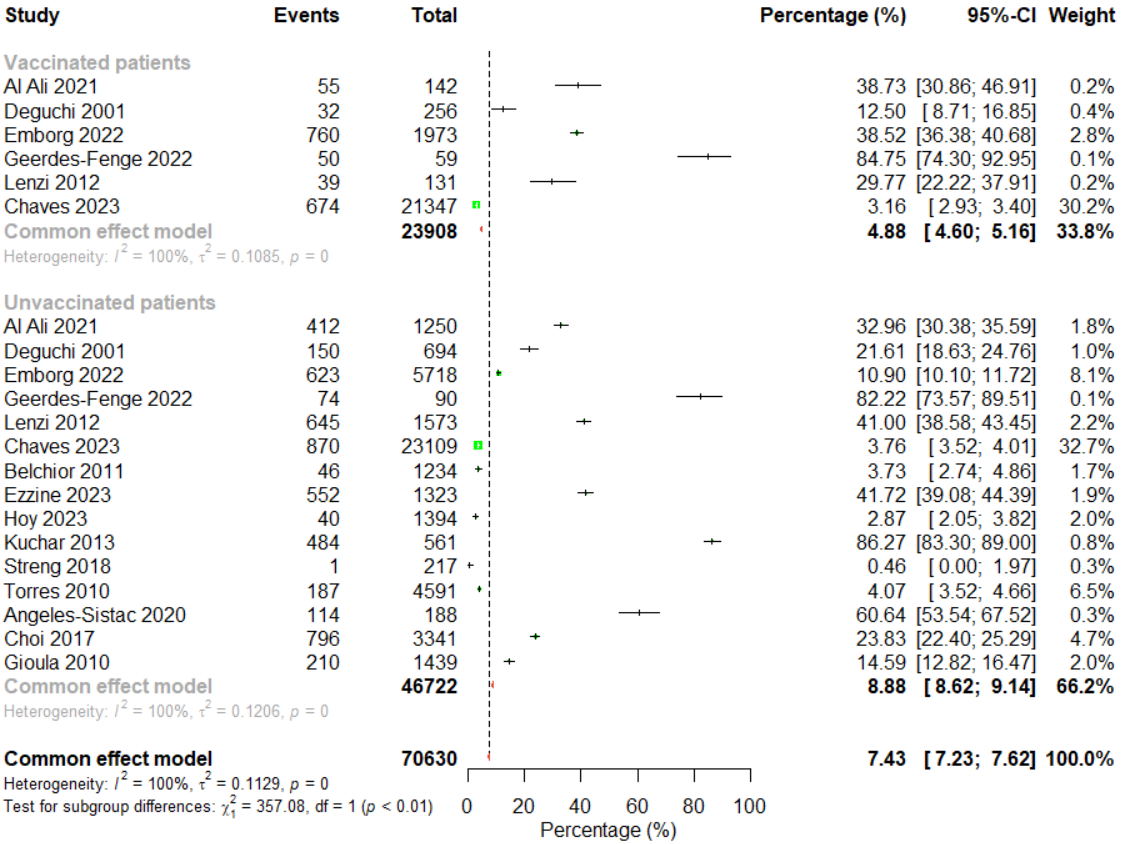

### Appendix 9. Between-study subgroup analysis of hospitalization rate in seasonal or pandemic influenza by countries

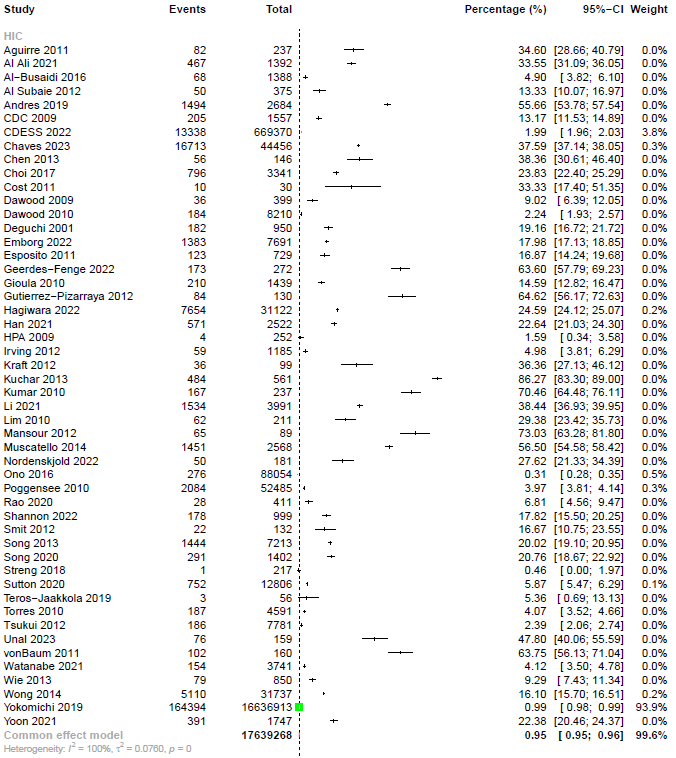

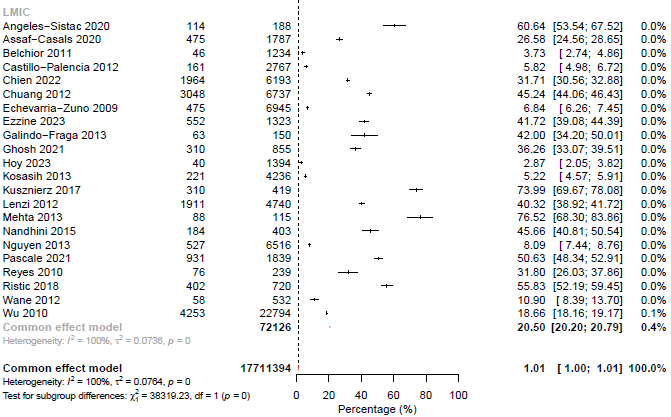

### Appendix 10. Between-study subgroup analysis of hospitalization rate in seasonal or pandemic influenza by influenza etiology

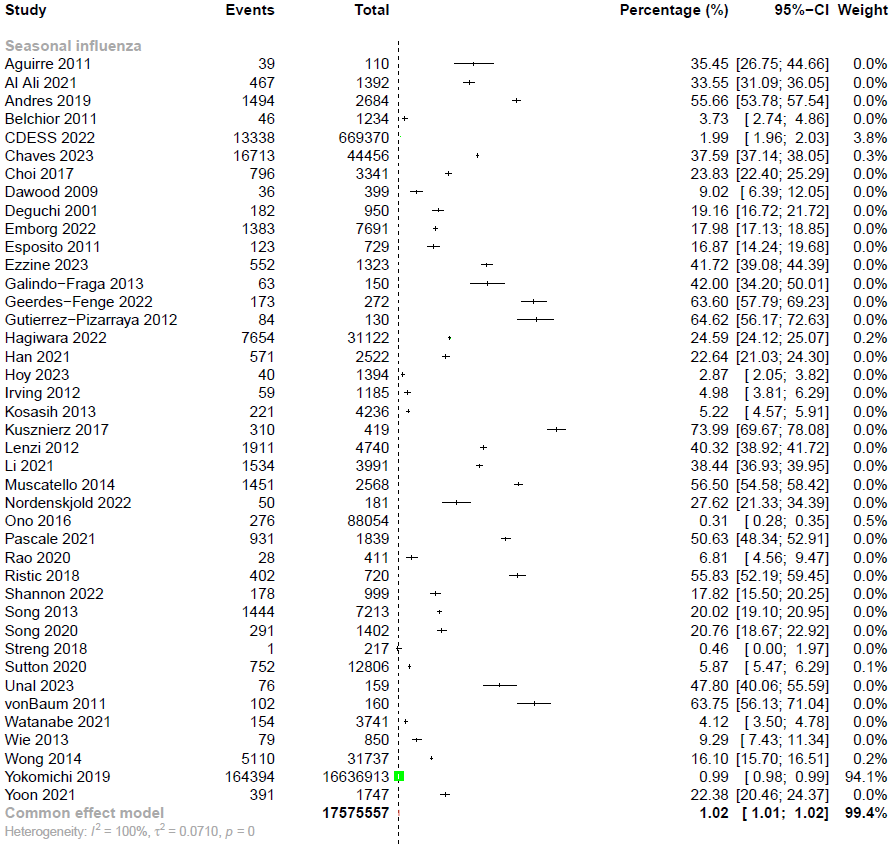

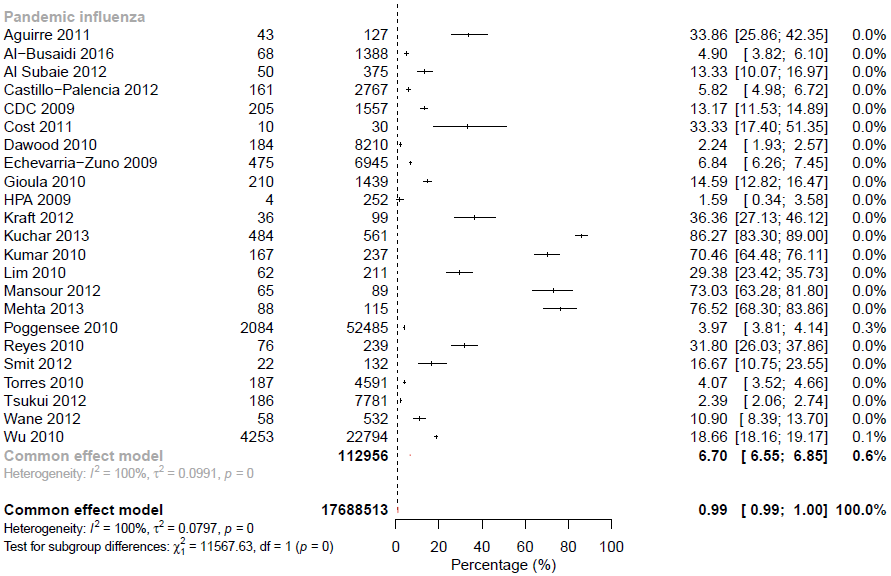

### Appendix 11. Between-study subgroup analysis of hospitalization rate in seasonal or pandemic influenza by chronic comorbidities

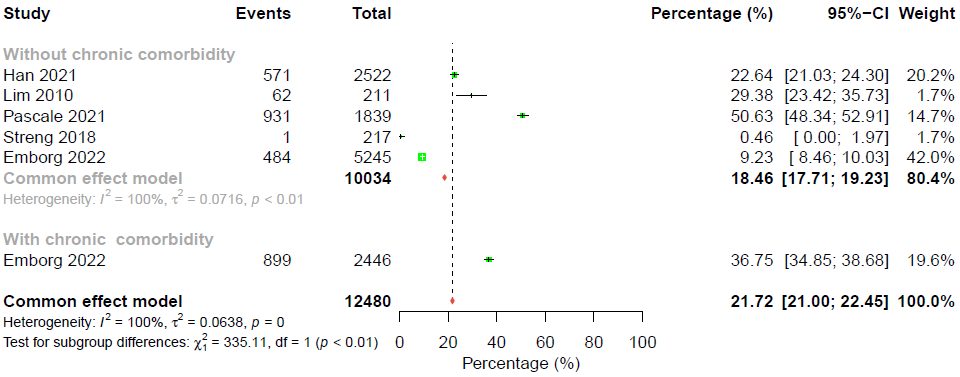

### Appendix 12. Sensitivity analysis only including studies with low risk of bias for hospitalization rate in seasonal or pandemic influenza

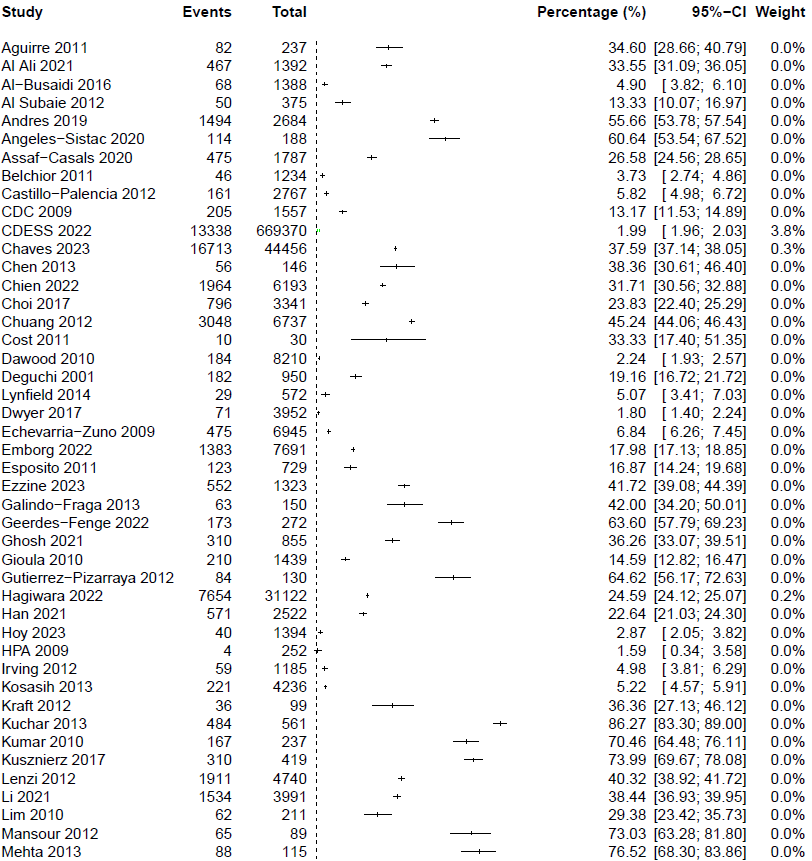

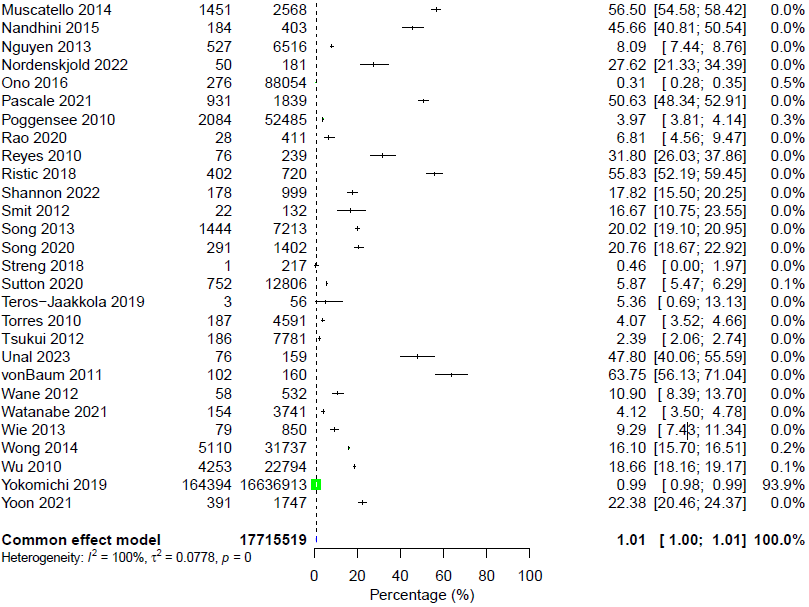

### Appendix 13. Meta-analyses of hospitalization rate in zoonotic influenza

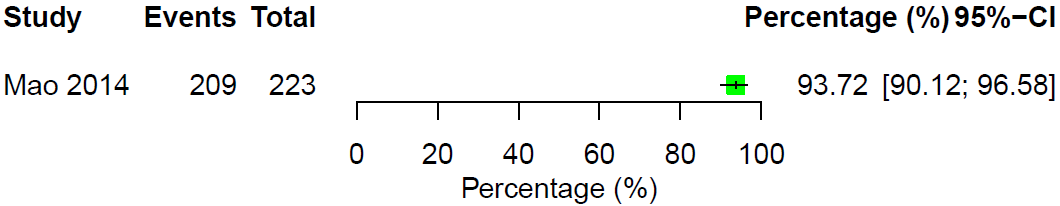

### Appendix 14. Meta-analyses of all-cause mortality rate in seasonal or pandemic influenza

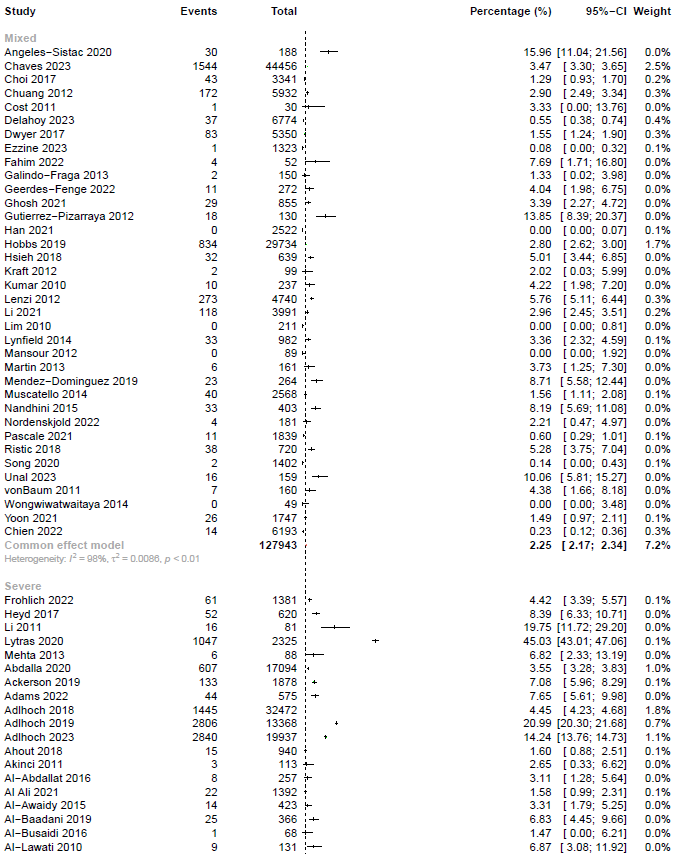

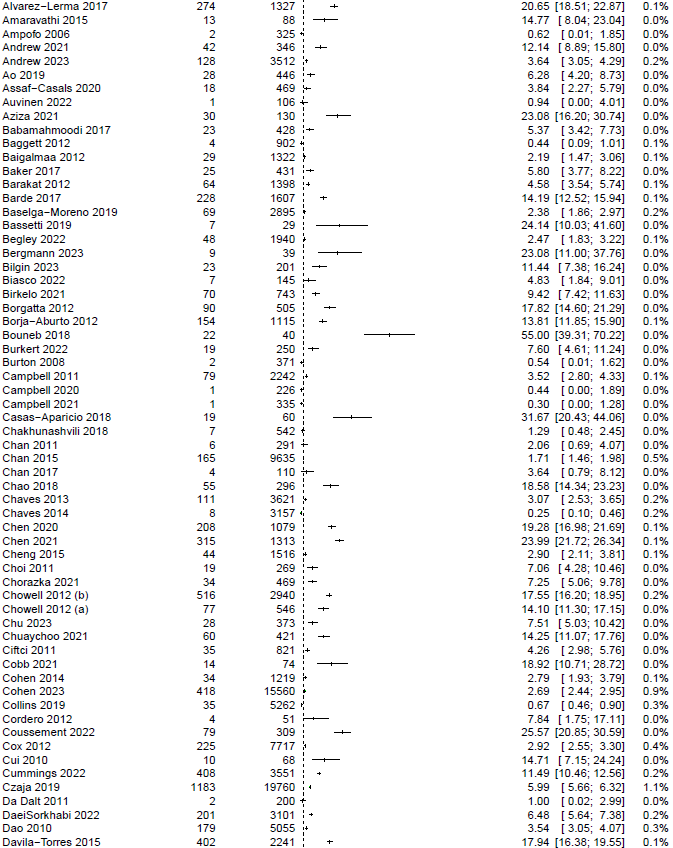

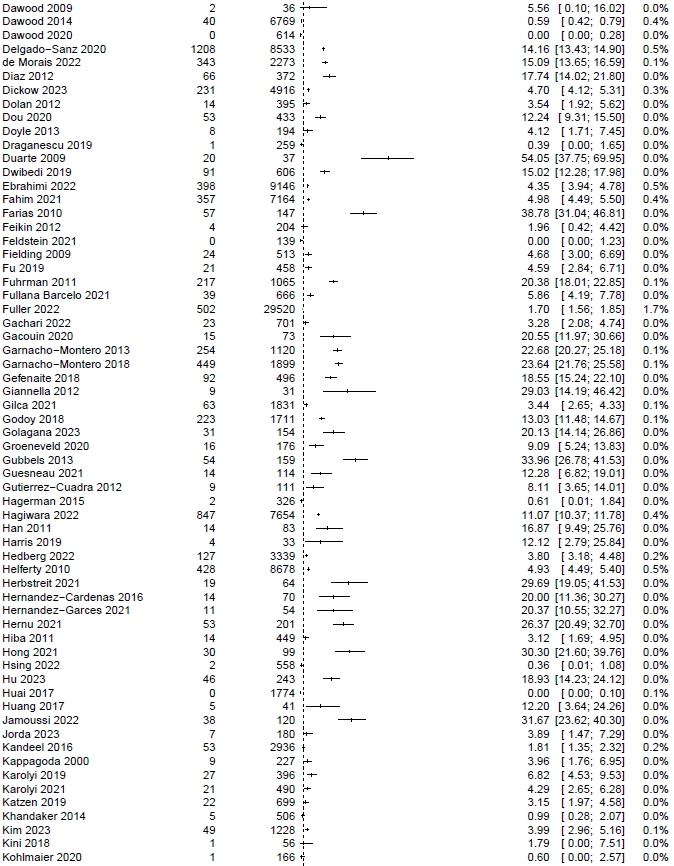

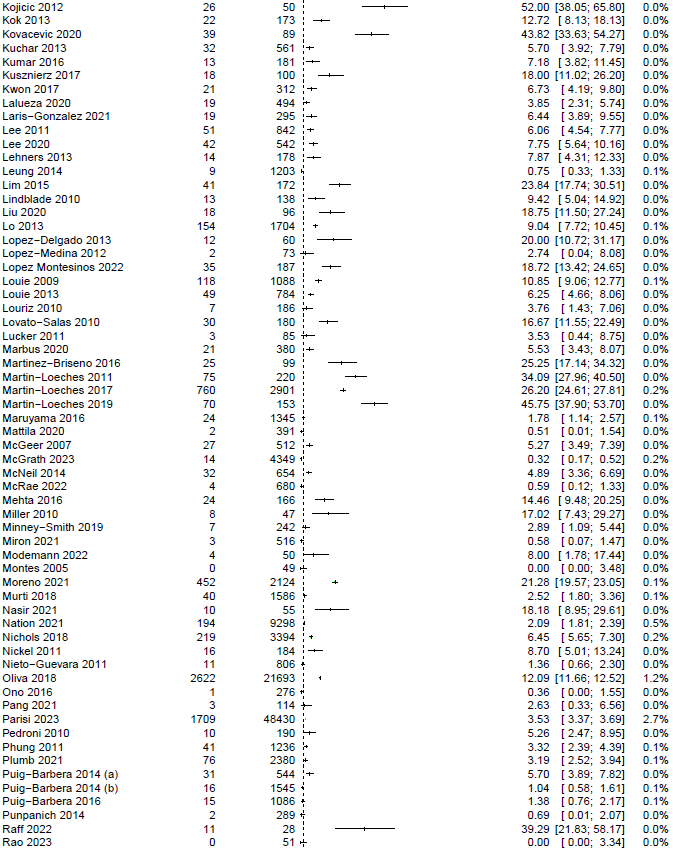

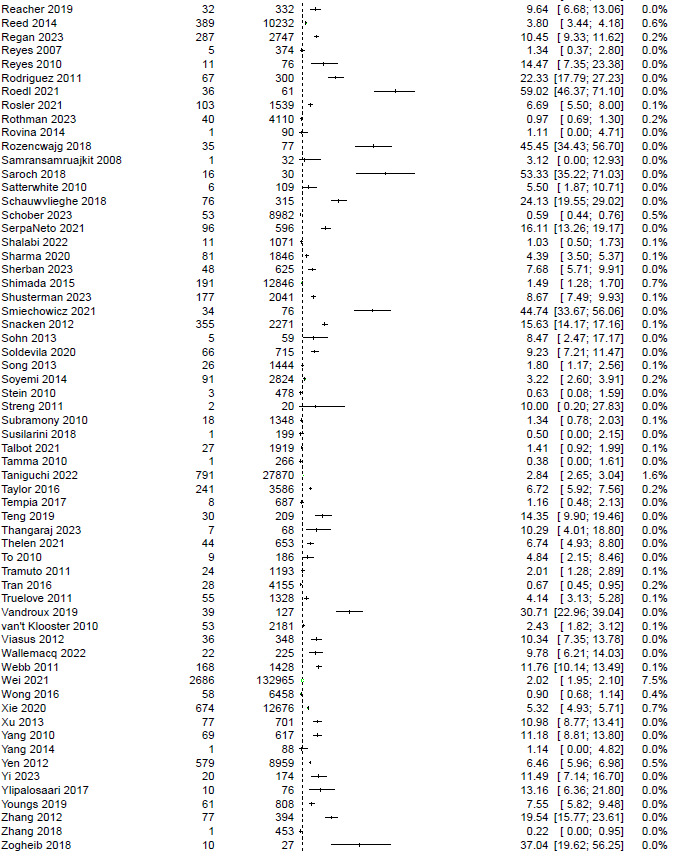

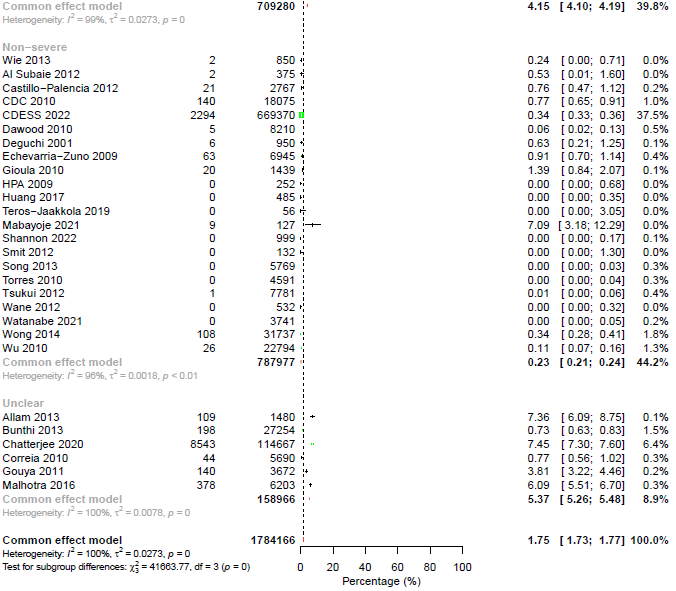

### Appendix 15. Meta-analyses of all-cause mortality rate in non-severe seasonal or pandemic influenza

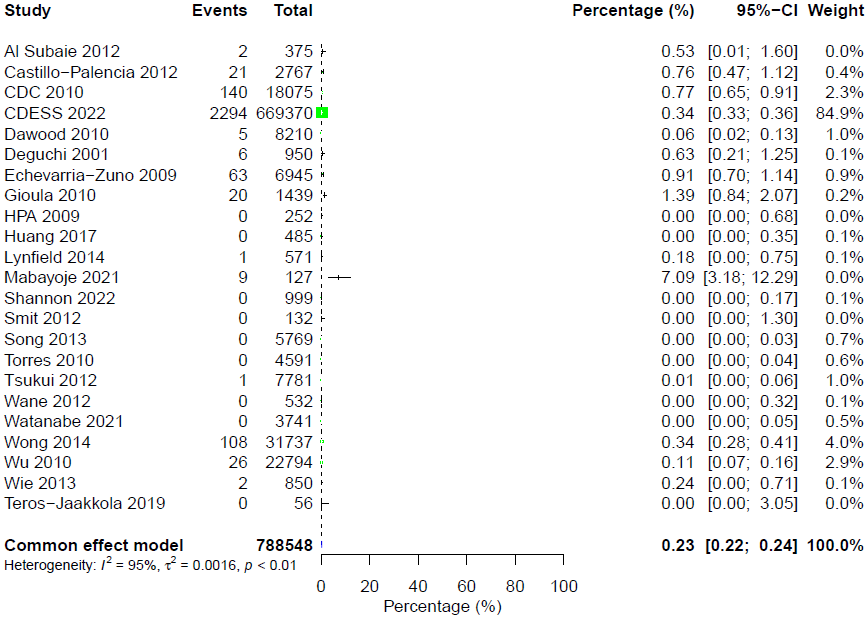

### Appendix 16. Between-study subgroup analysis of all-cause mortality rate in non-severe seasonal or pandemic influenza by countries

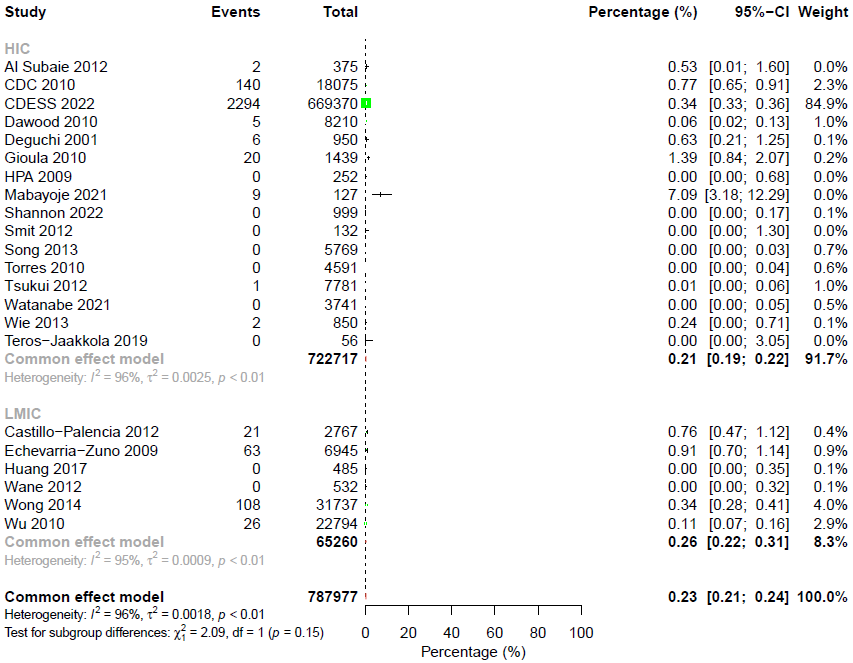

### Appendix 17. Between-study subgroup analysis of all-cause mortality rate in non-severe seasonal or pandemic influenza by influenza etiology

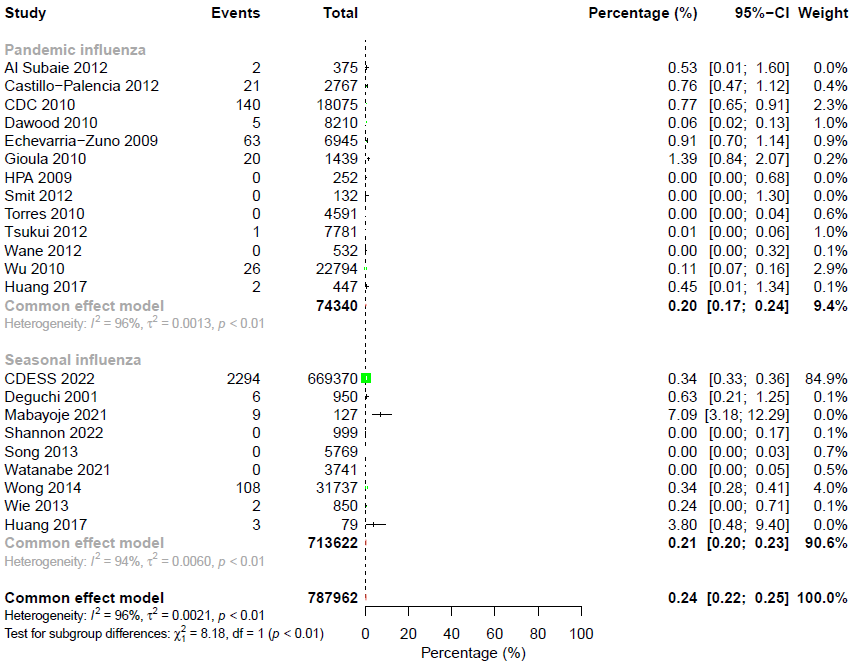

### Appendix 18. Credibility assessment of subgroup analyses for all-cause mortality rate in non-severe seasonal or pandemic influenza

| **ICEMAN item** | **By influenza etiology** |
| --- | --- |
| Is the analysis of effect modification based on comparison within rather than between studies? | Mostly between |
| For within-study comparisons, is the effect modification similar from study to study? | Not applicable |
| For between-study comparisons, is the number of studies large? | Rather large |
| Was the direction of effect modification correctly hypothesised a priori? | Definitely no |
| Does a test for interaction suggest that chance is an unlikely explanation of the apparent effect modification? | Chance an unlikely explanation |
| Did the authors test only a small number of effect modifiers or consider the number in their statistical analysis? | Probably no |
| Did the authors use a random effects model? | Definitely no |
| If the effect modifier is a continuous variable, were arbitrary cut points avoided? | Not applicable |
| Are there any additional considerations that may increase or decrease credibility? | Not applicable |
| How would you rate the overall credibility of the proposed effect modification? | Very low credibility |

### Appendix 19. Meta-analyses of all-cause mortality rate in severe seasonal or pandemic influenza

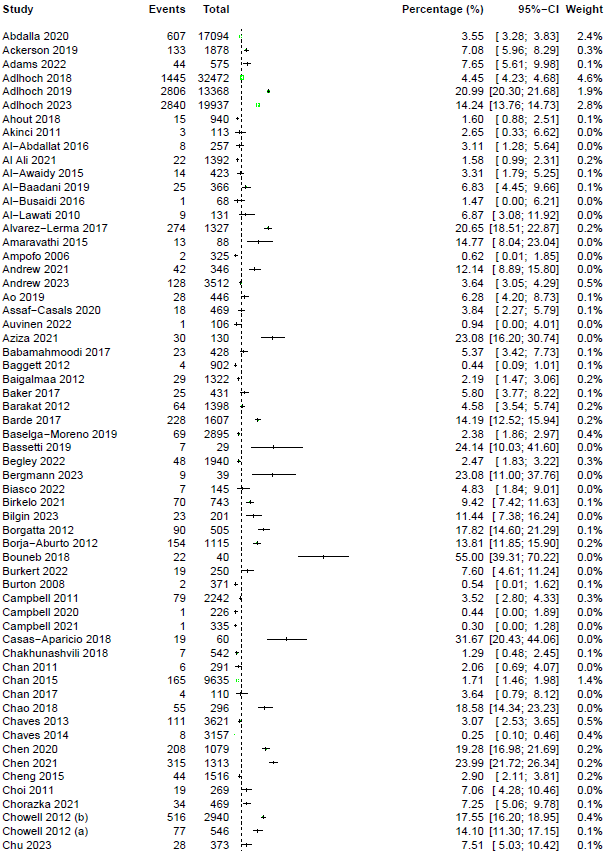

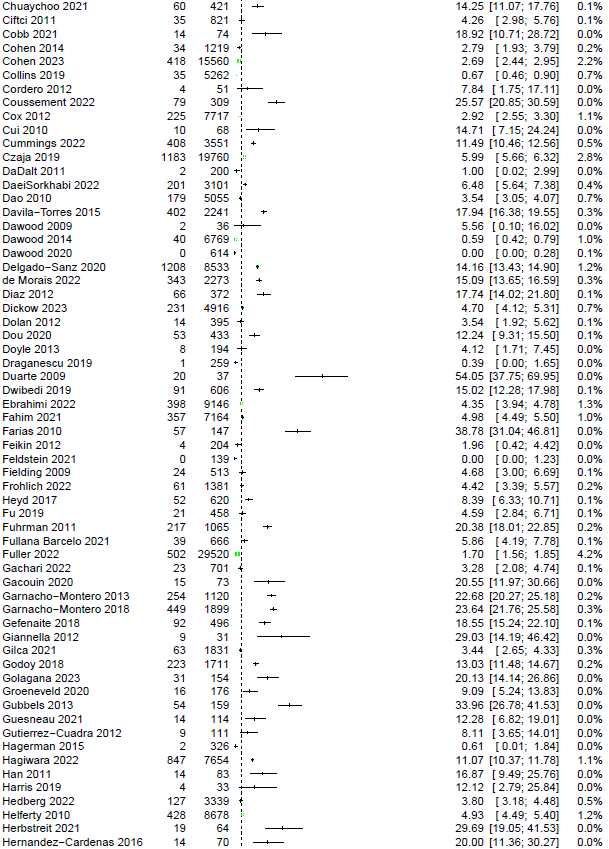

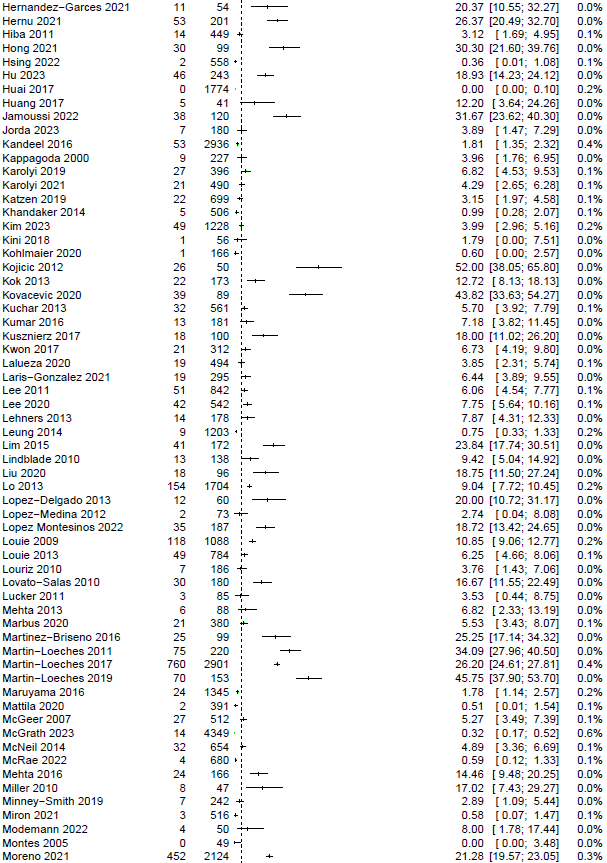

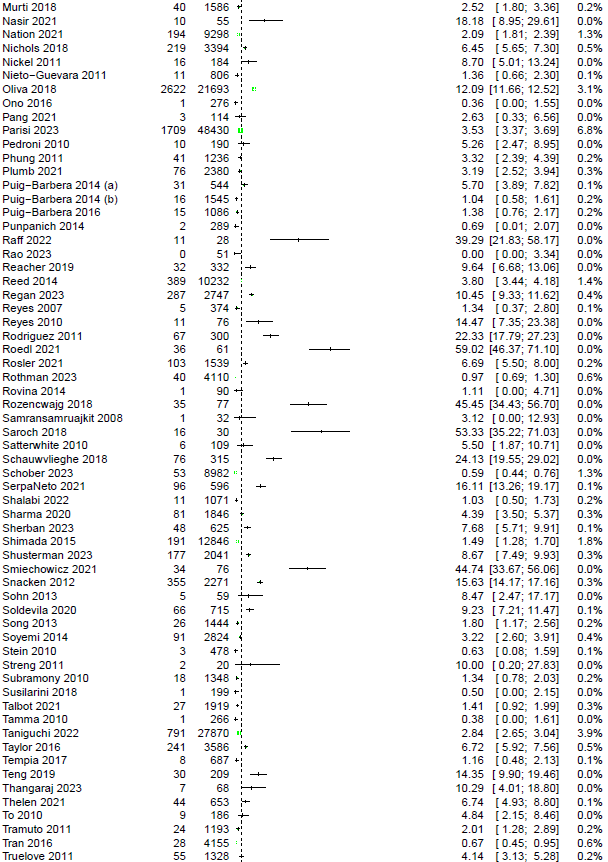

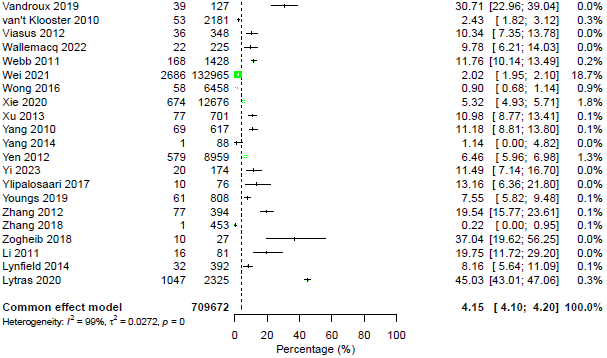

### Appendix 20. Subgroup analysis of all-cause mortality rate in severe seasonal or pandemic influenza by influenza etiology

#### 20.1. Within-study subgroup analysis of all-cause mortality rate in severe seasonal or pandemic influenza by influenza etiology

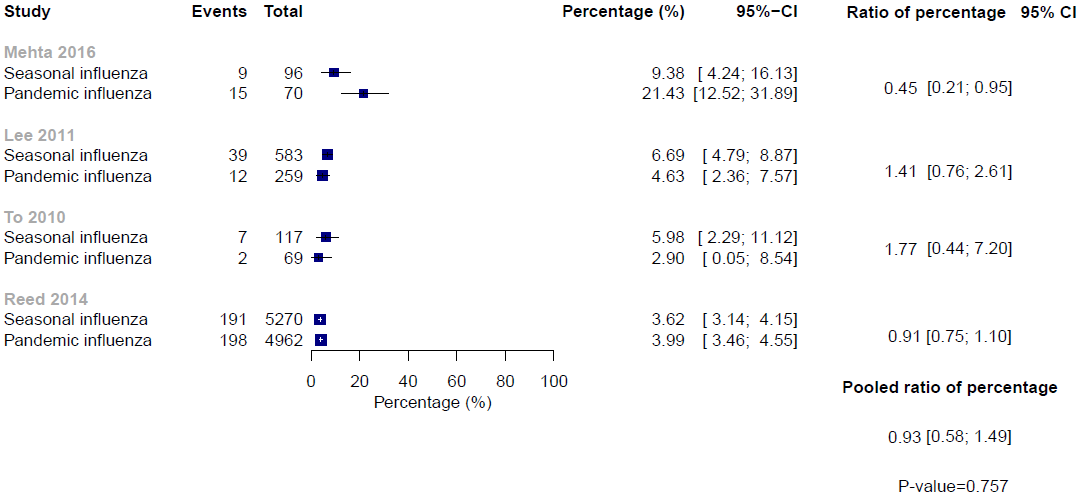

#### 20.2. Between-study subgroup analysis of all-cause mortality rate in severe seasonal or pandemic influenza by influenza etiology

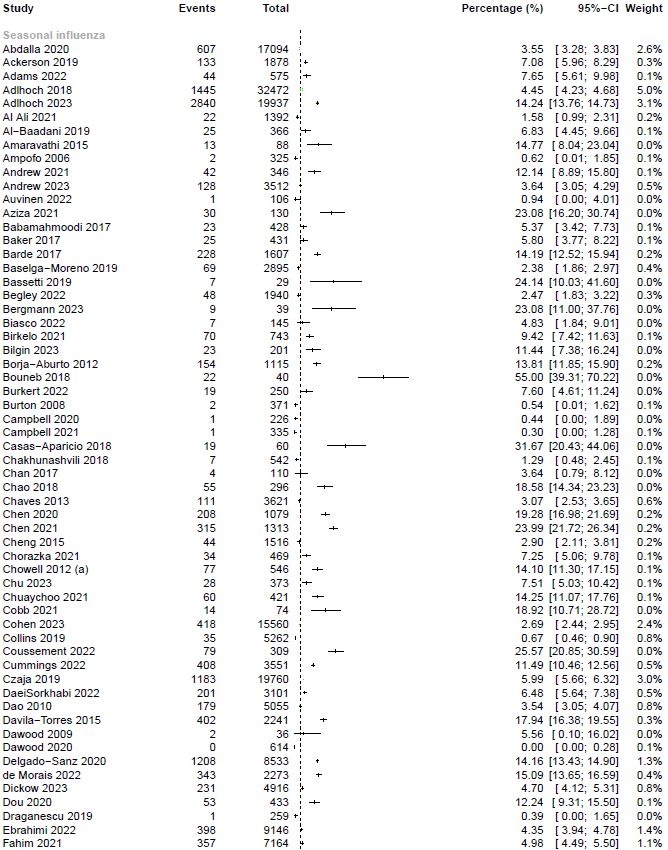

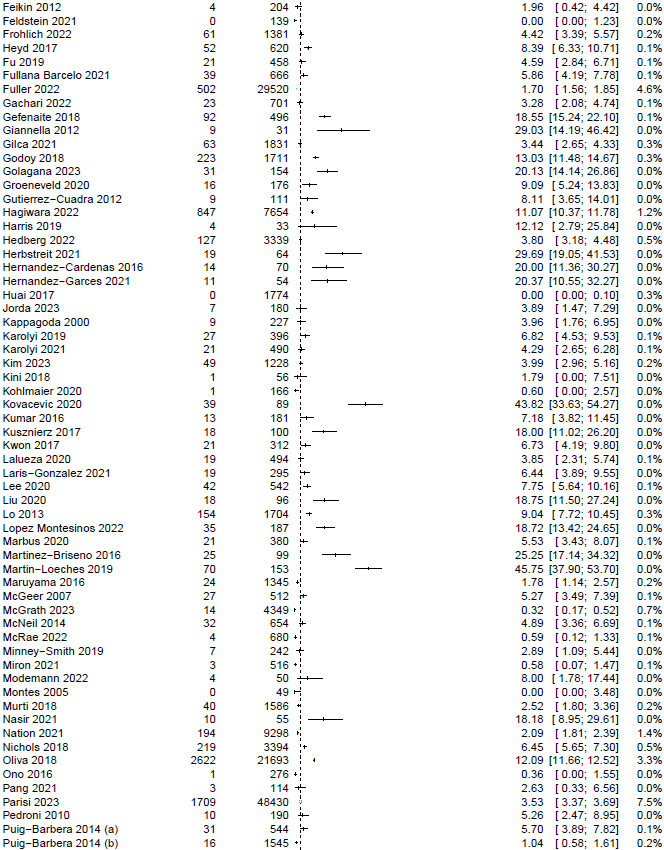

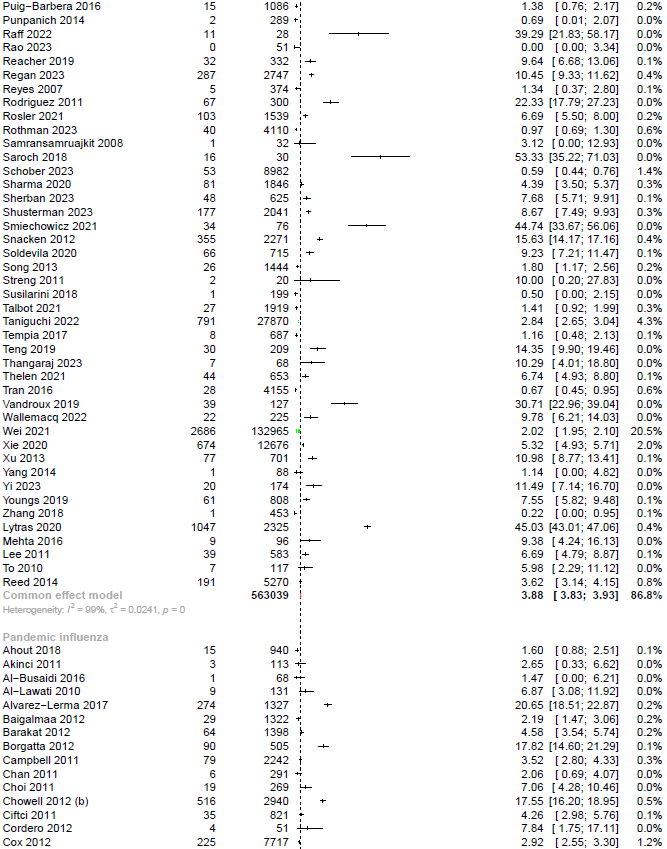

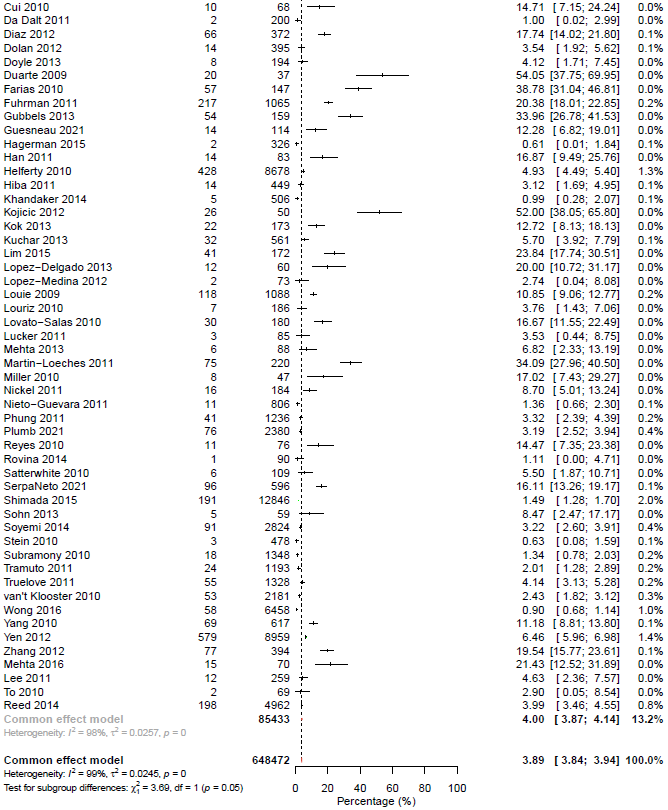

### Appendix 21. Subgroup analysis of all-cause mortality rate in severe seasonal or pandemic influenza by vaccination status

#### 21.1. Within-study subgroup analysis of all-cause mortality rate in severe seasonal or pandemic influenza by vaccination status

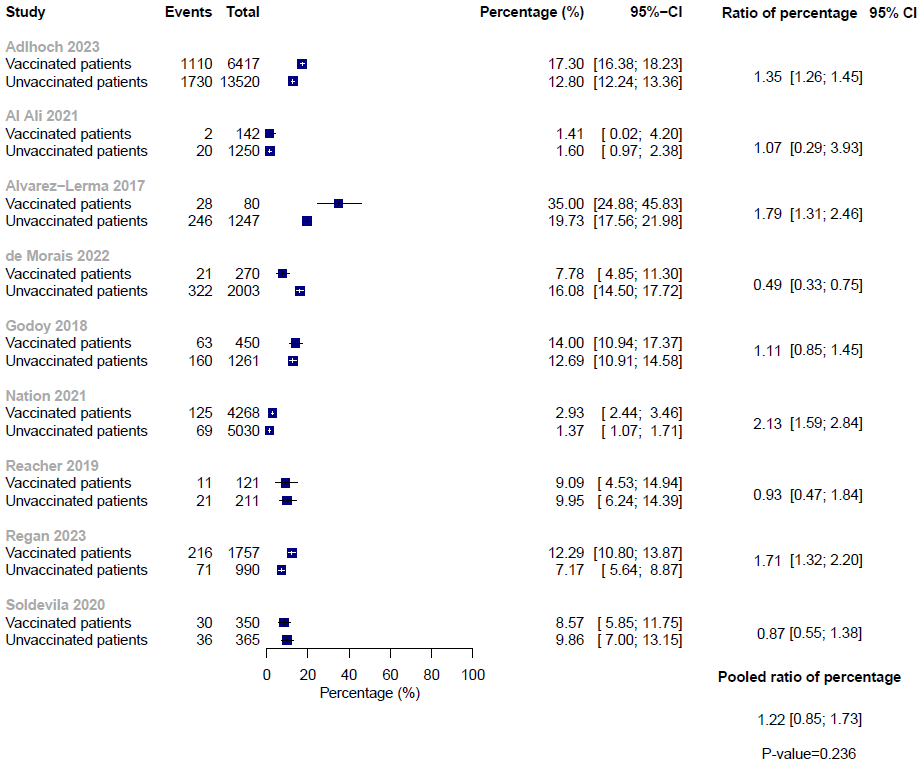

#### 21.2. Between-study subgroup analysis of all-cause mortality rate in severe seasonal or pandemic influenza by vaccination status

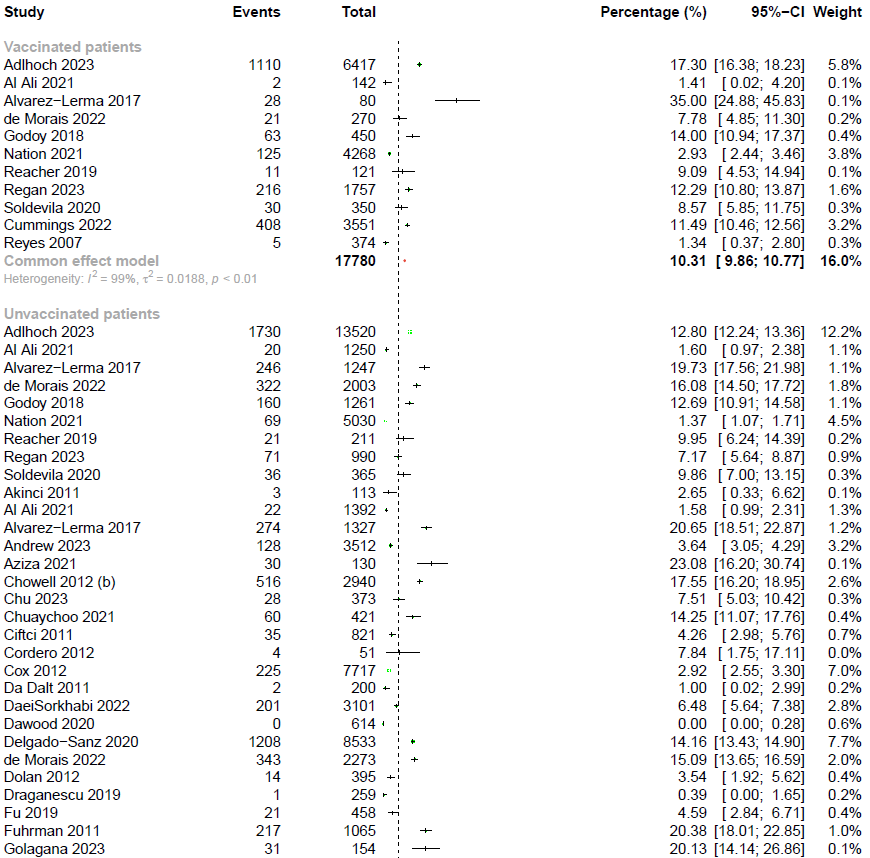

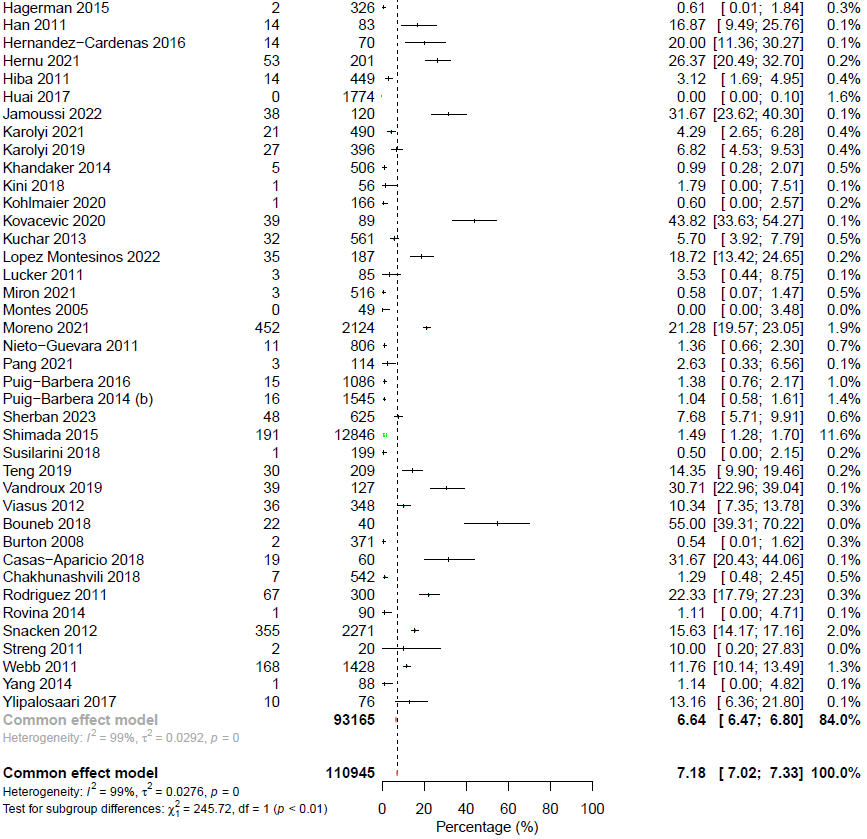

### Appendix 22. Subgroup analysis of all-cause mortality rate in severe seasonal or pandemic influenza by chronic comorbidities

#### 22.1. Within-study subgroup analysis of all-cause mortality rate in severe seasonal or pandemic influenza by chronic comorbidities

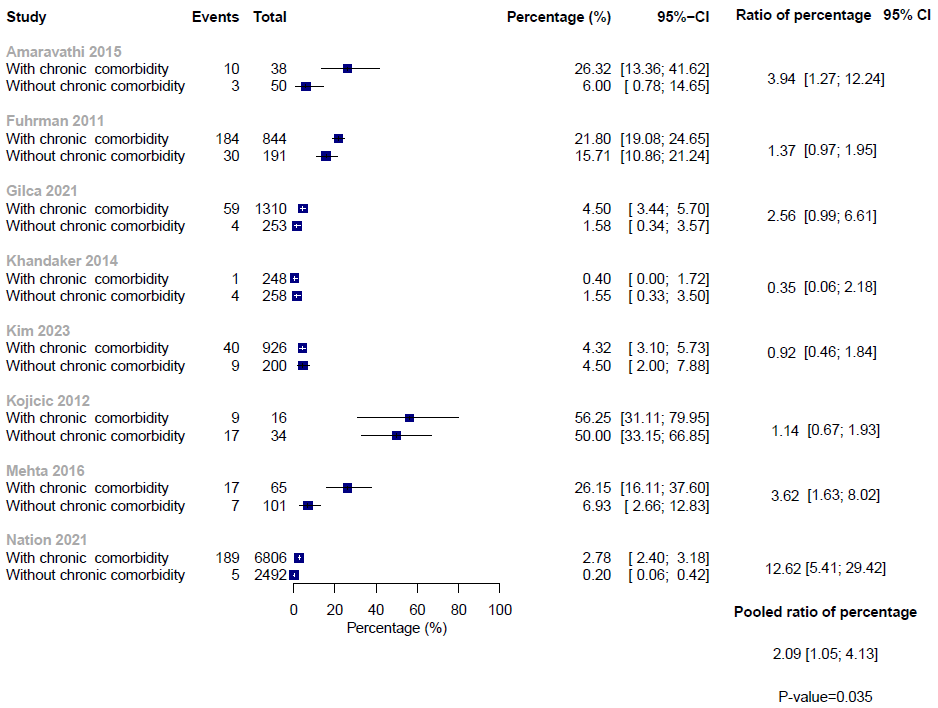

#### 22.2. Between-study subgroup analysis of all-cause mortality rate in severe seasonal or pandemic influenza by chronic comorbidities

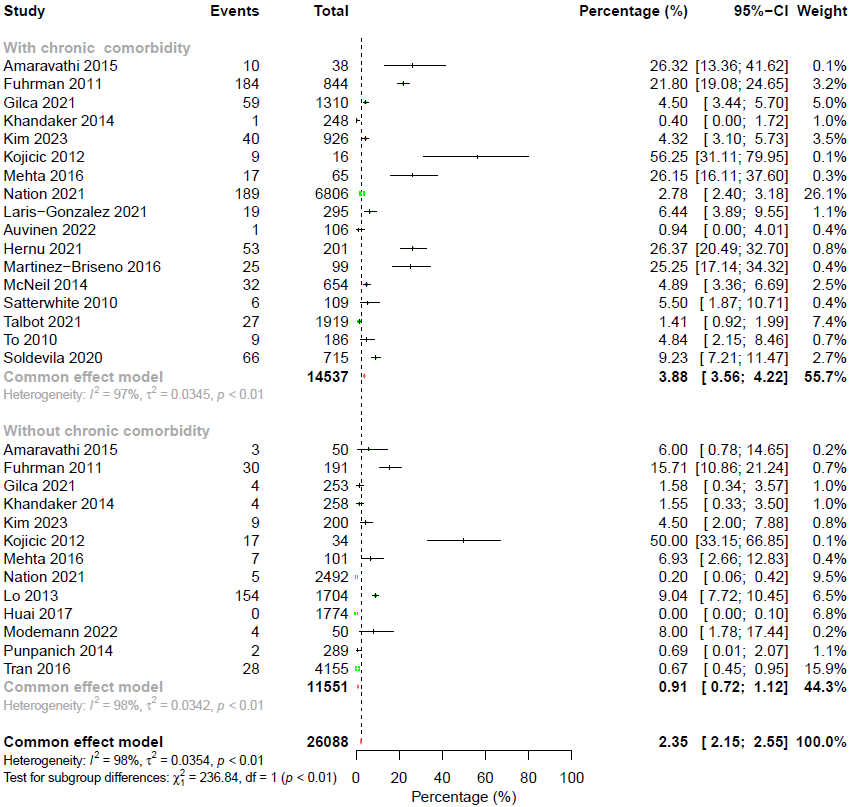

### Appendix 23. Credibility assessment of subgroup analyses for all-cause mortality rate in severe seasonal or pandemic influenza

| **ICEMAN item** | **By age (0-14 years vs 15-64 years)** | **By age (0-14 years vs ≥65 years)** | **By age (15-64 years vs ≥65 years)** | **By countries** | **By chronic comorbidities** |
| --- | --- | --- | --- | --- | --- |
| Is the analysis of effect modification based on comparison within rather than between studies? | Completely within | Completely within | Completely within | Completely between | Completely within |
| For within-study comparisons, is the effect modification similar from study to study? | Definitely similar | Definitely similar | Definitely similar | Not applicable | Mostly similar |
| For between-study comparisons, is the number of studies large? | Not applicable | Not applicable | Not applicable | Large | Not applicable |
| Was the direction of effect modification correctly hypothesised a priori? | Definitely no | Definitely yes | Definitely yes | Definitely yes | Definitely yes |
| Does a test for interaction suggest that chance is an unlikely explanation of the apparent effect modification? | Chance an unlikely explanation | Chance an unlikely explanation | Chance an unlikely explanation | Chance an unlikely explanation | Chance a likely explanation |
| Did the authors test only a small number of effect modifiers or consider the number in their statistical analysis? | Probably no | Probably no | Probably no | Probably no | Probably no |
| Did the authors use a random effects model? | Definitely yes | Definitely yes | Definitely yes | Definitely no | Definitely yes |
| If the effect modifier is a continuous variable, were arbitrary cut points avoided? | Not applicable | Not applicable | Not applicable | Not applicable | Not applicable |
| Are there any additional considerations that may increase or decrease credibility? | Not applicable | Not applicable | Not applicable | Not applicable | Not applicable |
| How would you rate the overall credibility of the proposed effect modification? | Low credibility | Moderate credibility | Moderate credibility | Low credibility | Low credibility |

### Appendix 24. Between-study subgroup analysis of all-cause mortality rate in severe seasonal or pandemic influenza by countries

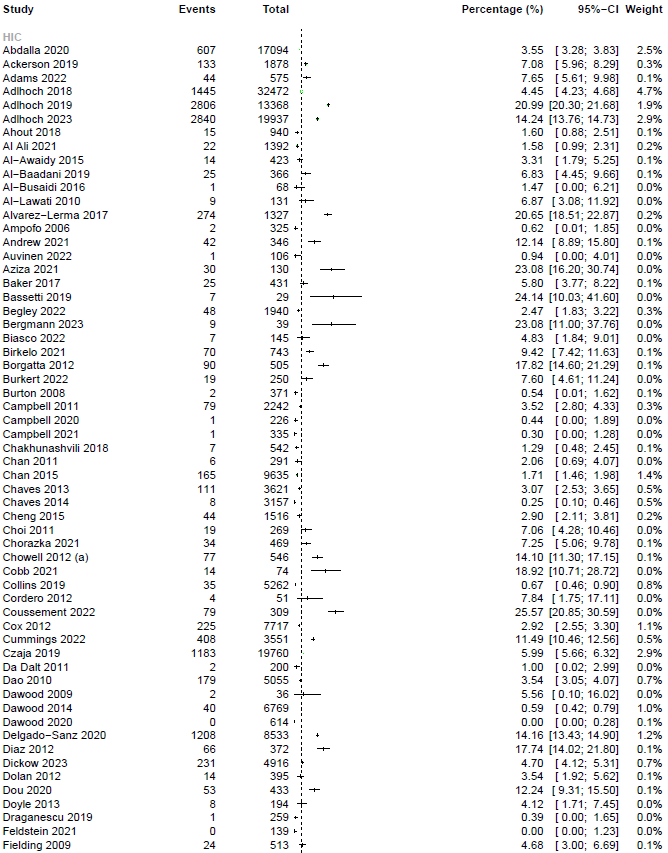

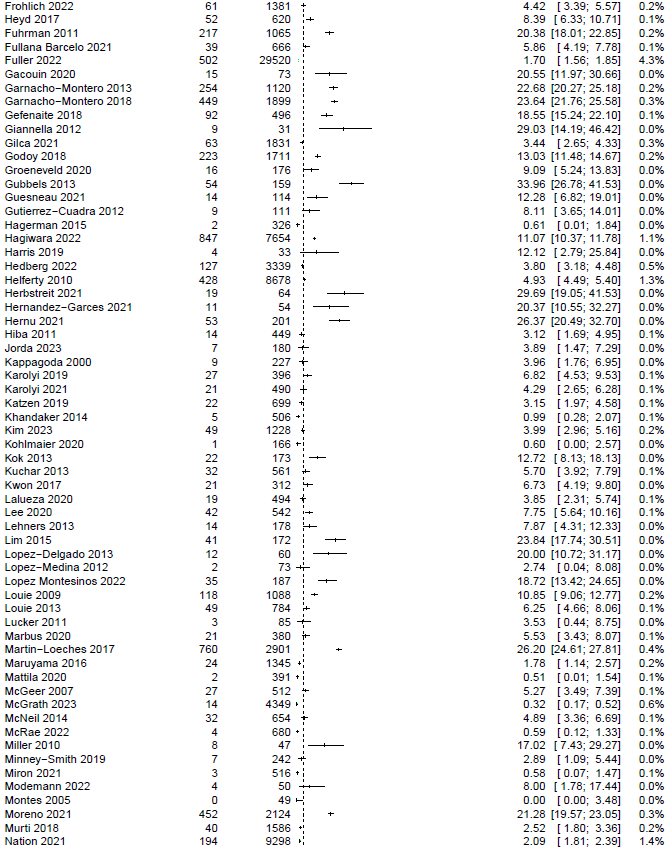

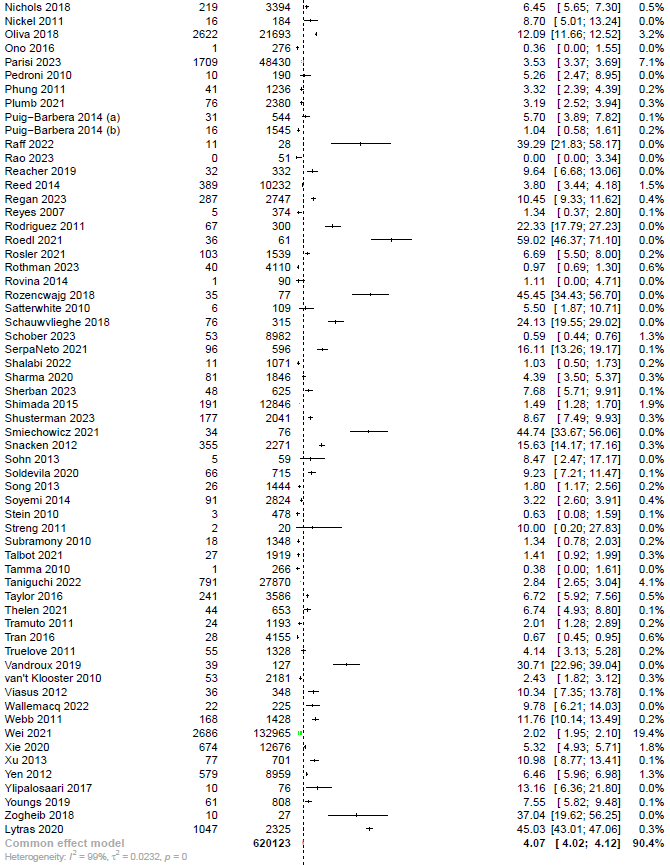

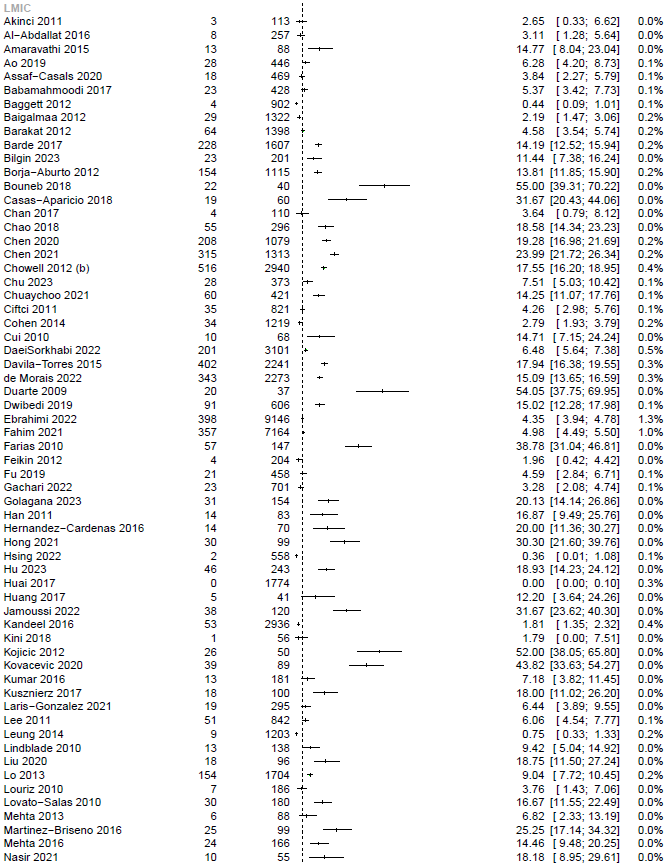

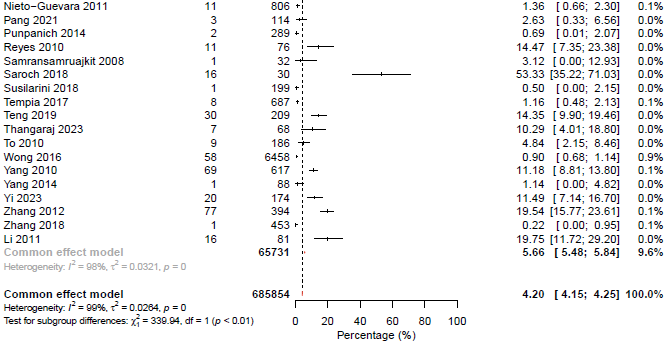

### Appendix 25. Sensitivity analysis only including studies with low risk of bias for all-cause mortality rate in severe seasonal or pandemic influenza

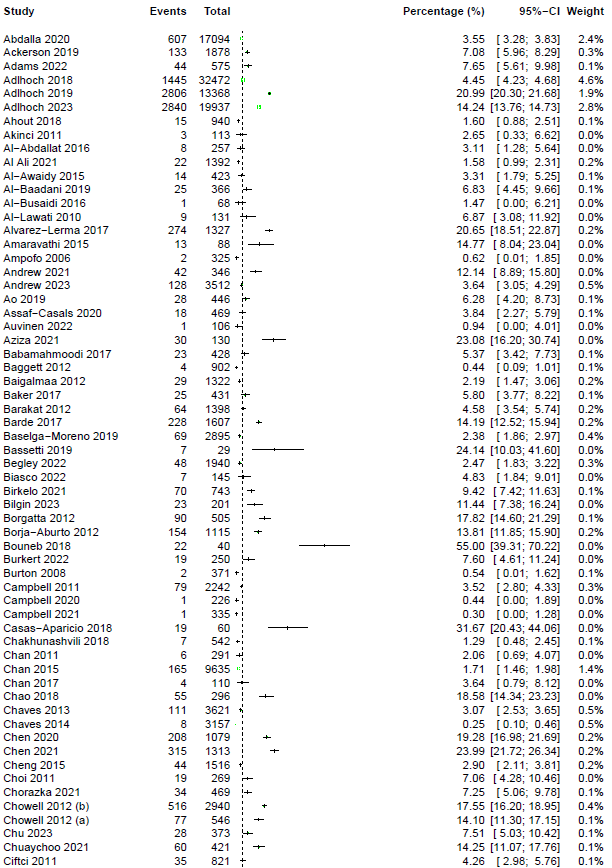

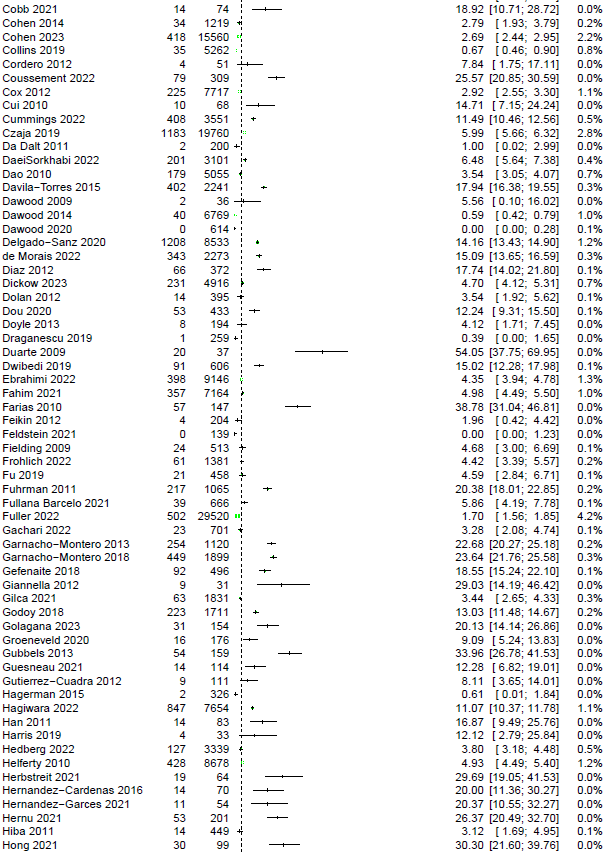

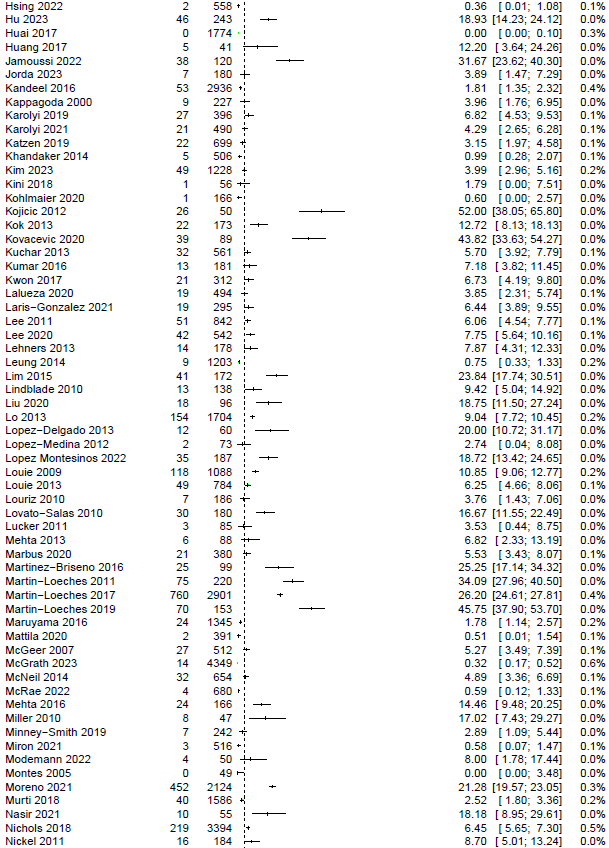

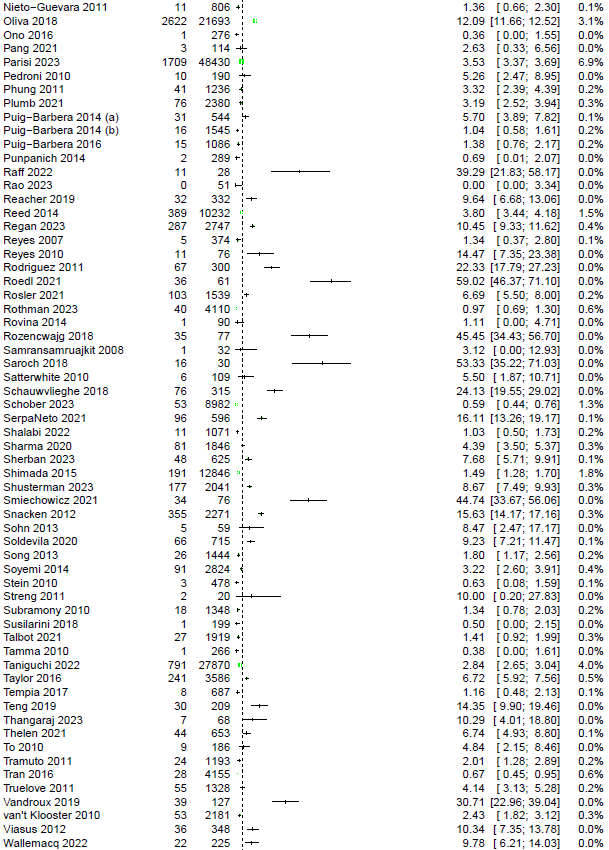

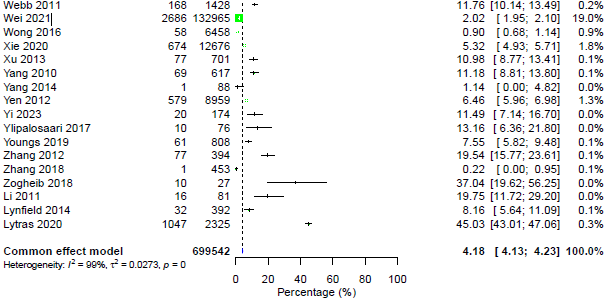

### Appendix 26. Meta-analyses of all-cause mortality rate in zoonotic influenza

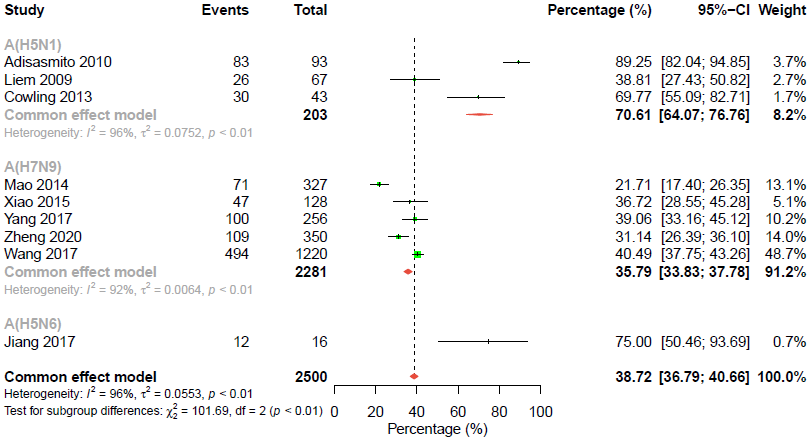
